# The Landscape of Shared and Divergent Genetic Influences across 14 Psychiatric Disorders

**DOI:** 10.1101/2025.01.14.25320574

**Authors:** Andrew D Grotzinger, Josefin Werme, Wouter J Peyrot, Oleksandr Frei, Christiaan de Leeuw, Lucy K Bicks, Qiuyu Guo, Michael P Margolis, Brandon J Coombes, Anthony Batzler, Vanessa Pazdernik, Joanna M Biernacka, Ole A Andreassen, Verneri Anttila, Anders D Børglum, Na Cai, Ditte Demontis, Howard J Edenberg, Stephen V Faraone, Barbara Franke, Michael J Gandal, Joel Gelernter, John M Hettema, Katherine G Jonas, James A Knowles, Karestan C Koenen, Adam X Maihofer, Travis T Mallard, Manuel Mattheisen, Karen S Mitchell, Benjamin M Neale, Caroline M Nievergelt, John I Nurnberger, Kevin S O’Connell, Elise B Robinson, Sandra S Sanchez-Roige, Susan L Santangelo, Hreinn Stefansson, Kari Stefansson, Murray B Stein, Nora I Strom, Laura M Thornton, Elliot M Tucker-Drob, Brad Verhulst, Irwin D Waldman, G Bragi Walters, Naomi R Wray, Anxiety Disorders Working Group, Attention-Deficit/Hyperactivity Disorder (ADHD) Working Group, Autism Spectrum Disorders Working Group, Bipolar Disorder Working Group, Eating Disorders Working Group, Major Depressive Disorder Working Group, Nicotine Dependence GenOmics (iNDiGO) Consortium, Obsessive-Compulsive Disorder Working Group, Post-Traumatic Stress Disorder Working Group, Schizophrenia Working Group, Substance Use Disorders Working Group, Tourette Syndrome Working Group, Phil H Lee, Kenneth S Kendler, Jordan W Smoller

## Abstract

Psychiatric disorders display high levels of comorbidity and genetic overlap^1,2^. Genomic methods have shown that even for schizophrenia and bipolar disorder, two disorders long-thought to be etiologically distinct^3^, the majority of genetic signal is shared^4^. Furthermore, recent cross-disorder analyses have uncovered over a hundred pleiotropic loci shared across eight disorders^5^. However, the full scope of shared and disorder-specific genetic basis of psychopathology remains largely uncharted. Here, we address this gap by triangulating across a suite of cutting-edge statistical genetic and functional genomic analyses applied to 14 childhood- and adult-onset psychiatric disorders (1,056,201 cases). Our analyses identify and characterize five underlying genomic factors^6^ that explain the majority of the genetic variance of the individual disorders (∼66% on average) and are associated with 268 pleiotropic loci. We observed particularly high levels of polygenic overlap^7^ and local genetic correlation^8^ and very few disorder-specific loci^9^ for two factors defined by: (*i*) schizophrenia and bipolar disorder (“SB factor”), and by (*ii*) major depression, PTSD, and anxiety (“internalizing factor”). At the functional level, we applied multiple methods^10–12^ which demonstrated that the shared genetic signal across the SB factor was substantially enriched in genes expressed in excitatory neurons, whereas the internalizing factor was associated with oligodendrocyte biology. By comparison, the genetic signal shared across all 14 disorders was enriched for broad biological processes (e.g., transcriptional regulation). These results indicate increasing differentiation of biological function at different levels of shared cross-disorder risk, from quite general vulnerability to more specific pathways associated with subsets of disorders. These observations may inform a more neurobiologically valid psychiatric nosology and implicate novel targets for therapeutic developments designed to treat commonly occurring comorbid presentations.

---

Approximately half the US population will meet criteria for at least one psychiatric disorder during their lifetime,^13^ with most concurrently meeting criteria for several disorders.^1^ High levels of psychiatric comorbidity have presented long standing challenges in defining disorder boundaries. These challenges are heightened by the reality that psychiatric disorders are defined by expert opinion and epidemiological research, as the underlying pathophysiologies remain largely unknown. Rapid progress in psychiatric genomics has uncovered hundreds of loci and biological pathways associated with individual psychiatric disorders. As comorbidity is the norm, not the exception, it is unsurprising that genetic liability is typically correlated among psychiatric disorders and that many genetic loci exhibit pleiotropic (i.e. shared) associations across disorders.^14^

Recent advances in genomic methods offer the critical opportunity to formally characterize genetic risk pathways as impacting most or all disorders, being more relevant to disorder subclusters, or even disorder-specific. The current analyses reflect the third major effort from the Psychiatric Genomics Consortium-Cross Disorder working group (CDG3; for prior efforts see CDG1^15^ and CDG2^5^), and collectively represent the best-powered, most comprehensive cross-disorder analysis to-date. Here, we analyzed 14 psychiatric disorders with genome-wide association study (GWAS) data, triangulating across multiple, complementary analytic approaches to dissect the genetic architecture across these 14 disorders at the genome-wide, regional, functional, and individual genetic variant levels of analysis. Our results offer a nuanced mapping of the psychiatric genetic landscape with implications for diagnosis, pathophysiology, and, potentially, treatment.

## Results

### GWAS Data for 14 Psychiatric Disorders

We included data from the most recent available GWAS of 14 psychiatric disorders (**Table 1**), reflecting a major update relative to the prior CDG2 analyses^5^ (average case increase of ∼165% compared to CDG2; **Suppl. Fig. 1**). This included new GWAS for all eight disorders from CDG2: attention-deficit/hyperactivity disorder (ADHD), anorexia nervosa (AN), autism spectrum disorder (ASD), bipolar disorder (BD), major depression (MD), obsessive-compulsive disorder (OCD), schizophrenia (SCZ), and Tourette’s syndrome (TS).^16–23^ Importantly, we also included six additional disorders: alcohol use disorder (AUD),^24^ anxiety disorders (ANX),^25^ cannabis use disorder (CUD),^26^ nicotine dependence as assessed using the Fagerström Test for Nicotine Dependence (NIC),^27^ opioid use disorder (OUD),^28^ and post-traumatic stress disorder (PTSD).^29^ Due to the uneven representation of diverse ancestral groups across the 14 disorders, the full set of cross-disorder analyses was restricted to a single genetic ancestry group, which we refer to as European (EUR) samples, with this group defined based on genetic similarity to one another (e.g., on principal components) or similarity to global reference panels^30^. We also report bivariate results for MD^31^ and SCZ^32^ in East Asian (EAS) genetic ancestry groups and AUD,^33^ CUD,^26^ OUD,^28^ and PTSD^29^ in African/African American (AAM) genetic ancestry groups defined using similar procedures.

**Table 1.**
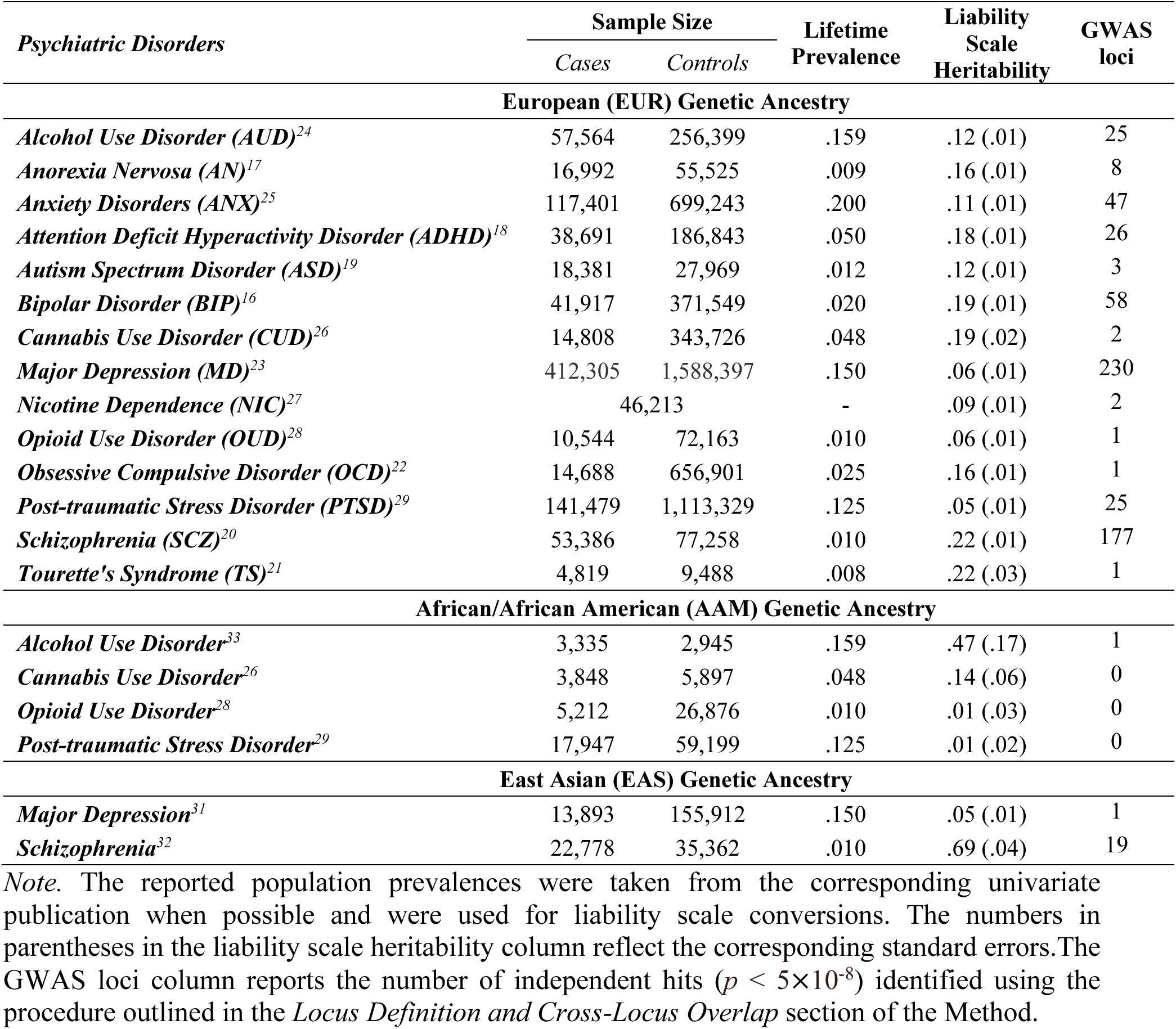
Summary of Psychiatric Disorder Datasets.

### Genetic Correlations Emphasize Importance of Cross-Ancestry Analyses

Genetic correlations (*r_g_*) estimated with linkage disequilibrium (LD) score regression (LDSC)^4^ revealed pervasive and substantial genetic overlap across disorders, with clusters of disorders demonstrating particularly high genetic overlap in individuals of EUR genetic ancestry (**Fig. 1; Online Suppl.; Suppl. Table 1**; see **Suppl. Figs. 2-4** for consideration of especially high *r_g_* across PTSD and MD). The LDSC estimates within AAM participants were all nonsignificant, most likely due to limited power (**Suppl. Table 4)**. Though, interestingly, the *r_g_* between MD and SCZ in EAS populations (*r*_g_ = 0.45; *SE* = 0.09) was double that observed in EUR participants (*r*_g_ = 0.22; *SE* = 0.04). This divergence is not fully understood, but highlights the importance of investing in genetic efforts for other ancestry groups.

**Figure 1.**
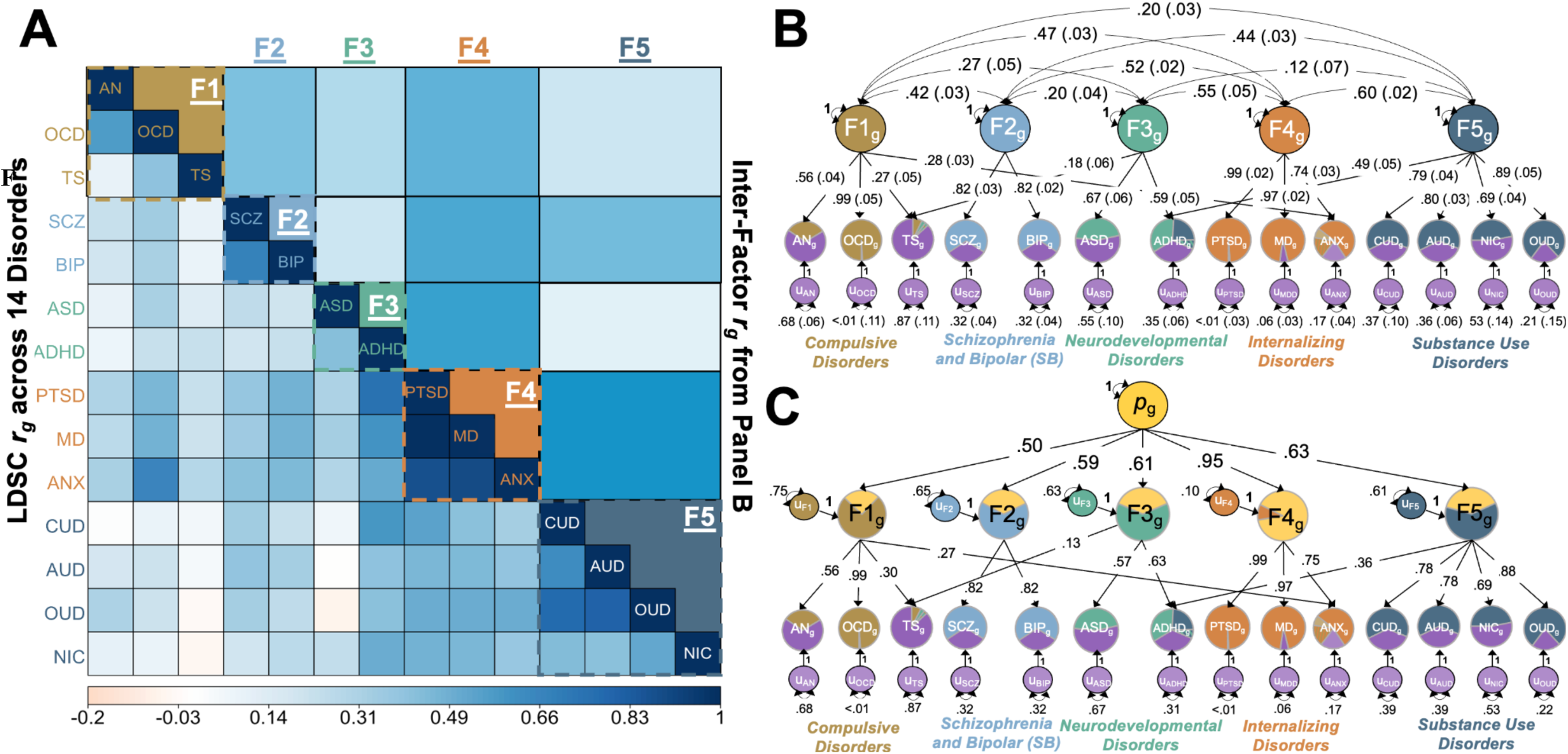
Genome-wide structural models. *Panel A* depicts the heatmap of *r*g’s across the 14 disorders as estimated using LD-score regression (LDSC) on the lower diagonal and the correlations among the psychiatric genomic factors as estimated using GenomicSEM above the diagonal. Disorders that load on the same factor are depicted in the same color. LDSC estimates were used as input to Genomic SEM to produce the results in the remaining two panels. *Panel B* depicts estimates from the five-factor model along with standard errors in parentheses. Estimates are standardized relative to the SNP-based heritabilities, where the total variance in the disorders is equal to the sum of the squared factor loading (the single-headed arrow(s) from the factor to the disorder) and the residual variance (the values on the double-headed arrows on the single-color circles). Disorders are shown as pie charts with the proportion of unique (residual) variance shaded in purple and the variance explained by the psychiatric factors in the color of the corresponding factor. *Panel C* displays the standardized estimates from the *p*-factor model. The disorders are again color coded in blue and orange as in *Panel B* and the first-order factors (F1-F5) additionally color coded to show variance explained by the second-order, *p*-factor in yellow. ADHD = attention-deficit hyperactivity disorder; ASD = autism spectrum disorder; OCD = obsessive compulsive disorder; SCZ = schizophrenia; TS = Tourette’s syndrome; MD = major depression; AN = anorexia nervosa; BIP = bipolar disorder; ANX = anxiety disorder; PTSD = post-traumatic stress disorder; AUD = alcohol use disorder; OUD = opioid use disorder; NIC = nicotine dependence; CUD = cannabis use disorder.

As the majority of analyses were restricted to participants of EUR genetic ancestry, we sought to gauge how generalizable our findings were across ancestry groups. This was achieved using Popcorn,^34^ which can be applied to estimate *r*_g_’s for the same trait across different ancestry groups. We specifically calculated the genetic impact correlation (ρ*_gi_*), which considers different allele frequencies across populations by calculating the correlation between the population-specific, allele-variance normalized SNP effect sizes. Results were underpowered for many of the comparisons, but included a strong EAS-EUR correlation for SCZ (ρ*_gi_* = 0.85, *SE* = 0.04), followed by a more moderate correlation between EAS and EUR for MD (ρ*_gi_* = 0.67, *SE* = 0.16), and finally the lowest estimate for AAM-EUR and PTSD (ρ*_gi_* = 0.59, *SE* = 0.27; **Suppl. Table 4**). These results indicate that our broader set of EUR findings may generalize better for some disorders (e.g., SCZ) than for others (e.g., PTSD). Of note, previously reported within-disorder, within-ancestry *r*_g_’s calculated across cohorts are considerably smaller for PTSD (*r*_g_ = 0.73, *SE*=0.21)^35^ and MD (*r*_g_ = 0.76, *SE* =0.03)^36^ than SCZ (*r*_g_ = 0.95, *SE* = 0.03)^37^, suggesting that cross-ancestry *r*_g_’s for these two disorders reflect a combination of etiological and phenotypic heterogeneity and ancestry-specific signal.

### MiXeR Identifies Pervasive Polygenic Overlap across Disorders

Genome-wide *r*_g_’s from LDSC indicate shared genetic risk across psychiatric disorders. However, LDSC may underestimate the full extent of genetic overlap if shared causal variants reflect a mixture of directionally concordant and discordant associations across traits. To account for this, we applied bivariate causal mixture modeling (MiXeR), which quantifies the degree of polygenic overlap reflecting the total number of shared causal variants irrespective of magnitude or directionality.^7^ Cross-trait analyses were limited to MD, SCZ, BIP, ANX, ADHD, PTSD, AUD, and AN, because the remaining disorders were underpowered for these analyses (**Method**; results for univariate MiXeR are reported in the Online Supplement; **Suppl. Table 5**; **Suppl. Fig. 5**). **Fig. 2** displays cross-trait MiXeR results for pairwise overlap across four particularly well-powered disorders: ADHD, SCZ, BIP, and MD (**Suppl. Figs. 6-9** and **Suppl. Table 6** for complete results). These findings indicate extensive polygenic overlap across psychiatric disorders that is greater than that suggested by the already high *r*_g_’s from LDSC. Overall, the results from our MiXeR analyses suggest that the genetic signal for psychiatric disorders primarily reflects variants with convergent effects across disorders, and that genetic risk is differentiated by a more limited number of shared variants with discordant effects.

**Figure 2.**
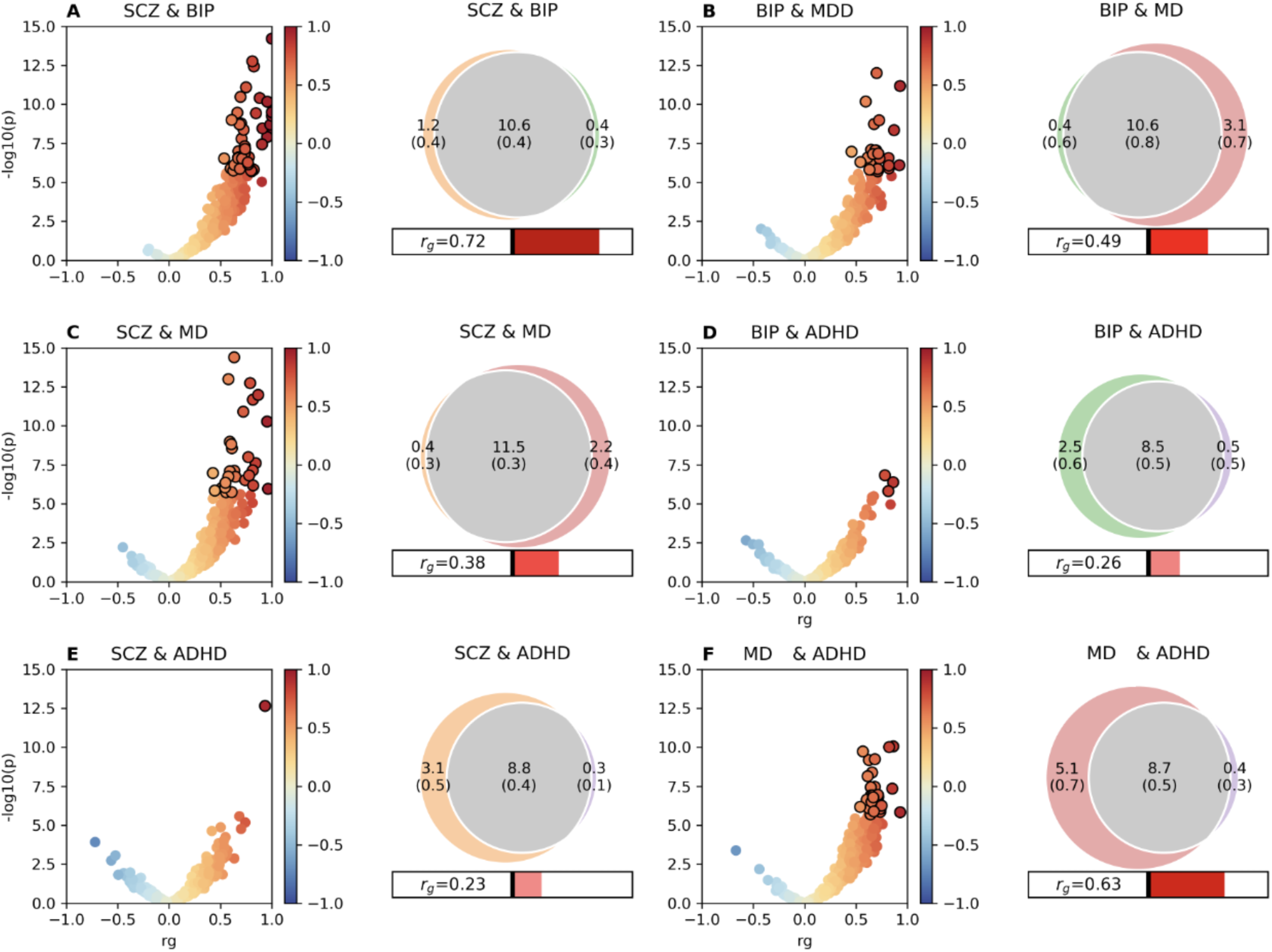
LAVA and MiXeR results for select disorders. Results are shown for LAVA (volcano plots) and MiXeR (Venn diagrams) for four of the most well-powered disorders: schizophrenia (SCZ), bipolar disorder (BIP), major depression (MD), and attention-deficit/hyperactivity disorder (ADHD). LAVA results are shown as a volcano plot of the local genetic correlations (*rg*) on the x-axis and -log10(*p*) values on the y-axis. Stronger negative *r*g’s are depicted by darker shades of blue, while stronger positive *r*g’s are shown in darker shades of red. The points for local *r*g’s that were significant at a Bonferroni corrected threshold are depicted with black outlines. MiXeR results are shown as adjacent Venn diagrams that convey unique and shared polygenic components at the causal level. The numbers within the Venn diagrams indicate the estimated quantity of causal variants (in thousands) per component, explaining 90% of SNP heritability in each phenotype, followed by the standard error. The size of the circles reflects the degree of polygenicity. *r*g is represented in the horizontal bars beneath the Venn diagrams.

### Genomic SEM Identifies Five Factors Underlying 14 Disorders

We used Genomic Structural Equation Modeling (Genomic SEM)^6,10^ in the EUR genetic ancestry datasets to model genetic overlap from LDSC across 14 disorders as latent factors, where the latent factors represent dimensions of shared genetic risk **(Method)**. Best fit was reached with a five-factor model (**Suppl. Tables 2-3**) that provided good fit to the data (Comparative Fix Index [CFI] = .971, Standard Root Mean Square Residual [SRMR] = .063). These five latent genomic factors (**Fig. 1**) comprised: (*F1*) a **Compulsive disorders factor** defined by AN, OCD, and, to a considerably lesser extent, TS and ANX; (*F2*) a **Schizophrenia and Bipolar (SB)** factor defined by BIP and SCZ; (*F3*) a **Neurodevelopmental disorders factor** defined by ASD, ADHD and, to a much lesser extent, TS; (*F4*) an **Internalizing disorders factor** defined by PTSD, MD, and ANX; and (*F5*) a **Substance Use factor** defined by OUD, CUD, AUD, NIC, and, to a much lesser extent, ADHD.

Within this five-factor model, the Internalizing and Substance Use factor displayed the highest inter-factor correlation (*r*_g_ = 0.60; *SE* = 0.02). The median residual genetic variance unexplained by the latent genomic factors was 33.5%, indicating that the majority of genetic risk is shared among disorder subsets. TS notably displayed the most unique genetic signal, with 87% of its genetic variance unexplained by the factors. The factor structure was very similar to that found by Genomic SEM applied to subsets of these disorders in prior work^5,10^, indicating stability in the underlying factor structure, even as sample sizes have increased and more disorders are included.

Evidence of moderate *r*_g_ between the factors suggested that a higher-order factor may explain common variance across the correlated factors. In line with this observation, we found that a hierarchical model also fit the data well (CFI = .959, SRMR = .074). We refer to this as the *p-* factor model as it consisted of a higher-order general psychopathology factor (*p-*factor) defined by the five, lower-order psychiatric factors from the five-factor model. The five-factor and *p-*factor models were brought forward for the remaining Genomic SEM analyses.

### LAVA Analyses Reveal Local Hotspots of Overlapping Genetic Effects

To account for potential heterogeneity in genetic overlap across the genome, we applied local analysis of [co]variant association (LAVA)^8^ to examine the pairwise local *r*_g_ between disorders at specific genomic regions and identify potential pleiotropy hotspots. We segmented the genome into 1,093 LD-independent regions with an average width of approximately 2.5 Mb, and restricted our analysis to loci with sufficient SNP-based heritability for the disorders analyzed (*p* < 4.6×10^-5^ = 0.05/1,093; **Method**). Correcting for the number of bivariate tests performed across all loci and disorder pairs, we detected a total of 458 significant pairwise local *r*_g_’s (*p* < 2.1×10^-6^ = 0.05/24,273). Consistent with the multivariate genetic structure identified using Genomic SEM, the pairs of disorders with the greatest number of significant local *r_g_* hits were MD and ANX (113 *r_g_* loci), MD and PTSD (88 *r_g_* loci), and BIP and SCZ (40 *r_g_* loci), accounting for just over half of all significant local *r_g_*’s detected (**Fig. 3A**).

**Figure 3.**
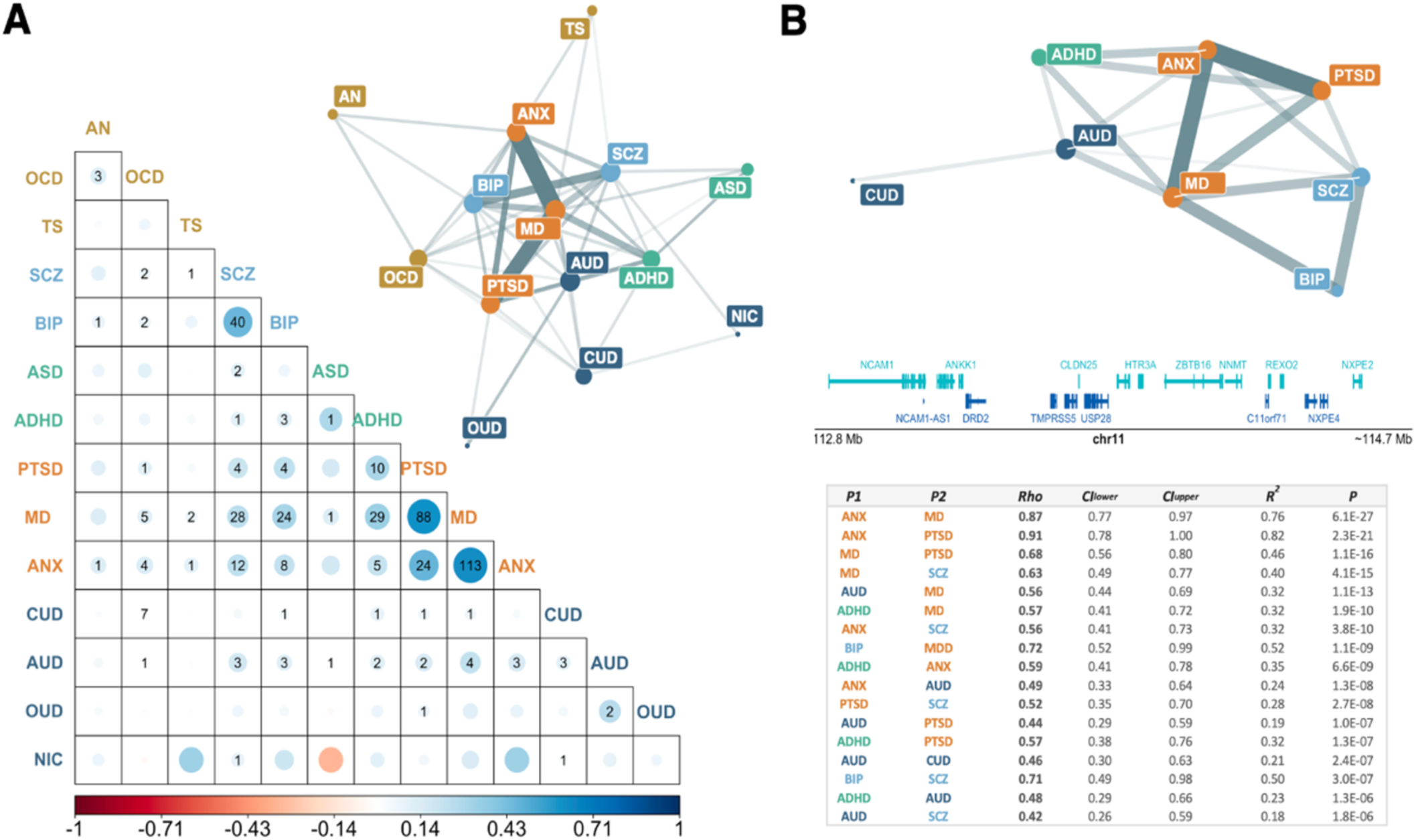
Local genetic correlations. Panel. **A** displays an overview of the average patterns of local genetic correlations (*r*g’s) across the genome for all pairs of disorders in the form of a heatmap (below diagonal) and a network plot (above diagonal). The colors of the heatmap represent the average local *r*g’s across all evaluated loci, with dot size reflecting the strengths of average associations, and numbers indicating how many of the local *r*g’s were significant. These results are mirrored in the network plot, where the width or the edges reflect the number of significant associations, meaning that only disorders with at least one significant local *r*g are connected, and the edge opacity the strength of the average local *r*g across tested loci. Note that label colors are concordant with the Genomic SEM factor structure from Fig. 1 and, as shown, disorders of similar colors also tend to be proximally located within the network. **Panel B** displays the local *r*g structure within the top *r*g hotspot on chromosome 11 (112,755,447-114,742,317 [GRCh37]), i.e. the locus where the greatest number of significant *r*g’s were found across all disorder pairs. Here, the network plot illustrates all significant *r*g’s detected in this locus, with both edge width and opacity reflecting the strength of the association. The region plot in the middle displays the genes contained within the hotspot, and the table below shows the genetic correlation estimates (Rho), 95% confidence intervals (CIlower, CIupper), variance explained (*R^2^*) and *p*-values (P) for all significant pairwise local *r*g’s in this locus. Label colors are concordant with those used for the Genomic SEM factor structure in Fig. 1.

Furthermore, we detected 101 regions that harbored significant local *r*_g_’s between several disorder pairs, which we call “*r_g_* hotspots” (**Suppl. Tables 6-15** for local *r_g_* across disorders in the top 10 hotspots). The most pleiotropic of these hotspots, as indicated by the number of significant *r_g_*’s detected, was on chromosome 11, which contained 17 significant local *r_g_*’s involving 8 of the 14 analyzed disorders (**Fig. 3B**). This locus also stands out as being the most significantly associated with 8 of the 17 disorder pairs, while ranking in the top 25% of associated loci for 12 of the 17 disorder pairs (**Suppl. Fig. 12**). Notably, this region has previously been flagged as a potential pleiotropy hotspot for cognitive and psychiatric phenotypes^8,38,39^, and contains the *NCAM1*-*TTC12*-*ANKK1*-*DRD2* gene cluster that is frequently associated with psychiatric outcomes^40–43^.

The average estimated local *r_g_*’s from LAVA were generally consistent with the global estimates from LDSC. Additionally, both global and local *r*_g_’s tended to be positive, indicating that, on average, the shared genetic risk for one disorder also tends to increase the risk of another (**Suppl. Fig. 10**). Essentially all significant local *r_g_*’s detected with LAVA were also positive, with significant negative *r*_g_’s identified in only three instances (**Suppl. Fig. 11**).

### Multivariate GWAS Identifies 428 Cross-Disorder Risk Variants

Focusing on the locus (individual genetic variant) level, we used multivariate GWAS within Genomic SEM^6^ to identify single nucleotide polymorphisms (SNPs) associated with the factors from the five-factor model and the *p*-factor. We also estimated factor-specific Q_SNP_ heterogeneity statistics that index SNPs that deviate strongly from the factor structure. Q_SNP_ is often significant for SNPs with highly disorder-specific or directionally discordant effects (**Method**). Q_SNP_ for the *p*-factor will also capture SNPs specific to, or with divergent effects across, the five lower-order factors. We defined significant hits for the factors as those that were significant after Bonferroni correction (*p* < 5×10^-8^/6_Genomic Factors_) and not within 100 kb of a Q_SNP_ hit for that same factor. We excluded SNPs physically proximal to Q_SNP_ hits as these are likely driven by disorder-specific effects that do not operate via the factor. Using these criteria, we identified 268 independent loci with pleiotropic effects from the five-factor model. The majority of these hits were for the SB (102 independent hits within the factor) and Internalizing factors (150 hits). There were 160 hits for the *p-*factor (**Fig. 4**; **Suppl. Fig. 13; Suppl. Tables 17-29**). Results across both models reflected 48 novel loci (across 428 total loci) relative to the univariate GWAS used as input.

**Figure 4.**
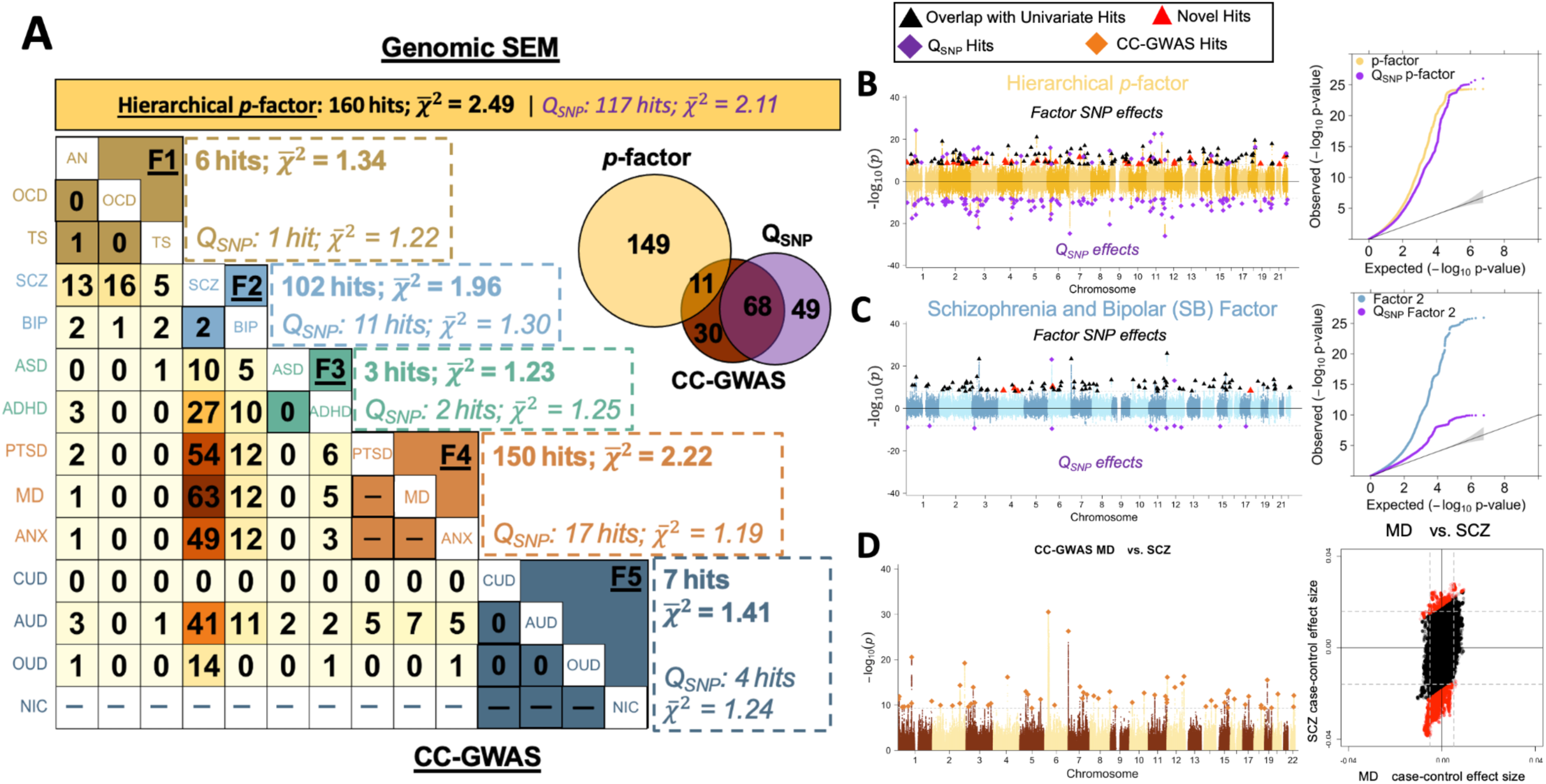
Locus level results. *Panel A* depicts a heatmap of CC-GWAS loci below the diagonal across pairwise combinations of disorders, where darker orange shading indicates a higher number of CC-GWAS hits. CC-GWAS results are not shown for the Internalizing disorders as their genetic correlations were too high or for nicotine dependence as this is a continuously measured trait. Genomic SEM results (number of hits and mean chi-square for each factor and factor-specific QSNP estimate) are reported above the diagonal. Results for the *p*-factor are depicted above the plot in yellow, along with a Venn diagram of overlap between *p*-factor, *p*-factor QSNP, and overall CC-GWAS hits. The disorders are ordered and colored according to the Genomic SEM factor structure from Fig. 1. *Panels B* and *C* depict the Miami and QQ-plots for the *p*-factor and SBs factors, respectively. These panels depict results for the -log10(*p*-values) for the factor on the top half of the Miami plot and the log10(*p*-values) for QSNP on the bottom half. Per the legend at the top of this set of panels, factors hits that were within 100 kb of univariate hits are shown in black triangles, novel hits for the factors that were not within 100 kb of a univariate or QSNP hit are depicted as red triangles, and QSNP hits as purple diamonds. The gray dashed line indicates the Bonferroni corrected, genome-wide significance threshold. Panel D depicts the Manhattan plot for the CC-GWAS comparison across major depression (MD) and schizophrenia (SCZ), which produced the most hits (depicted as orange diamonds), as well as the scatterplot of standardized case-control effect sizes of MD (x-axis) vs SCZ (y-axis), with CC-GWAS significant SNPs labeled in red.

We identified 33 independent loci with significant Q_SNP_ effects across the factors from the five-factor model. By comparison, we identified 117 independent Q_SNP_ loci from the *p-*factor model that showed significantly divergent effects across the five, lower-order psychiatric factors (**Suppl. Table 29**). These *p-*factor Q_SNP_ hits also included the chr 11 LAVA hotspot, where this region was found not to confer transdiagnostic risk via the *p-*factor due to an absence of signal for the Neurodevelopmental disorders factor. For the Substance Use factor, highly significant Q_SNP_ hits were driven by variants in the genes involved in well-described biological pathways specific to the metabolism of a particular substance, including the alcohol dehydrogenase genes (*ADH1A, ADH1B, ADH1C*) for AUD and the α-5 subunit of the nicotinic acetylcholine receptor (*CHRNA5*) gene for NIC. It is notable that the number of Q_SNP_ loci substantially greater in the *p-*factor model relative to the five-factor model, indicating that shared genetic relationships are better captured by the five factors (**Suppl. Figs. 14-15**).

PheWAS conducted in the Mayo Clinic Biobank using the factor hits as input revealed that loci discovered for the factors were associated with multiple psychiatric disorders, especially those that loaded on the corresponding factor (**Online Suppl.; Suppl. Table 30; Suppl. Fig. 16**). Notably, the *p-*factor (**Suppl. Fig. 16f**) and Internalizing factor (**Suppl. Fig. 16fd**) loci were also associated with a range of medical outcomes (e.g., chronic pain conditions, hypertension).

### Case-Case GWAS Reveals 412 Loci with Disorder-Specific Effect Between Disorder Pairs

In more fine-grained analyses of disorder pairs, Case-Case GWAS (CC-GWAS)^9^ was applied to identify loci with different allele frequencies across cases of different disorders. Such loci may reflect distinctive, rather than shared, genetic effects across disorder pairs. CC-GWAS was applied to a total of 75 disorder pairs, comparing 13 disorders (excluding NIC because it is a continuous trait). The pairs ANX-MD, ANX-PTSD, and MD-PTSD were excluded as these all had an *r*_g_ estimate > 0.8, thereby risking an inflated Type I error rate (**Method**). The genome-wide significance threshold was defined at 5.5×10^-10^ (i.e., 5×10^-8^/91 pairwise comparisons). An overview of CC-GWAS input parameters is provided in **Suppl. Table 31**.

A total of 412 loci showed significantly different effects across the 75 disorder pairs (**Suppl. Tables 31-33**); most of these loci (294/412) were in comparisons that included SCZ, which may reflect greater power for the GWAS of SCZ or more distinctive biology for this disorder. Due to overlap among the pairwise CC-GWAS hits, these 412 loci comprised 109 LD-independent loci (**Suppl. Table 34**). Five of these 109 loci were CC-GWAS-specific, implying they were not significantly associated with case-control status in either of the disorders in the respective disorder-pair. CC-GWAS also computes a genome-wide genetic distance between the cases of two disorders (F_ST,causal_), indicating how genetically dissimilar the cases are on average. As expected, these genetic distances were inversely correlated (*r* = -0.79, *SE* = 0.07) with *r_g_* (**Suppl. Table 35**). In support of the five-factor model, we find that >99% of the CC-GWAS hits were identified for disorder-pairs that loaded on separate factors (**Suppl. Tables 36-37**). This suggests that disorders that cluster on the same factor from the five-factor model are largely indistinguishable at the level of individual genetic variants.

### Functional Annotation Characterizes Pleiotropic and Disorder-Specific Signal

#### eQTL and Hi-C Datasets to Identify Target Genes

To understand the biological functions influenced by the risk loci identified with Genomic SEM, we prioritized candidate risk genes implicated by the multivariate GWAS loci using a combination of eQTL^44,45^ and Hi-C^44,46^ data sets collected from fetal and adult brain samples (**Method; Suppl. Tables 38-39**). Due to the limited number of variants associated with other factors, analyses were restricted to the transdiagnostic *p-*factor, the SB and Internalizing disorders factors, and Q_SNP_ for these latter two factors. We first compared the target gene expression along the temporal trajectory of human brain development, finding that genes associated with the three factors were expressed at higher levels than Q_SNP_ target genes across the lifespan, but with the largest difference observed at fetal stages and early life (**Fig. 5; Suppl. Fig. 17**). In line with prior cross-disorder findings,^5^ this suggests that pleiotropic variants are involved in early, fundamental neurodevelopmental processes. We went on to examine the biological processes regulated using gene ontology (GO) enrichment analysis.^12^ To enhance the specificity of the gene sets, we removed Internalizing and SB target genes that also appeared for the *p-*factor. The target genes of the *p-*factor were primarily enriched in broader biological processes related to gene regulation (**Fig. 5**). In contrast, SB (minus *p*-factor) target genes were enriched in more specific terms related to neuron development (no significant results were identified for Internalizing). Results from MAGMA^47^ (**Supplemental Method**) provided convergent support for the role of early neurodevelopmental processes in transdiagnostic psychiatric risk. Specifically, these results showed enrichment across the psychiatric factors, including the *p-*factor, with gene lists constructed from recent rare variant studies of neurodevelopmental disorders^48,49^ (**Suppl. Fig. 18**).

**Figure 5.**
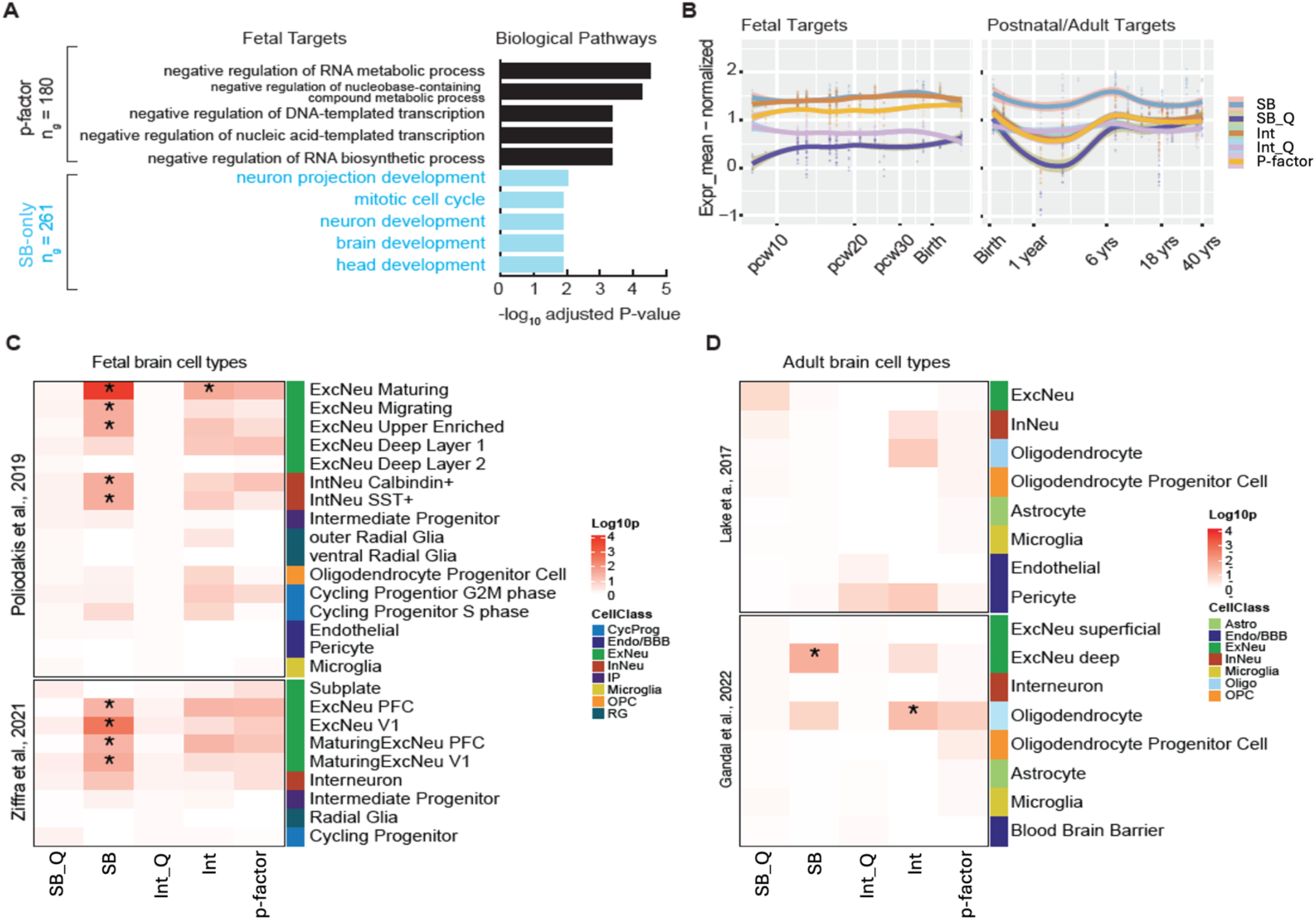
Functional genomics annotation of multi-diagnostic variants. (A) Gene ontology enrichment analysis of predicted target genes with transdiagnostic associations (i.e., variants associated with the *p-*factor), or those target genes associated with the SB factor that were not overlapping with *p-*factor target genes. (B) Averaged and normalized expression levels of target genes of the indicated classes along the temporal trajectory of human brain development. Shading around the lines reflect 95% confidence intervals. (C and D) Enrichment by expression of CDG3 variants target genes associated with the indicated factors in (C) fetal brain cell types using two independent scRNA-Seq data sets^50,51^ or (D) adult brain cell types using two independent snRNA-Seq data sets.^52,53^ SB_Q = SB factor QSNP; Int_Q = Internalizing disorders QSNP; Int = Internalizing disorders factor.

#### Expression Weighted Cell Type Enrichment (EWCE)

EWCE^11^ was estimated to determine whether genes for the *p-*factor, SB, and Internalizing factors were enriched within specific neuronal cell types in fetal and adult single-cell datasets.^50–53^ We found that genes associated with the SB factor during fetal development were significantly enriched within excitatory neuron subtypes across two datasets from fetal brain (**Fig. 5**) and in interneurons in one of the fetal datasets.^50^ SB genes also showed enrichment in deep-layer excitatory neurons derived from adult brain (**Fig. 5**). Internalizing disorder genes showed significant enrichment within excitatory maturing neurons and oligodendrocytes in the fetal and adult datasets, respectively (**Fig. 5**). We highlight, however, that a significant proportion of these genes are expressed during both fetal and adult stages, and that cell type enrichment is largely driven by genes that are not developmentally differentiated (**Suppl. Fig. 19**).

We also tested enrichment for CC-GWAS results to benchmark factor-level results against the disorder-specific signals. We specifically examined results for our two best powered disorders using the full set of MD or SCZ CC-GWAS hits, reflecting loci specific to these disorders. In contrast to the factor-level results, the only enrichments observed were with cycling progenitors in fetal brain for MD-specific genes and endothelial cell type in adult brain for SCZ-specific genes.

#### Stratified Genomic SEM

We applied Stratified Genomic SEM^10^ to characterize further the functional signal captured by the psychiatric factors in the five-factor and *p-*factor models. This involved estimating enrichment for 162 functional annotations that passed QC (**Method**; **Suppl. Table 40**). Functional annotations reflect sets of genetic variants grouped according to some property (e.g., heightened expression in a specific brain tissue). Enrichment of the factor variances in the five-factor model or for the *p-*factor reflects groups of genetic variants that index a disproportionate concentration of genetic risk sharing. For the *p-*factor model, we also examined the enrichment of the residual variances of the five lower-order factors. Annotations significant for a factor in both models are, therefore, likely to be specific to that factor. We employed a Bonferroni-corrected significance threshold of *p* < 2.81×10^-5^ (**Method**).

We identified 34 annotations significant for the SB factor in both models that are likely to be specifically relevant to the neurobiology of the SB factor. This included protein-truncating variant intolerant (PI) genes and the intersection between PI genes and several neuronal subtypes, including the intersection between PI genes and excitatory CA1 and CA3 hippocampal neuron annotations **(Fig. 6**; **Suppl. Table 40)**. 51 significant annotations were identified for the Internalizing disorders factor, including evolutionarily conserved and PI-oligodendrocyte precursor annotations (**Fig. 6**). We also found strong enrichment for an annotation reflecting neural progenitor biology,^54^ further implicating early neurobiological processes in shared psychiatric risk. No annotations, however, remained significant for the Internalizing factor’s residual variance (i.e., unique of the *p*-factor), as would be expected given that only 10% of the genetic variance in the Internalizing disorders factor was unique of *p*. Results revealed two significant annotations for the Compulsive factor (Fetal Female Brain H3K4me3, and dlPFC H3K27ac) across both models. There were five significant annotations for the Substance Use disorders factor in the five-factor model, including two evolutionarily conserved annotations that remained significant in the *p-*factor model. We identified no significant annotations for the Neurodevelopmental disorders factor. Finally, 64 significant annotations were detected for the *p*-factor, the strongest of which included the conserved primate, fetal male brain H3K4me1, and PI-GABAergic neuron annotations (**Suppl. Table 40**).

**Figure 6.**
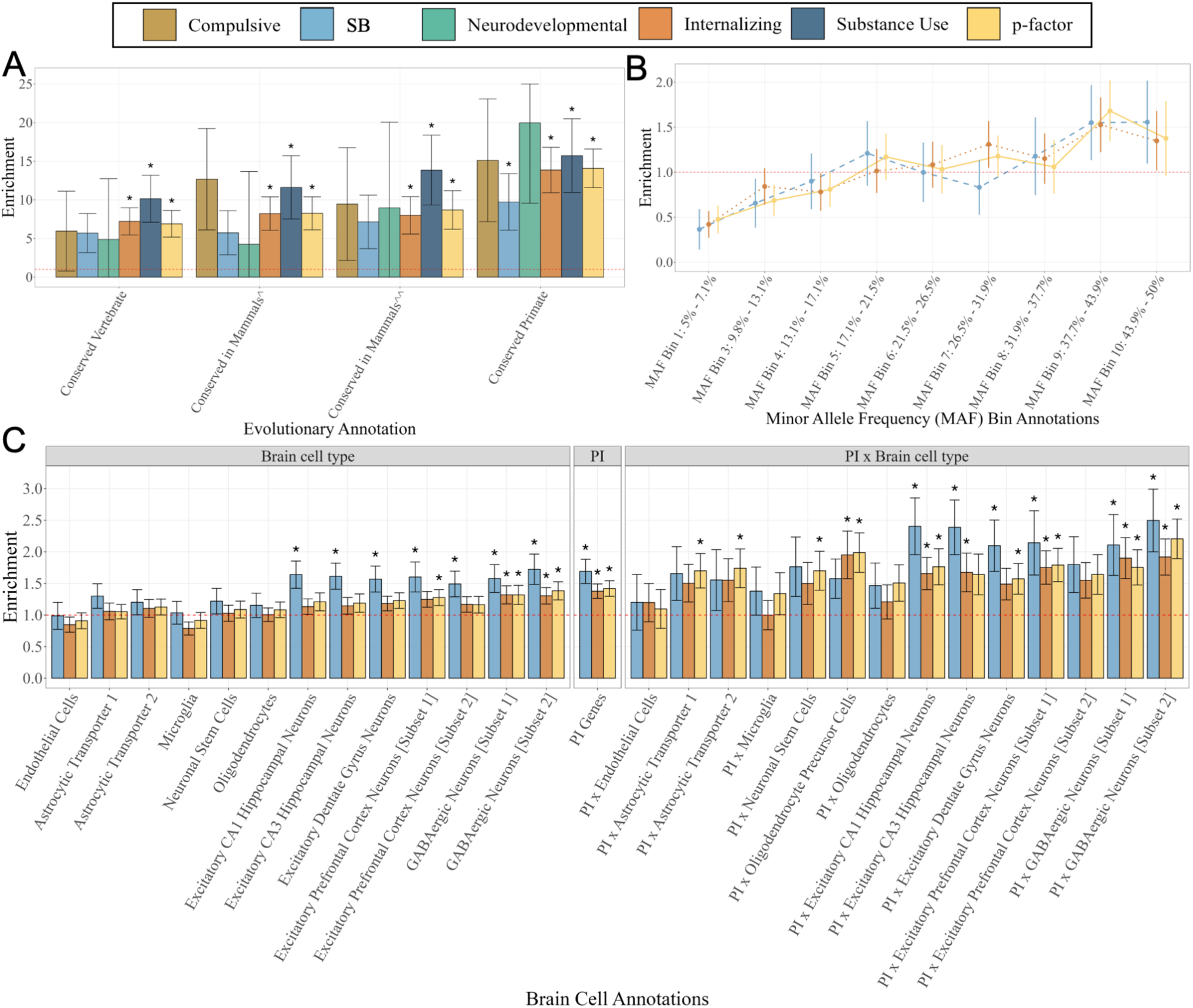
Stratified Genomic SEM results. *Panel A* depicts the enrichment point estimates for the four evolutionarily conserved annotations for the five-factor and *p-*factor models. The remaining two panels display only results for the SB, Internalizing, and *p*-factor due to the limited signal for the other factors. The two Conserved Mammals annotations reflect two sets of evolutionarily conserved genetic variants created by the original S-LDSC developers^61,62^ from data from Lindblad-Toh et al.^63^ (denoted with a ^) and genomic evolutionary rate profiling scores from Davydov et al.^64^ (denoted with ^^). *Panel B* displays patterns of enrichment across 9 minor allele frequency (MAF) bins (note that one of the 10 initial bins was removed following QC procedures described in the **Method**). Within the MAF range of 5% to 50%, these results reveal increasing enrichment across higher levels of MAF, suggesting that the genetic signal is most concentrated in the most common variants. *Panel C* shows the enrichment results for different brain cell types, protein-truncating variant intolerant (PI) genes, and the intersection across PI genes and brain cell types. All point estimates are depicted with 95% confidence intervals. Enrichment estimates that were significant at a strict Bonferroni significance threshold are shown with a *.

## Discussion

Our analyses characterized the complex genetic landscape of shared and differentiated risk for 14 psychiatric disorders. Several findings are notable across the applied methods. First, at a genome-wide level, we confirm pervasive genetic overlap across 14 clinically distinguished psychiatric disorders, as indicated by large pairwise *r*_g_ across multiple genetic ancestries and even greater overlap when considering divergent directional effects of shared loci. Second, this overlap is parsimoniously captured by five genomic factors (which we term the Compulsive, SB, Neurodevelopmental, Internalizing, and Substance Use disorders factors), that explain the majority of the genetic variance of the individual disorders (66% on average). Third, at a regional level, we identified 101 regions that harbor pleiotropic loci, including a hotspot on chromosome 11 with associations for 8 of the 14 disorders. Fourth, at the functional level we find that broadly pleiotropic variants are primarily involved in early neurobiological processes, while also identifying specific biological categories (e.g., different brain cell types) that uniquely confer risk to a more circumscribed subset of disorders. Finally, at an individual variant level, we identify 268 genetic loci associated with one of the five genomic factors, along with 412 loci that distinguish disorders that primarily belong to different factors.

Across multiple methods and levels of analysis, we find that the SB (defined by schizophrenia and bipolar disorder) and Internalizing (defined by major depression, PTSD, and anxiety) factors offer a particularly useful way to understand shared risk across these two sets of disorders. For these factors, a diverse set of methods produced convergent results across genome-wide (LDSC; MiXeR; Genomic SEM), regional (LAVA), and locus-level (multivariate GWAS; CC-GWAS) results, indicating that the disorders within these factors are characterized by highly overlapping genetic signal. At the functional level, we identified consistent results across different methods that highlighted the relevance of excitatory neurons, specifically hippocampal CA1 and CA3 neurons, for the SB factor and oligodendrocytes for the Internalizing disorders factor. Convergent evidence for shared signal across methods, along with functional results that characterize the specific biological substrata that underlie this shared signal, offers useful insight into how to conceptualize the shared etiology of these clusters of disorders.

At the genome-wide level, the *p-*factor statistically modeled the genetic overlap across the five factors and was found to be very strongly related to the Internalizing disorders factor, while the loadings for the remaining factors were more modest. This latter finding is consistent with the initial conceptualizations of *p* as reflecting a general tendency towards negative emotionality.^55^ On the one hand, in support of *p,* LAVA identified several pleiotropic hotspots characterized by widespread local *r*_g_ across many disorders, and multivariate GWAS yielded 160 hits for this factor alone. On the other hand, the *p-*factor also had more Q_SNP_ hits (117) than the five-factor model (33), indicating that the *p-*factor alone is insufficient for explaining cross-disorder risk. Consistent with the idea that the *p*-factor may reflect a more pluripotent level of variation, we observed enrichment of genetic signal for particularly broad biological categories, including gene regulation and evolutionarily conserved regions. By contrast, functional categories detected for the five factors implicated specific brain cell subtypes (e.g., excitatory neurons; oligodendrocytes). These results suggest a conceptual model in which there is a partial, broadly transdiagnostic component of genetic vulnerability to psychiatric disorders that largely captures internalizing genetic signals, with subsequent levels of more canalized and neurobiologically meaningful subdomains of psychopathology (as captured by the five factors).

Our study has several limitations. Analyses were restricted primarily to European genetic ancestry populations due to the limited availability of GWAS data for other genetic ancestry groups. Of note, cross-ancestry, within-trait *r*_g_ results were significantly <1, and the genetic correlation between MD and SCZ in EAS genetic ancestry individuals was double that observed in EUR participants (.45 vs .22). This divergence is not fully understood and highlights the importance of investing in genetic efforts for other ancestry groups. Results should be interpreted cautiously in light of recent evidence that cross-trait assortative mating may inflate *r_g_* estimates.^56^ However, it is unlikely that cross-trait assortment alone can explain the pattern of results for disorders with exceptionally high *r_g_*.^57^ Future research should also examine how genetic overlap across the psychiatric space varies across different environments (e.g., socioeconomic status; trauma exposure), for different comorbidity patterns observed within individuals, and build upon recent work that has looked at the psychiatric genetic architecture across biological sex.^58^

The current investigation into the genetic structure of psychopathology reflects the most comprehensive genomic examination of cross-disorder psychiatric risk to-date. By applying multiple cutting-edge methods at varying levels of biological granularity we produce an especially robust set of findings via triangulation^59^. Our findings identify subsets of disorders with especially high genetic overlap and characterize the biological processes implicated by this shared risk. Certain pharmacological interventions have proven to be effective across a range of disorders (e.g., selective serotonin reuptake inhibitors [SSRIs])^60^, indicating that future work could build on these findings to identify new or repurposed therapeutics for the genetic clusters of disorders identified here. While much remains to be done, cross-disorder genetics continues to fill in critical gaps in our understanding of shared and unique psychiatric risk with implications for constructing a more biologically valid psychiatric nosology.

## Online Methods

### Standardized Quality Control of Summary Statistics

A standard set of quality control (QC) filters was applied to all univariate GWAS summary statistics prior to conducting cross-disorder analyses. Any additional QC filters applied by a method are noted in its corresponding section below. These QC filters included removing strand ambiguous SNPs, restricting to SNPs with an imputation score (INFO) > 0.6 and with a minor allele frequency (MAF) > 1% when this information was available in the GWAS summary stats. Finally, we restrict to SNPs with a SNP-specific sum of the effective sample that is > 50% of the total sum of the effective sample or, when this SNP-specific information was not available, to SNPs for which > 50% of the cohorts contributed information, as indexed by the direction column in the GWAS summary stats. The MHC region was excluded from all summary statistics prior to the analysis. Base pair location is given in genome build GRCh37/hg19 throughout the manuscript and supplementary materials.

### Genomic SEM

#### Genome-wide models

All GWAS summary statistics were run through the *munge* function prior to running the multivariable version of LDSC used as input to Genomic SEM.^7^ The *munge* function aligns GWAS effects to the same reference allele and restricts to HapMap3 SNPs and SNPs with INFO > 0.9. LDSC was estimated using these “munged” summary statistics, applying a liability threshold model for all case-control psychiatric disorders (i.e., all disorders except for the NIC outcome, which reflects a GWAS of the continuous Fagerström Test for Nicotine Dependence [FTND]^27^). For comparability, population prevalence was chosen to match what was used in the corresponding manuscript for each trait. The ascertainment correction was performed using the sum of effective sample sizes across contributing cohorts for each disorder and setting the observed prevalence rate to .5 to reflect the fact that the effective sample size corresponds to the expected sample size under a balanced design.^65^ We note that for CUD^26^ we used the recently described formula^65^ for estimating the sum of effective sample size directly from the GWAS data. This is because in this instance we found the implied sum of effective sample size was much smaller than the value computed from the reported sample sizes, which is likely attributable to the complex familial structure in the deCODE sample used for those analyses.

The two primary estimates from multivariable LDSC are the genetic covariance matrix and the corresponding sampling covariance matrix. The genetic covariance matrix contains SNP-based heritabilities on the diagonal and the co-heritabilities (genetic covariances) across every pairwise combination of included disorders on the off-diagonal. The sampling covariance matrix contains squared standard errors (sampling variances) on the diagonal, which allows Genomic SEM to appropriately account for differences in the precision of GWAS estimates for disorders with unequal power. The off-diagonal contains sampling dependencies, which will arise in the presence of sample overlap across GWAS phenotypes. As these sampling dependencies are estimated directly from the data, summary statistics can be included with varying and unknown levels of sample overlap.

To guard against model overfitting, the EFA was performed on even chromosomes and used to inform the fitting of the CFA in odd chromosomes. The EFA was performed using the *factanal* R package for 2-5 factors using both promax (correlated) and varimax (orthogonal) rotations. Disorders were specified to load on a factor in the CFA when the standardized EFA loadings were > 0.3, with disorders allowed to cross-load (e.g., TS on the Compulsive and Neurodevelopmental factors) if this was the case for multiple factors. Models specified based on varimax EFA results still allowed for inter-factor correlations, as only allowing subsets of disorders to load on each factor will induce genetic overlap. A common factor model was also modeled to test a single latent factor model predicting all 14 disorders. We did not evaluate models with more than 5 factors as these caused issues with model convergence. Results revealed that a five-factor model specified based on the promax EFA results (**Suppl. Table 3**) fit the data best in odd chromosomes (CFI = .973, SRMR = .073; **Suppl. Table 2**). This model also fit the data well in all autosomes, and was subsequently carried forward for all analyses, along with the *p*-factor described in the main text. Considering the high *r*_g_ across PTSD and MD we also evaluated a model (in odd autosomes) that estimated the residual genetic covariance across these two disorders; however, we find that this does not significantly improve model fit relative to the model that does not include this parameter (model *X*^2^[1] difference = 2.86, *p* = .094).

#### Stratified Genomic SEM

Stratified Genomic SEM proceeds in two stages.^10^ In Stage 1, the *s_ldsc* function in Genomic SEM, a multivariable implementation of Stratified LDSC (S-LDSC),^61^ was used to estimate the stratified genetic covariance and sampling covariance matrices within each functional annotation. We specifically use the zero-order estimates for these analyses. In Stage 2, the *enrich* function was used to estimate the enrichment of the factor variances and residual genetic variances unique to the indicators. This is achieved by first estimating the model in the genome-wide annotation including all SNPs. The factor loadings from these genome-wide estimates are then fixed and the (residual) variances of the factors and disorders are freely estimated within each annotation. These reflect the within-annotation estimates for each variance component that are scaled to be comparable to the genome-wide estimates. This cumulative set of results is used to calculate the enrichment ratio of ratios. The numerator reflects the ratio of the within-annotation estimate over the genome-wide estimate. The denominator is the ratio of SNPs in the annotation over the total number of SNPs examined. Enrichment estimates greater than the null of 1 are therefore observed when an annotation explains a disproportionate level of genetic variance relative to the annotation’s size.

Functional annotations used to estimate the stratified matrices were obtained from a variety of data resources. This includes: (*i*) the baseline annotations from the 1000 Genomes Phase 3 BaslineLD v2.2^66^ from the S-LDSC developers;^61^ (*ii*) tissue specific gene expression annotation files created using data from GTEx^67^ and DEPICT^68^; (*iii*) tissue specific histone marks from the Roadmap Epigenetics project^69^; (*iv*) annotations we created^10^ from data in GTEx^67^ and the Genome Aggregate Database (gnomAD)^70^ that index protein-truncating variant (PTV)-intolerant (PI) genes, genes expressed in different types of brain cells in the human hippocampus and prefrontal cortex, and their intersection; (*v*) 11 neuronal cell type annotations defined by peaks from single-cell assay for transposase accessibility by sequencing (scATAC-seq) in the human forebrain;^51^ (*vi*) an annotation defined by peaks from transposase-accessible chromatin sequencing (ATAC-seq) data with greater accessibility in neural progenitor enriched regions encompassing the ventricular, subventricular, and intermediate zones (GZ) over neuron-enriched regions within the subplate, marginal zone and cortical plate (CP; GZ>CP), and a second CP>GZ annotation reflecting the converse;^54^ and (*vii*) a fetal and an adult annotations defined by expression quantitative trait loci (eQTLs) identified via high-throughput RNA-seq.^45^ We exclude 22 annotations that produce stratified genetic covariance matrices that were highly nonpositive definite to examine a total of 162 annotations. We correct for multiple testing employing a strict Bonferroni correction for the 162 annotations analyzed that passed quality control across the 11 factors examined (the factors from the five-factor factor model and the *p*-factor and residuals of the five factors from the *p-*factor model) of *p* < 2.81×10^-5^.

#### Multivariate GWAS

The *sumstats* function in Genomic SEM was used to align SNP effects across SNPs to the same reference allele and standardize the effects and their corresponding *SEs* relative to the total variance in the respective outcome. The *SE*s were additionally corrected for uncontrolled population stratification by taking the product of *SE*s and the LDSC univariate intercept when this value was > 1. After removing 136 SNPs that produced highly non-positive definite matrices when combined with the genetic covariance matrix, the final listwise deleted set consisted of 2,795,800 SNPs present across all 14 disorders. The *userGWAS* function was used to estimate the multivariate GWAS for SNP effects on the five factors from the five-factor model and the *p-*factor. We use significance threshold of *p* < 8.33×10^-9^, reflecting the standard genome-wide threshold of 5×10^-8^ with a Bonferroni correction for the six factors. As a quality control check we confirmed that the attenuation ratio^32^ was near 0 for all factors **(Suppl. Table 17),** indicating that the factor signal is not due to uncontrolled confounds (e.g., population stratification). This is to be expected given the noted LDSC intercept correction applied to the univariate traits prior to estimating multivariate GWAS.

The Q_SNP_ heterogeneity metric is a *X*^2^ distributed test statistic produced via a nested model comparison of a common pathway model, in which the SNP predicts a latent factor, to an independent pathways model, where the SNP directly predicts the factor indicators. Factor-specific Q_SNP_ estimates for the five-factor model were estimated using five independent pathways models that consisted of the SNP predicting both the indicators for one factor and the remaining four factors. For the *p-*factor model, the SNP predicted the five, first-order factors to obtain Q_SNP_ estimates for the second-order, *p-*factor.

### Popcorn Cross-Ancestry Analyses

We applied the cross-ancestry Popcorn^34^ method to estimate genetic impact correlation (ρ_*gi*_metric) across European (EUR), East Asian (EAS), and African/African American (AAM) ancestry groups. Six disorders were included in the analysis, including EAS summary statistics for MD and SCZ and AAM summary statistics for OUD, AUD, PTSD and CUD. The reference panel for the EAS dataset was based on 504 individuals from EAS population of the 1000 Genomes Phase3 data. For AAM, we performed the analysis using three alternative references from 1000G Phase3 data: (*i*) the African Ancestry in the southwest United States subgroup (*n* = 61); (*ii*) the African population (*n* = 661); and (*iii*) a reference panel created to capture the admixed ancestral background of some AAM individuals reflecting the combination across the EUR and AFR sample (*n* = 1,164). Cross-ancestry results and within-ancestry LDSC results for AAM and EAS populations are reported in **Suppl. Table 4.**

### MiXeR

MiXeR (v1.3) was applied following the procedure outlined in the original publication.^7^ We performed additional simulations to evaluate appropriate threshold for inclusion of a GWAS study in cross-trait MiXeR analysis. In prior simulations, we demonstrated that MiXeR cannot produce reliable estimates for analyses using low-powered input.^34^ More specifically, as statistical power increases, the AIC differences indicate that MiXeR-modelled estimates become increasingly more distinguishable from minimum and maximum overlap, corresponding to the increasing precision of MiXeR estimates. This demonstrates that AIC differences are sensitive to the input power of the summary statistics and can be used to support the reliability of MiXeR estimates. Based on these prior simulations, psychiatric disorders were brought forward for cross-trait MiXeR analysis when the product of *N_Eff_* and MiXeR ℎ^!^ estimates were > 12,000, where this cut point reflects the product of *N_Eff_* ≥ 100,000 and ℎ^!^ ≥ 0.12. As a result, we excluded OUD, TS, NIC, OCD, ASD, and CUD. Because AN was very close to this threshold and had a high AIC in univariate analysis, it was brought forward for cross-trait analyses along with the seven remaining psychiatric disorders. For the NIC summary statistics, we excluded two loci defined as a 2 Mb window around *CHRNA5* and *CHRNA4* genes known to have such a large effect on the phenotype that it would skew results. We note that for PTSD, ANX, and MD, the *r*_g_’s were so high that there was little room for additional overlap beyond correlation, given MiXeR’s modeling assumptions. More specifically, the range in size of the putative shared component is too small to allow for an accurate model fit in this situation, as demonstrated by the range on the respective x-axes (**Suppl. Fig. 7**). Also, there is a considerable uncertainty of polygenicity estimates for PTSD and ANX. Thus, cross-trait MiXeR results for PTSD, ANX, and MD should be interpreted with caution.

### LAVA

Local *r*_g_ analyses were conducted using LAVA v0.1.0.^8^ To avoid evaluating local *r*_g_’s in regions where there is a low amount of genetic signal (which could lead to unstable or uninterpretable estimates) for all phenotype pairs and loci separately, we used the univariate test in LAVA as a filtering step, computing bivariate local *r_g_*’s only in loci where both analyzed phenotypes have a ℎ^!^ significant at *p* < 4.6×10^-5^= .05 / 1,093 (where 1,093 represents the total number of analyzed loci). Given this filtering step, we performed 24,273 local *r*_g_ tests across all loci and phenotype pairs, resulting in a Bonferroni corrected *p*-value threshold of *p* < 2.1×10^-6^= .05 / 24,273 for the bivariate, local *r_g_* analyses.

Genomic loci used for the regional *r*_g_ analyses were defined by segmenting the genome into approximately equal-sized, semi-independent blocks using the LAVA partitioning algorithm (available at https://github.com/cadeleeuw/lava-partitioning). This algorithm works by iteratively splitting the chromosomes into smaller chunks, creating break points at regions where the LD between SNPs is the lowest (see the program manual for more details). To achieve a balance between block size and correlations between adjacent blocks, we ran the algorithm at the default parameters, changing only the minimum size requirement (in the number of SNPs) to 5,000, based on the 1,000 genomes data. Sample overlap was accounted for by obtaining the estimated intercepts from bivariate LDSC and providing these to LAVA.

### Case-Case GWAS (CC-GWAS)

CC-GWAS^9^ was applied to identify loci with different allele frequencies across cases of different disorders, contrasting cases one disorder-pair at a time. CC-GWAS is based on estimating a weighted difference of the case-control GWAS results of the disorders considered, thereby avoiding the necessity to match cases across disorders at individual level. CC-GWAS combines two components. The first component (CC-GWAS_OLS_) optimizes power and protects against type I error rate at null-null SNPs (SNPs that impact neither of both disorders), based on analytical expectations of genetic differences between cases and controls of both diseases. The second component (CC-GWAS_Exact_) controls type I error rate at ‘stress test’ SNPs (SNPs impacting both disorders resulting in no allele frequency difference across cases of both disorders). A SNP is significantly associated with case-case status when the *p*-value of the OLS component reaches genome-wide significance and when the *p*-value of the Exact-component is <10^-4^ (there is an upper bound on the number of stress test SNPs as these are causal SNPs). Importantly, CC-GWAS also filters false-positive associations that may arise due to (subtle) differential tagging of a stress test SNP in the respective case-control GWAS, which are present even in within-ancestry analysis.^9^ CC-GWAS excludes analyses of any disorder-pair with an *r*_g_ > 0.8 because these have a small genetic distance between cases with increased risk of type-I error at stress test SNPs.

### Locus Definition and Cross-Locus Overlap

The same locus definition (also referred to as a “hit” in the main text) was used for CC-GWAS and Genomic SEM. Significant loci were identified using the clumping functionality in PLINK v1.9 with an *r*^2^ threshold of 0.1 and a 3000 kb window. Physically proximal loci were additionally collapsed into a single locus when the locus windows were within 100 kb of one another. For the univariate results, we use the same locus definition applied to the complete set (i.e., without our QC filters) of GWAS summary statistics along with a genome-wide significance threshold of *p* < 5×10^-8^ without a Bonferroni correction. These more liberal QC and significance thresholds were utilized as univariate loci were strictly identified to benchmark whether Genomic SEM and CC-GWAS loci could be considered strictly novel. Loci across Genomic SEM, CC-GWAS, and univariate GWAS results were categorized as overlapping when the locus windows were within 100 kb of one another. The 1000 Genomes Phase 3 reference files were used for LD pruning for each respective genetic ancestry group (i.e., EUR, EAS, AAM).

### Functional Annotation of Genetic Variants

To predict the target genes of the variants (**Suppl. Fig. 17**), we first expanded the variants by including any variants within the linkage disequilibrium block (*r^2^* > 0.6) based on the EUR population using LDProxy from the *LDlink* R package.^71^ We began by curating the genes whose promoters (+/-500 bp from the TSS) or exons overlap with the variants of interest. Conversely, to map target genes that are not near the variants, we first filtered the variants for those localized in either human fetal brain open chromatin regions^54^ or human adult brain H3K27ac ChIP-Seq regions,^44^ both of which indicate enhancer activity, but during different stages of brain development. Next, we assigned target genes to each filtered variant using eQTL^44,45^ or HiC loops^44,46^ generated from samples from the corresponding stages. We also assigned variants present in promoter or exonic regions to the corresponding genes (**Suppl. Fig. 17**). Finally, we filter all the target genes for those expressed (RNA-Seq count > 0) in the corresponding tissues. In this way, we obtained 715 and 572 target genes in fetal and adult brains, respectively (**Suppl. Tables 40-41**). Notably, there is a prominent overlap between the two sets of genes, which is a result of the shared, positional mapping of genes to promoters or exons (**Suppl. Fig. 17**). Both the fetal and adult target genes were enriched in gene ontology (GO) terms related to neuron or brain development, suggesting the biological relevance of the genetic variants.

To plot the temporal expression trends of the predicted target genes, we used gene expression data sets from the BrainSpan (https://brainspan.org/static/download.html). We plotted the averaged gene expression (RPKM) of the selected genes over all samples collected from the cortex at the available stages of development, then generated a smoothened curve with the ‘loess’ method. We performed gene ontology (GO) enrichment analysis using the ToppGene suite.^36^ We filtered the enriched terms by containing at least 10% of the input list of genes, then displayed up to top 5 terms by adjusted p-values under the indicated category.

Finally, Expression Weighted Cell Type Enrichment (EWCE; https://nathanskene.github.io/EWCE/)^11^ was used to assess the cell type enrichment of target genes for the variants using a size-biased averaging method. This method uses single cell datasets to compute the average expression of a set of genes (in this case, genes assigned to variants for each factor) compared to a background set of genes, which is the set of expressed genes in the single cell dataset by randomly sampling sets from the background this 100,000 times. Annotations were taken from publicly available datasets^50–53^ but simplified to provide cell-type level instead of cluster level enrichments. For example, several upper-layer clusters in the Gandal et al. (2022)^53^ dataset were combined into “ExcNeu superficial”, etc. *P*-values were FDR corrected based on the number of cell types × gene-lists.

## Data Availability

The data that support the findings of this study are all publicly available or can be requested for access. Specific download links for various datasets are directly below.

Psychiatric disorder GWAS summary statistics for data from the PGC can be downloaded or requested here: https://www.med.unc.edu/pgc/download-results/

Links to the LD-scores and reference panel data for GenomicSEM analyses can be found here: https://github.com/GenomicSEM/GenomicSEM/wiki

Links to the BaselineLD v2.2 annotations can be found here: https://data.broadinstitute.org/alkesgroup/LDSCORE

GWAS summary statistics for alcohol use disorder, opioid use disorder, and anxiety disorder included restricted access data from the Million Veterans Program. Access to this data can be requested through dbGAP. We also make available multivariate GWAS summary statistics for the latent psychiatric factors in GenomicSEM using only public access data. This data will be deposited in the GWAScatalog at the time of publication in a peer-reviewed journal outlet.

## Code Availability

Genomic SEM analyses were implemented using the publicly available code here for v0.5.0: https://github.com/GenomicSEM/GenomicSEM

CC-GWAS was conducted using publicly available code here for v0.1.0: https://github.com/wouterpeyrot/CCGWAS

MiXeR was conducted using publicly available code here for v1.3: https://github.com/precimed/mixer

LAVA was conducted using publicly available code here for v.0.1.0: https://github.com/josefin-werme/LAVA

## Acknowledgements

We acknowledge the work of the individual Psychiatric Genomics Consortium working groups, the iNDiGO consortium, and the Million Veterans Program that contributed summary statistics to these analyses. This work was made possible by the contributions of the many investigators that comprise these working groups and the numerous grants from governmental and charitable bodies as well as philanthropic donation. We acknowledge the Mayo Clinic Biobank (MCB) research team, as well as the patient-participants who consented to participate in this research program. We also acknowledge the Mayo Clinic Center for Individualized Medicine for support of the Mayo Clinic Biobank, and Regeneron Genetics Center for providing genetic data for MCB participants for the analysis. In particular, we want to thank the research participants worldwide who shared their life experiences and biological samples to make work like this possible. The PGC has been supported by the following grants: MH085508, MH085513, MH085518, MH085520, MH094411, MH094421, MH094432, MH096296, MH109499, MH109501, MH109514, MH109528, MH109532, MH109536, MH109539, MH124871, MH124851, MH124839, MH124847, MH124873, MH124875, DA054869. Specific investigators have been supported by the following grants: A.D.G.: R01MH120219, R01AG073593; J.W.: European Union Horizon 2020 grant agreement 964874 (RealMent); Q.G.: Autism Speaks Postdoctoral Fellowship; S.S.R.: DP1DA054394, T32IR5226; C.L.: ERC-2018-ADG 834057; S.V.F.:European Union Horizon grant agreement 965381, U01AR076092, R01MH116037, 1R01NS128535, R01MH131685, 1R01MH130899, U01MH135970 and Supernus; M.P.M.: F30MH135712; S.N.B.: Fapesp 2014/50917-0 - CNPq 465550/2014-2; T.T.M.: K08MH135343; A.D.B.: Lundbeck Foundation (R102-A9118, R155-2014-1724, and R248-2017-2003), NIH/NIMH (1R01MH124851-01), and EU’s Horizon Europe program under grant agreement no. 101057385 (R2D2-MH); K.S.O.: R01MH124839-02, Research Council of Norway (RCN) #334920; S.L.G.: U54GM115516; J.M.H.: R01MH124847; H.J.E.: R01DA054869; A.X.M.: R01MH106595; K.C.C., C.M.N., and M.B.S.: R01MH106595; C.M.N.:R01MH124847; J.A.K.: R01MH112904, R01MH123775, U24MH068457, R01MH104964, R01MH123451; P.H.L.: R01MH119243, R01GM148494; E.M.T.: R01MH120219, R01AG073593, P30AG066614, P2CHD042849; B.J.C., A.B., V.P., and J.M.B.: R01MH121924; M.J.G.: R01MH123922, R01MH121521; B.F.: R01MH124851; L.M.T.: R01MH136149, R01120170; K.G.J.: R21MH123908, K08MH122673.; D.D.: The Novo Nordisk Foundation (NNF20OC0065561, NNF21SA0072102), the Lundbeck Foundation (R344-2020-1060), the European Union’s Horizon 2020 research and innovation program under grant agreement No. 965381(TIMESPAN).

## Author Contributions

A.D.G., J.W., W.J.P., and O.F. conducted the primary analyses presented in the paper for GenomicSEM, LAVA, CC-GWAS, and MiXeR, respectively.

L.K.B., Q.G., and M.P.M. ran the functional follow-up analyses.

B.J.C., A.B., V.P., and J.M.B. conducted the PheWAS analyses.

C.L., E.M.T., and P.H.L. provided additional feedback on the analyses and included data.

A.D.G., J.W., W.J.P., O.F., K.S., and J.W.S. wrote the initial draft of the manuscript.

K .S. and J.W.S. jointly supervised the research.

All named authors provided iterative feedback on the manuscript.

All collaborators within the listed working group banners approved the contents of the manuscript.

## Additional information

### Supplementary information

The online version contains supplementary tables and an online supplement.

**Correspondence and requests for materials** should be addressed to Andrew D. Grotzinger and Jordan W. Smoller.

^†^*Working group members and affiliations. Note that the list of working group members is organized alphabetically by last name. We also highlight that these lists are not exhaustive with respect to the members of the working groups; rather, they reflect individual members of the working groups that approved the contents of this manuscript*.

### Anxiety Disorders Working Group

Daniel E Adkins^1,2^, Georg W Alpers^3^, Helga Ask^4,5^, Sintia I Belangero^6,7^, Ottar Bjerkeset^8,9^, Sigrid Børte^10,11,12,13,14^, Gerome Breen^15,16,17^, Sandra A Brown^18,19^, Enrique Castelao^20^, Hilary Coon^21,22^, William E Copeland^23^, Elizabeth C Corfield^24,5^, Darina Czamara^25^, Jürgen Deckert^26^, Anna R Docherty^21,27,28,29^, Katharina Domschke^30^, Ole Kristian Drange^31,32,33,8^, Thalia C Eley^17^, Angelika Erhardt-Lehmann^26,34^, Andreas J Forstner^35,36,37^, Miguel Garcia-Argibay^38,39^, Scott D Gordon^40,41^, Ian B Hickie^42^, Iiris Hovatta^43,44^, Matthew H Iveson^45^, James L Kennedy^46,47,48,49^, Henrik Larsson^38,39^, Daniel F Levey^50,51^, Christine Lochner^52^, Michelle K Lupton^53,54^, Hermine HM Maes, Eduard Maron^55,56^, Nicholas G Martin^40,41,57^, Manuel Mattheisen^26,58,59,60,61,62,63^, Sandra M Meier^64^, Christiane A Melzig^65^, Brittany L Mitchell^41^, Teemu Palviainen^66^, Roseann E Peterson^67^, Giorgio Pistis^20^, Martin Preisig^20^, Börge Schmidt^68^, Johannes Schumacher^35^, Andrey A Shabalin^21,28^, Anne Heidi Skogholt^11^, Dan J Stein^69^, Murray B Stein^18,70^, Eystein Stordal^71,72,73,8^, Andreas Ströhle^74^, Nora I Strom^61,75,76,77^, Elisa Tasanko, Laurent Thomas^11,78,79,80^, Henning Tiemeier^81,82^, Heike Weber^26^, Bendik S Winsvold^10,11,13,83^, Clement C Zai^47,84,85^, Gwyneth Zai^47,86^, John-Anker Zwart^10,11,12,13^

### Attention-Deficit/Hyperactivity Disorder (ADHD) Working Group

Silvia Alemany^87,88,89,90^, Claiton HD Bau^91,92^, Sintia I Belangero^6,7^, Dorret I Boomsma^93,94^, Rosa Bosch^87,90,95,96,97,98^, Isabell Brikell^38,76,99^, Christie L Burton^100^, Miquel Casas^101,102,87,90,96,97,98^, Elizabeth C Corfield^24,5^, Bru Cormand, Jennifer Crosbie^100,47^, Alysa E Doyle^103,104^, Josephine Elia, Stephen V Faraone^105^, Barbara Franke^106,107,108^, Miguel Garcia-Argibay^38,39^, Joseph T Glessner^109,110^, Eugenio H Grevet^111,91^, Jan Haavik^112,113^, Alexandra Havdahl^5^, Ziarih Hawi^114^, Anke Hinney^115,116^, Daniel P Howrigan^117,84^, Marieke Klein^106,108^, Henry R Kranzler^118,119,120^, Jonna Kuntsi^17^, Kate Langley^121^, Henrik Larsson^38,39^, Klaus-Peter Lesch^122,123^, Calwing Liao^117,124,125,84^, Sandra K Loo^126^, Hermine HM Maes, James J McGough^126,127^, Sarah E Medland^128,129,40,41^, Nina R Mota^106,108^, Benjamin M Neale^117,130,84^, Michael C O’Donovan^131,132^, Roseann E Peterson^67^, Danielle Posthuma^133,134,135,136^, Josep Antoni Ramos-Quiroga^137,138,87,90,95,96,97^, Andreas Reif^139,140,141^, Marta Ribasés^87,88,89,90,97^, Diego L Rovaris^142,143^, Russell J Schachar^100,47^, Stephen W Scherer^144,145^, Yingjie Shi^106,108^, María Soler Artigas^87,88,89,90,97^, Edmund js Sonuga-Barke^146,147,148^, Hans-Christoph Steinhausen^149^, Ludger Tebartz van Elst^30^, Martin Tesli^150^, Raymond K Walters^117,84^, Stephanie H Witt^151,152^, Yanli Zhang-James^153^

### Autism Spectrum Disorders Working Group

Edwin H Cook^154^, Elizabeth C Corfield^24,5^, Jakob Grove^155,156,58,62,63,76^, Alexandra Havdahl^5^, Susan S Kuo^103,157^, Joseph Piven^158,159^, Danielle Posthuma^133,134,135,136^, Elise B Robinson^103,84^, Stephan J Sanders^160,161,162^, Stephen W Scherer^144,145^, Ludger Tebartz van Elst^30^, Mohammed Uddin^163,164^, Jacob AS Vorstman^144,165^, Varun Warrier^166^, Lauren A Weiss^167^

### Bipolar Disorder Working Group

Rolf Adolfsson^168^, Kristina Adorjan^169,61^, Ingrid Agartz^170,171,59^, Esben Agerbo^172,63^, Mariam Al Eissa^173^, Diego Albani^174^, Martin Alda^175,64^, Silvia Alemany^87,88,89,90^, Lars Alfredsson^176^, Ney Alliey-Rodriguez^177,178^, Thomas D Als^58,62,63^, Till F M Andlauer^179^, Ole A Andreassen^171,180^, Adebayo Anjorin^181^, Verneri Antilla^117^, Anastasia Antoniou^182^, Swapnil Awasthi^183^, Lena Backlund^184,185^, Ji Hyun Baek^186^, Marie Bækvad-Hansen^187,63^, Nicholas Bass^173^, Anthony J Batzler^188^, Michael Bauer^189^, Bernhard T Baune^190,191,192,193^, Eva C Beins^36^, Frank Bellivier^194,195^, Susanne Bengesser^196^, Sarah E Bergen^38^, Wade H Berrettini^197^, Joanna M Biernacka^188,198,199^, Tim B Bigdeli^200,201,202,203^, Armin Birner^196^, Douglas H R Blackwood^204^, Carsten Bøcker Pedersen^172,63^, Michael Boehnke^205^, Erlend Bøen^170^, Marco P Boks^206^, Anders D Børglum^156,207,63^, Sigrid Børte^10,11,12,13,14^, Rosa Bosch^87,90,95,96,97,98^, Gerome Breen^15,16,17^, Murielle Brum^139^, Ben M Brumpton^13^, Nathalie Brunkhorst-Kanaan^139^, Julien Bryois^38^, Monika Budde^61^, Jonas Bybjerg-Grauholm^187,208,63^, William Byerley^209^, Murray Cairns^210^, Vaughan J Carr^211^, Miquel Casas^101,102,87,90,96,97,98^, Stanley Catts^212^, Pablo Cervantes^213^, Alexander W Charney^214^, Sven Cichon^215,216,36,37^, Toni-Kim Clarke^204^, Jonathan RI Coleman^15,16,17^, Brandon J Coombes^188,198^, Aiden Corvin^217^, Nicholas Craddock^132^, Cristiana Cruceanu^213,218^, Alfredo B Cuellar-Barboza^199,219^, Julie Cunningham^220^, David Curtis^221,222^, Piotr M Czerski^223^, Anders M Dale^224^, Nina Dalkner^196^, Udo Dannlowski^192,225^, Friederike S David^226,36^, Franziska Degenhardt^116,36^, J Raymond DePaulo^227^, Arianna Di Florio^132,228^, Dimitris Dikeos^229^, Srdjan Djurovic^230,231^, Amanda Dobbyn^214,232^, Athanassios Douzenis^182^, Ole Kristian Drange^31,32,33,8^, Howard J Edenberg^233,234^, Torbjørn Elvsåshagen^12,171,83^, Valentina Escott-Price^132^, Tõnu Esko^130,235,236,237^, Bruno Etain^194,195^, Panagiotis Ferentinos^17,182^, I Nicol Ferrier^238^, Alessia Fiorentino^173^, Tatiana M Foroud^234^, Andreas J Forstner^35,36,37^, Liz Forty^132^, Josef Frank^152^, Oleksandr Frei^12,180^, Nelson B Freimer^239,240^, Louise Frisén^59^, Mark Frye^199^, Janice M Fullerton^241,242,243,244^, Katrin Gade^245,61^, Michael J Gandal^240^, Julie Garnham^64^, Micha Gawlik^26^, Joel Gelernter^246,247,51^, Elliot S Gershon^177,248^, Marianne Giørtz Pedersen^172,63^, Ian R Gizer^249^, Fernando S Goes^227^, Scott D Gordon^40,41^, Katherine Gordon-Smith^250^, Melissa J Green^211,242^, Tiffany A Greenwood^18^, Maria Grigoroiu-Serbanescu^251^, Jakob Grove^155,156,58,62,63,76^, José Guzman-Parra^252^, Kyooseob Ha^253^, Saskia P Hagenaars^16,17^, Magnus Haraldsson^254^, Joanna Hauser^223^, Martin Hautzinger^255^, Urs Heilbronner^61^, Dennis Hellgren^38^, Frans Henskens^210^, Stefan Herms^215,216,36^, Jan Hillert^59^, Per Hoffmann^215,216,36^, Peter A Holmans^132^, Kyung Sue Hong^186^, David M Hougaard^187,208,63^, Laura Huckins^214,232^, Christina M Hultman^38^, HUNT All-In Psychiatry, Kristian Hveem^11,13^, Masashi Ikeda^256^, Nakao Iwata^256^, Assen V Jablensky^257^, Stéphane Jamain^258,259^, James A Knowles^260,261,262^, Jessica S Johnson^214,232^, Ian Jones^132^, Lisa A Jones^250^, René S Kahn^206,214^, Janos L Kalman^169,263,61^, Yoichiro Kamatani^264,265^, Nolan Kamitaki^235,84^, John R Kelsoe^18^, James L Kennedy^46,47,48,49^, Minsoo Kim^240^, George Kirov^132^, Sarah Kittel-Schneider^139,26^, Manolis Kogevinas^266^, Maria Koromina^267^, Thorsten M Kranz^139^, Henry R Kranzler^118,119,120^, Kristi Krebs^237^, Michiaki Kubo^268^, Ralph Kupka^269,270,271^, Steven A Kushner^272^, Mikael Landén^121,38^, Catharina Lavebratt^184,185^, Jacob Lawrence^273^, Markus Leber^274^, Marion Leboyer^258,259,275^, Heon-Jeong Lee^276^, Phil H Lee^277^, Shawn E Levy^278^, Cathryn M Lewis^16,17,279^, Catrin Lewis^132^, Qingqin S Li^280,281^, Calwing Liao^117,124,125,84^, Penelope A Lind^41,54^, Jolanta Lissowska^282^, Christine Lochner^52^, Carmel Loughland^210^, Susanne Lucae^218^, Martin Lundberg^184,185^, Jurjen J Luykx^123,283,284^, Donald J MacIntyre^285^, Sigurdur H Magnusson^286^, Wolfgang Maier^287^, Adam Maihofer^18^, Dolores Malaspina^214,232^, Mirko Manchia^288,289^, Eirini Maratou^290^, Nicholas G Martin^40,41,57^, Lina Martinsson^59^, Carol A Mathews^291,292^, Manuel Mattheisen^26,58,59,60,61,62,63^, Morten Mattingsdal^293,33^, Fermin Mayoral^252^, Steven A McCarroll^235,84^, Susan L McElroy^294^, Nathaniel W McGregor^295^, Peter McGuffin^17^, Andrew M McIntosh^285,296^, James D McKay^297^, Francis J McMahon^298^, Andrew McQuillin^173^, Helena Medeiros^262^, Sarah E Medland^128,129,40,41^, Sandra M Meier^64^, Ingrid Melle^180,299^, Patricia Michie^210^, Lili Milani^237^, Vincent Millischer^184,185^, Brittany L Mitchell^41^, Philip B Mitchell^211,300^, Grant W Montgomery^301^, Jennifer L Moran^302,84^, Gunnar Morken^303,8^, Derek W Morris^304^, Ole Mors^305,63^, Preben Bo Mortensen^172,62,63^, Bryan Mowry^212^, Thomas W Mühleisen^215,37^, Bertram Müller-Myhsok^218,306,307^, Niamh Mullins^214,232^, Richard M Myers^278^, Woojae Myung^253,308^, Benjamin M Neale^117,130,84^, Caroline M Nievergelt^18,309^, Merete Nordentoft^310,311,63^, Markus M Nöthen^36^, John I Nurnberger^312^, John I Nurnberger Jr^313^, Niamh O’Brien^173^, Kevin S O’Connell^171,180^, Claire O’Donovan^64^, Michael C O’Donovan^131,132^, Ketil J Oedegaard^113,314^, Loes M Olde Loohuis^239,240^, Tomas Olsson^315^, Roel A Ophoff^239,240,272,316^, Lilijana Oruc^317^, Michael J Owen^131,132^, Sara A Paciga^318^, Georgia Panagiotaropoulou^183^, Chris Pantelis^319^, Sergi Papiol^169,61^, Antonio F Pardiñas^132^, Carlos Pato^262^, Michele T Pato^262^, George P Patrinos^267,320,321,322,323,324^, Roy H Perlis^325,326^, Amy Perry^250^, Andrea Pfennig^189^, Evgenia Porichi^182^, Danielle Posthuma^133,134,135,136^, James B Potash^227^, Zhen Qiao^301^, Digby Quested^327,328^, Towfique Raj^232,329,330,331^, Josep Antoni Ramos-Quiroga^137,138,87,90,95,96,97^, Mark H Rapaport^332^, Eline J Regeer^333^, Andreas Reif^139,140,141^, Eva Z Reininghaus^196^, Marta Ribasés^87,88,89,90,97^, John P Rice^334^, Marcella Rietschel^152^, Stephan Ripke^117,183,84^, Fabio Rivas^252^, Margarita Rivera^335,336^, Julian Roth^26^, Guy A Rouleau^125,337^, Panos Roussos^214,232,329^, Douglas M Ruderfer^338^, Takeo Saito^256^, Cristina Sánchez-Mora^87,88,90,97^, Ulrich Schall^210^, Martin Schalling^184,185^, Brian M Schilder^232,329,330,331^, Peter R Schofield^242,243^, Eva C Schulte^169,339,36,61^, Thomas G Schulze^152,153,227,245,61^, Laura J Scott^205^, Rodney J Scott^210^, Fanny Senner^169,61^, Alessandro Serretti^340,341^, Cynthia Shannon Weickert^211,242,342^, Sally Sharp^173^, Paul D Shilling^18^, Engilbert Sigurdsson^254,343^, Lea Sirignano^152^, Claire Slaney^64^, Laura G Sloofman^232^, Olav B Smeland^171,180^, Daniel J Smith^285^, Jordan W Smoller^277,302,84^, Janet L Sobell^344^, Christine Søholm Hansen^187,63^, María Soler Artigas^87,88,89,90,97^, Anne T Spijker^345^, Eli A Stahl^130,214,232^, Hreinn Stefansson^286^, Kari Stefansson^286,346^, Dan J Stein^69^, Stacy Steinberg^286^, Eystein Stordal^71,72,73,8^, John S Strauss^46^, Fabian Streit^152^, Jana Strohmaier^152^, Patrick F Sullivan^228,347,38^, Beata Świątkowska^348^, Chikashi Terao^264^, Martin Tesli^150^, Thorgeir E Thorgeirsson^286^, Claudio Toma^242,243,349^, Paul Tooney^210^, Vassily Trubetskoy^183^, Evangelia-Eirini Tsermpini^267^, Gustavo Turecki^350^, Arne E Vaaler^351^, Marquis P Vawter^352^, Helmut Vedder^353^, Eduard Vieta^354^, John B Vincent^355,46^, Irwin D Waldman^356^, James T R Walters^132^, Thomas W Weickert^211,242,342^, Thomas Werge^310,357,358,63^, Bendik S Winsvold^10,11,13,83^, Stephanie H Witt^151,152^, Hong-Hee Won^359^, Naomi R Wray^301,327,360^, Simon Xi^361^, Wei Xu^362^, Jessica Mei Kay Yang^132^, Allan H Young^363,364^, Hannah Young^232^, Peter P Zandi^227^, Hang Zhou^246,365,51^, Lea Zillich^152^, John-Anker Zwart^10,11,12,13^

### Eating Disorders Working Group

Roger A Adan^366,367^, Lars Alfredsson^176^, Helga Ask^4,5^, Andreas Birgegård^38^, Gerome Breen^15,16,17^, Cynthia M Bulik^228,368,38^, Jonathan RI Coleman^15,16,17^, Christian Dina^369^, Monika Dmitrzak-Weglarz^370^, Elisa Docampo^371,372^, Karin M Egberts^122^, Fernando Fernandez-Aranda^373,374,375,376^, Katrin E Giel^377,378,379^, Scott D Gordon^40,41^, Philip Gorwood^380^, Alexandra Havdahl^5^, Anke Hinney^115,116^, Christopher Hübel^17,172^, James I Hudson^381^, Susana Jimenez-Murcia^373,374,375,376^, Jennifer Jordan^382,383^, Gursharan K Kalsi^15,17^, Jaakko Kaprio^66^, Leila Karhunen^384^, Martien JH Kas^366,385^, James L Kennedy^46,47,48,49^, Martin A Kennedy^386^, Mikael Landén^121,38^, Janne T Larsen^172^, Qingqin S Li^280,281^, Lisa R Lilenfeld^387^, Jurjen J Luykx^123,283,284^, Nicholas G Martin^40,41,57^, Sarah E Medland^128,129,40,41^, Alessio Maria Monteleone^388^, Melissa A Munn-Chernoff^389^, Benedetta Nacmias^390,391^, Roel A Ophoff^239,240,272,316^, Liselotte V Petersen^172^, Dalila Pinto^392^, Anu Raevuori^393,394^, Nicolas Ramoz^380^, Valdo Ricca^395^, Marion E Roberts^396^, Filip Rybakowski^397^, Ulrike H Schmidt^398^, Alexandra Schosser^399,400^, Sandro Sorbi^390,391^, Michael A Strober^240^, Laura M Thornton^401^, Hunna J Watson^401,402,403^, Stephan Zipfel^404,405,406^

### Major Depressive Disorder Working Group

Till F M Andlauer^179^, Helga Ask^4,5^, Bernhard T Baune^190,191,192,193^, Sintia I Belangero^6,7^, Michael E Benros^310,407^, Tim B Bigdeli^200,201,202,203^, Ottar Bjerkeset^8,9^, Dorret I Boomsma^93,94^, Gerome Breen^15,16,17^, Rodrigo A Bressan^408^, Enda M Byrne^409^, Carolina M Carvalho^410,7^, Enrique Castelao^20^, Boris Chaumette^350,380^, Sven Cichon^215,216,36,37^, Jonathan RI Coleman^15,16,17^, Lucia Colodro-Conde^129^, Hilary Coon^21,22^, William E Copeland^23^, Elizabeth C Corfield^24,5^, Darina Czamara^25^, Udo Dannlowski^192,225^, Eske M Derks^41^, Anna R Docherty^21,27,28,29^, Katharina Domschke^30^, Erin C Dunn^103,104^, Chiara Fabbri^340^, Giuseppe Fanelli^106,340^, Jerome C Foo^152^, Andreas J Forstner^35,36,37^, Josef Frank^152^, Ary Gadelha^408^, Zachary F Gerring^41,54^, Fernando S Goes^227^, Scott D Gordon^40,41^, Hans J Grabe^411^, Jakob Grove^155,156,58,62,63,76^, Jan Haavik^112,113^, Andrew C Heath^334^, Matthew H Iveson^45^, James A Knowles^260,261,262^, Jaakko Kaprio^66^, James L Kennedy^46,47,48,49^, Henry R Kranzler^118,119,120^, Kristi Krebs^237^, Mikael Landén^121,38^, Kelli Lehto^237^, Daniel F Levey^50,51^, Douglas F Levinson^412^, Glyn Lewis^173^, Qingqin S Li^280,281^, Penelope A Lind^41,54^, Jurjen J Luykx^123,283,284^, Hermine HM Maes, Eduard Maron^55,56^, Nicholas G Martin^40,41,57^, Sarah E Medland^128,129,40,41^, Lili Milani^237^, Brittany L Mitchell^41^, Woojae Myung^253,308^, Michael C O’Donovan^131,132^, Vanessa K Ota^6,7^, Michael J Owen^131,132^, Teemu Palviainen^66^, Pedro M Pan^7^, Peristera Paschou^413^, Roseann E Peterson^67^, Giorgio Pistis^20^, Danielle Posthuma^133,134,135,136^, James B Potash^227^, Martin Preisig^20^, Andreas Reif^139,140,141^, John P Rice^334^, Marcella Rietschel^152^, Brien P Riley^414^, Giovanni A Salum^415,416,417,418^, Marcos L Santoro^419,7^, Eva C Schulte^169,339,36,61^, Alessandro Serretti^340,341^, Andrey A Shabalin^21,28^, Lea Sirignano^152^, Dan J Stein^69^, Murray B Stein^18,70^, Fabian Streit^152^, Jana Strohmaier^152^, Martin Tesli^150^, Jackson G Thorp^41,54^, Henning Tiemeier^81,82^, Sandra Van der Auwera^411^, Bradley T Webb^29,420^, Stephanie H Witt^151,152^, Naomi R Wray^301,327,360^

### Nicotine Dependence GenOmics (iNDiGO) Consortium

Timothy B Baker^421^, Dorret I Boomsma^93,94^, Danielle M Dick^422,423^, Dmitriy Drichel^424^, Lindsay A Farrer^425^, Nathan C Gaddis^420^, Dana B Hancock^426^, John E Hokanson^427^, Jouke-Jan Hottenga^93^, Eric O Johnson^420,428^, Jaakko Kaprio^66^, Henry R Kranzler^118,119,120^, Pamela A Madden^334^, Mary L Marazita^429^, Jesse A Marks^420^, Daniel W McNeil^430^, Michael Nothnagel^431^, Teemu Palviainen^66^, Bryan C Quach^420^, Marcella Rietschel^152^, Nancy L Saccone^432,433^, Nancy YA Sey^434^, Richard Sherva^425,435^, Scott Vrieze^436^, Alex Waldrop^437^, Georg Winterer^438^, Kendra Young^427^, Stephanie Zellers^436,66^

### Obsessive-Compulsive Disorder Working Group

Silvia Alemany^87,88,89,90^, Helga Ask^4,5^, Csaba Barta^439^, Katharina Bey^287^, Oscar J Bienvenu^227^, Julia Boberg^75^, Rosa Bosch^87,90,95,96,97,98^, Christie L Burton^100^, Jonas Bybjerg-Grauholm^187,208,63^, Enda M Byrne^409^, Adrian Camarena^440^, Beatriz Camarena^441^, Miquel Casas^101,102,87,90,96,97,98^, Danielle C Cath^442,443^, Edwin H Cook^154^, Jennifer Crosbie^100,47^, James J Crowley^347^, Eske M Derks^41^, Andrea Dietrich^444,445^, Katharina Domschke^30^, Peter Falkai^169,34^, Thomas V Fernandez^446^, Daniel A Geller^302^, Zachary F Gerring^41,54^, Fernando S Goes^227^, Hans J Grabe^411^, Marco A Grados^227^, Erica L Greenberg^302^, Jakob Grove^155,156,58,62,63,76^, Edna Grünblatt^447,448,449^, Jan Haavik^112,113^, Kristen Hagen^450,451,452^, Gregory L Hanna^453^, Bjarne Hansen^450,454^, Gary A Heiman^261,455^, Pieter J Hoekstra^444,445^, David M Hougaard^187,208,63^, James A Knowles^260,261,262^, Jaakko Kaprio^66^, Norbert Kathmann^77^, Julia Klawohn^456,77^, Gerd Kvale^113,457^, Nuria Lanzagorta^458^, Stephanie Le Hellard^459^, Daniel F Levey^50,51^, Christine Lochner^52^, Jurjen J Luykx^123,283,284^, Fabio Macciardi^460^, Brion S Maher^461^, Irene A Malaty^462^, David Mataix-Cols^463,75^, Carol A Mathews^291,292^, Manuel Mattheisen^26,58,59,60,61,62,63^, Nicole CR McLaughlin^464,465^, Euripedes C Miguel^466^, Humberto Nicolini^458,467^, Erika L Nurmi^468,469^, Michael S Okun^462^, Danielle Posthuma^133,134,135,136^, Raquel Rabionet^470,471,472,88^, Alfredo Ramirez^473,474,475,476,477^, Josep Antoni Ramos-Quiroga^137,138,87,90,95,96,97^, Marta Ribasés^87,88,89,90,97^, Renata Rizzo^478^, Cristina Rodriguez-Fontenla^479^, Jeremiah M Scharf^103,157,480^, Elles de Schipper^75^, Harvey S Singer^481^, María Soler Artigas^87,88,89,90,97^, Dan J Stein^69^, Murray B Stein^18,70^, Eric A Storch^482^, Nora I Strom^61,75,76,77^, Jackson G Thorp^41,54^, Jeremy Veenstra-VanderWeele^483,484^, Michael Wagner^287,474,477^, Christopher P Walker^261,455^, Dongmei Yu^103,84^, Gwyneth Zai^47,86^

### Post-Traumatic Stress Disorder Working Group

Søren B Andersen^485^, Helga Ask^4,5^, Sintia I Belangero^6,7^, Laura J Bierut^334^, Gerome Breen^15,16,17^, Rodrigo A Bressan^408^, Sandra A Brown^18,19^, Carolina M Carvalho^410,7^, Chia-Yen Chen^486^, Jonathan RI Coleman^15,16,17^, Lucia Colodro-Conde^129^, Nikolaos P Daskalakis^381,84^, Jürgen Deckert^26^, Seth G Disner^487,488^, Anna R Docherty^21,27,28,29^, Norah C Feeny^489^, Ary Gadelha^408^, Scott D Gordon^40,41^, Lana R Grasser^490^, Magali Haas^491^, Kelly M Harrington^492,493^, Victor M Hesselbrock^494^, Mohammed H Ibrahim^255^, Seyma Katrinli^495^, James L Kennedy^46,47,48,49^, Nathan A Kimbrel^496,497,498^, Karestan C Koenen^157,302,499^, Kristi Krebs^237^, Kelli Lehto^237^, Daniel F Levey^50,51^, Jurjen J Luykx^123,283,284^, Jessica L Maples-Keller^332^, Sarah E Medland^128,129,40,41^, Jacquelyn L Meyers^500^, Janitza L Montalvo-Ortiz^246,501^, Charles P Morris^54^, Vanessa K Ota^6,7^, Pedro M Pan^7^, Robert H Pietrzak, Renato Polimanti^246^, Richard J Rosenblum^502^, Barbara O Rothbaum^332^, Bart PF Rutten^123,84^, Nancy L Saccone^432,433^, Giovanni A Salum^415,416,417,418^, Marcos L Santoro^419,7^, Soraya Seedat^503^, Andrey A Shabalin^21,28^, Alicia K Smith^332,495,504^, Dan J Stein^69^, Murray B Stein^18,70^, Ralph E Tarter^505^, Clement C Zai^47,84,85^, Gwyneth Zai^47,86^

### Schizophrenia Working Group

Muhammad Ayub^506^, Nicholas Bass^173^, Bernhard T Baune^190,191,192,193^, Sintia I Belangero^6,7^, Tim B Bigdeli^200,201,202,203^, Rodrigo A Bressan^408^, Dominique Campion^507,508^, Boris Chaumette^350,380^, Sven Cichon^215,216,36,37^, David Cohen^509,510,511^, Angel Consoli^509,510^, Marta Di Forti^17^, Johan G Eriksson^512,513^, Olga Yu Fedorenko^514,515^, Josef Frank^152^, Robert Freedman^516^, Janice M Fullerton^241,242,243,244^, Ary Gadelha^408^, Raul R Gainetdinov^517,518^, Marianna Giannitelli^509,510^, Ina Giegling^519^, Stephen J Glatt^153,461^, Stephanie Godard^520^, Jakob Grove^155,156,58,62,63,76^, Olivier Guillin^507,508,521^, Annette M Hartmann^522^, Svetlana A Ivanova^514,523^, James A Knowles^260,261,262^, James L Kennedy^46,47,48,49^, Alexander O Kibitov^524,525^, Bettina Konte^522^, Claudine Laurent-Levinson^509,510^, Anastasia Levchenko^526^, Douglas F Levinson^412^, Qingqin S Li^280,281^, Jurjen J Luykx^123,283,284^, Brion S Maher^461^, Morten Mattingsdal^293,33^, Andrew McQuillin^173^, Sandra M Meier^64^, Robin Murray^527^, Merete Nordentoft^310,311,63^, Cristiano Noto^408^, Michael C O’Donovan^131,132^, Roel A Ophoff^239,240,272,316^, Vanessa K Ota^6,7^, Michael J Owen^131,132^, Danielle Posthuma^133,134,135,136^, Diego Quattrone^17^, Marcella Rietschel^152^, Brien P Riley^414^, Dan Rujescu^522^, Bart PF Rutten^123,84^, Safaa Saker-Delye^528^, Marcos L Santoro^419,7^, SIbylle G Schwab^529,530^, Alessandro Serretti^340,341^, Fabian Streit^152^, Jana Strohmaier^152^, Florence Thibaut^531,532^, Marquis P Vawter^352^, Bradley T Webb^29,420^, Thomas Werge^310,357,358,63^, Dieter B Wildenauer^533^, Stephanie H Witt^151,152^, Clement C Zai^47,84,85^

### Substance Use Disorders Working Group

Daniel E Adkins^1,2^, Arpana Agrawal^334^, Silvia Alemany^87,88,89,90^, David AA Baranger^534^, Anthony J Batzler^188^, Joanna M Biernacka^188,198,199^, Laura J Bierut^334^, Tim B Bigdeli^200,201,202,203^, Jason D Boardman^535^, Joseph M Boden^382^, Ryan Bogdan^534^, Sandra A Brown^18,19^, Karhleen K Bucholz^334^, Doo-Sup Choi^536,537^, Sarah MC Colbert^232^, Brandon J Coombes^188,198^, William E Copeland^23^, Joseph D Deak^246,538^, Marta Di Forti^17^, Nancy Diazgranados^539^, Danielle M Dick^422,423^, Anna R Docherty^21,27,28,29^, Alexis C Edwards^540^, Jerome C Foo^152^, Josef Frank^152^, Raul R Gainetdinov^517,518^, Ina Giegling^519^, Alison M Goate^232^, David Goldman^541^, Laura M Hack^542^, Dana B Hancock^426^, Kathleen Mullan Harris^543^, Annette M Hartmann^522^, Sarah M Hartz^334^, Alexander S Hatoum^534,544^, Caroline Hayward^545^, Andrew C Heath^334^, John K Hewitt^546^, Per Hoffmann^215,216,36^, Christian J Hopfer^516^, Daniel P Howrigan^117,84^, Emma C Johnson^334^, Eric O Johnson^420,428^, Jaakko Kaprio^66^, Victor M Karpyak^199^, Martin A Kennedy^386^, Alexander O Kibitov^524,525^, Bettina Konte^522^, Henry R Kranzler^118,119,120^, Kenneth S Krauter^547^, Evgeny M Krupitsky^525,548^, Samuel Kuperman^549^, Jari Lahti^43,550^, Marius Lahti-Pulkkinen^43,551,552^, Dongbing Lai^234^, Anastasia Levchenko^526^, Daniel F Levey^50,51^, Penelope A Lind^41,54^, Jurjen J Luykx^123,283,284^, Pamela A Madden^334^, Hermine HM Maes, Brion S Maher^461^, Nicholas G Martin^40,41,57^, Sarah E Medland^128,129,40,41^, Jacquelyn L Meyers^500^, Alex P Miller^334^, Janitza L Montalvo-Ortiz^246,501^, John I Nurnberger Jr^313^, Abraham A Palmer^18,553^, Rohn H Palmer^356,554,555^, Teemu Palviainen^66^, John F Pearson^556^, Roseann E Peterson^67^, Renato Polimanti^246^, Bernice Porjesz^557^, Ulrich W Preuss^558,559^, Diego Quattrone^17^, Josep Antoni Ramos-Quiroga^137,138,87,90,95,96,97^, Marta Ribasés^87,88,89,90,97^, John P Rice^334^, Brien P Riley^414^, Daniel M Rosenblum^560^, Richard J Rosenblum^502^, Dan Rujescu^522^, Nancy L Saccone^432,433^, Sandra Sanchez-Roige^18,561^, Norbert Scherbaum^562^, Andrey A Shabalin^21,28^, Richard Sherva^425,435^, María Soler Artigas^87,88,89,90,97^, Fabian Streit^152^, Ralph E Tarter^505^, Michael Vanyukov^563^, Tamara L Wall^18^, Raymond K Walters^117,84^, Bradley T Webb^29,420^, Robbee Wedow^234,564^, Stanley H Weiss^560^, Leah Wetherill^234^, Stephanie H Witt^151,152^, Norbert Wodarz^565^, Stephanie Zellers^436,66^, Haitao Zhang^566^, Hongyu Zhao^567^, Hang Zhou^246,365,51^, Peter Zill^169^, Lea Zillich^152^

### Tourette Syndrome Working Group

Cathy L Barr^100,568,569^, Csaba Barta^439^, Julia Boberg^75^, Beatriz Camarena^441^, Danielle C Cath^442,443^, James J Crowley^347^, Andrea Dietrich^444,445^, Thomas V Fernandez^446^, Zachary F Gerring^41,54^, Marco A Grados^227^, Erica L Greenberg^302^, Gary A Heiman^261,455^, Pieter J Hoekstra^444,445^, James A Knowles^260,261,262^, Nuria Lanzagorta^458^, Christine Lochner^52^, Fabio Macciardi^460^, Irene A Malaty^462^, David Mataix-Cols^463,75^, Carol A Mathews^291,292^, Manuel Mattheisen^26,58,59,60,61,62,63^, Euripedes C Miguel^466^, Kirsten R Müller-Vahl^570^, Humberto Nicolini^458,467^, Erika L Nurmi^468,469^, Michael S Okun^462^, Peristera Paschou^413^, Renata Rizzo^478^, Paul Sandor^47^, Jeremiah M Scharf^103,157,480^, Elles de Schipper^75^, Harvey S Singer^481^, Nora I Strom^61,75,76,77^, Dongmei Yu^103,84^, Gwyneth Zai^47,86^

^1^Department of Sociology, Univerity of Utah. ^2^Graduate Program in Statistics, Unviersity of Utah. ^3^Department of Psychology, School of Social Sciences, University of Mannheim, Germany. ^4^PROMENTA research center, University of Oslo. ^5^PsychGen Centre for Genetic Epidemiology and Mental Health, Norwegian Institute of Public Health. ^6^Department of Morphology and Genetics, Universidade Federal de Sao Paulo. ^7^Laboratory of Integrative Neuroscience - Universidade Federal de Sao Paulo, Brasil. ^8^Department of Mental Health, Faculty of Medicine and Health Sciences, Norwegian University of Science and Technology, Trondheim, Norway. ^9^Faculty of Nursing and Health Sciences, Nord University, Levanger, Norway. ^10^Department of Research and Innovation, Division of Clinical Neuroscience, Oslo University Hospital, Oslo, Norway. ^11^HUNT Center for Molecular and Clinical Epidemiology, Department of Public Health and Nursing, Faculty of Medicine and Health Sciences, Norwegian University of Science and Technology, Trondheim, Norway. ^12^Institute of Clinical Medicine, University of Oslo, Oslo, Norway. ^13^K. G. Jebsen Center for Genetic Epidemiology, Department of Public Health and Nursing, Faculty of Medicine and Health Sciences, Norwegian University of Science and Technology, Trondheim, Norway. ^14^Research and Communication Unit for Musculoskeletal Health, Division of Clinical Neuroscience, Oslo University Hospital, Ullevål, Oslo, Norway. ^15^NIHR Maudsley Biomedical Research Centre, South London and Maudsley NHS Trust, London, UK. ^16^NIHR Maudsley BRC, King’s College London, London, GB. ^17^Social, Genetic & Developmental Psychiatry Centre, Institute of Psychiatry, Psychology & Neuroscience, King’s College London, UK. ^18^Department of Psychiatry, University of California San Diego, La Jolla, CA, USA. ^19^Department of Psychology, University of California San Diego. ^20^Department of Psychiatry, Lausanne University Hospital and University of Lausanne, Lausanne, Switzerland. ^21^Department of Psychiatry, University of Utah School of Medicine, Salt Lake City, UT, USA. ^22^Huntsman Mental Health Institute, University of Utah School of Medicine. ^23^Department of Psychiatry, University of Vermont. ^24^Nic Waals Institute, Lovisenberg Diaconal Hospital. ^25^Max-Planck-Insitute of Psychiatry, Department Genes and Environment, Munich, Germany. ^26^Department of Psychiatry, Psychosomatics and Psychotherapy, University of Würzburg, Würzburg, Germany. ^27^Center for Genomic Medicine, Salt Lake City, UT USA. ^28^Huntsman Mental Health Institute, Salt Lake City, UT USA. ^29^Virginia Institute for Psychiatric and Behavioral Genetics, Virginia Commonwealth University, Richmond, VA, USA. ^30^Department of Psychiatry and Psychotherapy, Medical Center - University of Freiburg, Faculty of Medicine, University of Freiburg, Freiburg, Germany. ^31^Department of Østmarka, Division of Mental Health Care, St. Olavs Hospital, Trondheim University Hospital, Trondheim, Norway. ^32^Department of Psychiatry, Sørlandet Hospital, Kristiansand/Arendal, Norway. ^33^NORMENT Centre, Division of Mental Health and Addiction, Oslo University Hospital, Oslo, Norway. ^34^Max-Planck-Institute for Psychiatry, Munich, Germany. ^35^Centre for Human Genetics, University of Marburg, Marburg, Germany. ^36^Institute of Human Genetics, University of Bonn, School of Medicine & University Hospital Bonn, Bonn, Germany. ^37^Institute of Neuroscience and Medicine (INM-1), Research Center Juelich, Juelich, Germany. ^38^Department of Medical Epidemiology and Biostatistics, Karolinska Institutet, Stockholm, Sweden. ^39^School of Medical Sciences, Örebro University, Faculty of Medicine and Health, Örebro, Sweden. ^40^Genetics and Computational Biology, QIMR Berghofer Medical Research Institute, Brisbane, QLD, AU. ^41^Mental Health and Neuroscience Research Program, QIMR Berghofer Medical Research Institute, Brisbane, Australia. ^42^Brain and Mind Centre, The University of Sydney Australia. ^43^Department of Psychology and Logopedics, University of Helsinki and Helsinki University Central Hospital, Helsinki, FInland. ^44^SleepWell Research Program, Faculty of Medicine, University of Helsinki, Helsinki, Finland. ^45^Centre for Clinical Brain Sciences, The University of Edinburgh, UK. ^46^Campbell Family Mental Health Research Institute, Centre for Addiction and Mental Health, Toronto, ON, CA. ^47^Department of Psychiatry, Institute of Medical Science, University of Toronto, Toronto, Canada. ^48^Institute of Medical Sciences, University of Toronto, Toronto, ON, CA. ^49^Neurogenetics Section, Centre for Addiction and Mental Health, Toronto, ON, CA. ^50^Division of Human Genetics, Department of Psychiatry, Yale University School of Medicine, New Haven, CT, USA. ^51^Veterans Affairs Connecticut Healthcare Center, West Haven, CT, USA. ^52^SA MRC Unit on Risk and Resilience in Mental Disorders, Department of Psychiatry, Stellenbosch University, South Africa. ^53^QIMR Berghofer Medical Research Institute, Brisbane, Queensland, Australia. ^54^School of Biomedical Sciences, Faculty of Health, Queensland University of Technology, Brisbane, Australia. ^55^Department of Psychiatry, University of Tartu, Tartu, Estonia. ^56^Faculty of Medicine, Department of Medicine, Centre for Neuropsychopharmacology, Division of Brain Sciences, Imperial College London, London, UK. ^57^School of Psychology, The University of Queensland, Brisbane, QLD, AU. ^58^Department of Biomedicine - Human Genetics, Aarhus University, Aarhus, DK. ^59^Department of Clinical Neuroscience, Centre for Psychiatry Research, Karolinska Institutet, Stockholm, Sweden. ^60^Department of Community Health and Epidemiology and Faculty of Computer Science, Dalhousie University, Halifax, NS, Canada. ^61^Institute of Psychiatric Phenomics and Genomics (IPPG), University Hospital, LMU Munich, Munich, Germany. ^62^iSEQ, Center for Integrative Sequencing, Aarhus University, Aarhus, DK. ^63^The Lundbeck Foundation Initiative for Integrative Psychiatric Research, iPSYCH, Aarhus, Denmark. ^64^Department of Psychiatry, Dalhousie University, Halifax, Canada. ^65^Department of Clinical Psychology, Experimental Psychopathology and Psychotherapy, University of Marburg, Germany. ^66^Institute for Molecular Medicine Finland (FIMM), University of Helsinki, Helsinki, Finland. ^67^Department of Psychiatry and Behavioral Sciences, Institute for Genomics in Health, State University of New York Downstate Health Sciences University, Brooklyn, NY. ^68^Institute for Medical Informatics, Biometry and Epidemiology, University Hospital Essen, University Duisburg-Essen, Essen, Germany. ^69^SAMRC Unit on Risk & Resilience in Mental Disorders, Department of Psychiatry & Neuroscience Institute, University of Cape Town. ^70^VA San Diego Healthcare System, San Diego, CA, USA. ^71^Department of Neuroscience, Norges Teknisk Naturvitenskapelige Universitet Fakultet for naturvitenskap og teknologi, Trondheim, NO. ^72^Department of Psychiatry, Hospital Namsos, Namsos, NO. ^73^Department of Psychiatry, Hospital Namsos, Nord-Trøndelag Health Trust, Namsos, Norway. ^74^Department of Psychiatry and Psychotherapy, Charité Campus Mitte, Charité - Universitätsmedizin Berlin, Germany. ^75^Centre for Psychiatry Research, Department of Clinical Neuroscience, Karolinska Institutet & Stockholm Health Care Services, Region Stockholm, Stockholm, Sweden. ^76^Department of Biomedicine, Aarhus University, Aarhus, Denmark. ^77^Department of Psychology, Humboldt-Universität zu Berlin, Berlin, Germany. ^78^BioCore - Bioinformatics Core Facility, Norwegian University of Science and Technology, Trondheim. Norway. ^79^Clinic of Laboratory Medicine, St.Olavs Hospital, Trondheim University Hospital, Trondheim, Norway. ^80^Department of Clinical and Molecular Medicine, Norwegian University of Science and Technology, Trondheim, Norway. ^81^Department of Child and Adolescent Psychiatry, Erasmus University Medical Center, Rotterdam, Netherlands. ^82^Department of Social and Behavioral Sciences, Harvard T.H. Chan School of Medicine, Boston. ^83^Department of Neurology, Oslo University Hospital, Oslo, Norway. ^84^Stanley Center for Psychiatric Research, Broad Institute of MIT and Harvard, Cambridge, MA, USA. ^85^Tanenbaum Centre for Pharmacogenetics, Campbell Family Mental Health Research Institute, Centre for Addiction and Mental Health, Toronto, Canada. ^86^Neurogenetics Section, Campbell Family Mental Health Research Institute, Centre for Addiction and Mental Health, Toronto, Canada. ^87^Biomedical Network Research Centre on Mental Health (CIBERSAM), Instituto de Salud Carlos III, Madrid, Spain. ^88^Department of Genetics, Microbiology and Statistics, Faculty of Biology, Universitat de Barcelona, Barcelona, Catalonia, Spain. ^89^Department of Mental Health, Hospital Universitari Vall d’Hebron, Barcelona, Spain. ^90^Psychiatric Genetics Unit, Group of Psychiatry, Mental Health and Addiction, Vall d’Hebron Research Institute (VHIR), Universitat Autònoma de Barcelona, Barcelona, Spain. ^91^ADHD and Developmental Psychiatry Programs, Hospital de Clínicas de Porto Alegre, Universidade Federal do Rio Grande do Sul, Porto Alegre, RS, Brazil. ^92^Department of Genetics, Instituto de Biociências, Universidade Federal do Rio Grande do Sul, Porto Alegre, RS, Brazil. ^93^Department of Biological Psychology, Vrije Universiteit, Amsterdam, The Netherlands. ^94^Netherlands Twin Register, Vrije Universiteit Amsterdam. ^95^Biomedical Network Research Centre on Mental Health (CIBERSAM), Barcelona, Catalonia, Spain. ^96^Department of Psychiatry and Forensic Medicine, Universitat Autònoma de Barcelona, Barcelona, ES. ^97^Department of Psychiatry, Hospital Universitari Vall d’Hebron, Barcelona, ES. ^98^SJD MIND Schools Program, Hospital Sant Joan de Déu, Institut de Recerca Sant Joan de Déu, Esplugues de Llobregat, Spain. ^99^Department of Global Public Health and Primary Care, University of Bergen, Årstadveien 17, 5009 Bergen, Norway. ^100^Program in Neurosciences and Mental Health, Hospital for Sick Children, Toronto, ON, Canada. ^101^Department of Psychiatry and Legal Medicine, Universitat Autònoma de Barcelona. ^102^Fundació Privada d’Investigació Sant Pau (FISP), Barcelona, Spain. ^103^Center for Genomic Medicine, Massachusetts General Hospital, Harvard Medical School, Boston MA, USA. ^104^Department of Psychiatry, Harvard Medical School. ^105^Department of Psychiatry, Norton College of Medicine, SUNY Upstate Medical University. ^106^Department of Human Genetics, Radboud University Medical Center, Nijmegen, The Netherlands. ^107^Dept Cognitive Neuroscience, Radboud University Medical Center, Nijmegen, The Netherlands. ^108^Donders Institute for Brain, Cognition and Behaviour, Radboud University, Nijmegen, The Netherlands. ^109^Center for Applied Genomics, The Children’s Hospital of Philadelphia, Philadelphia, PA, USA. ^110^Department of Pediatrics, University of Pennsylvania, Philadelphia, PA, USA. ^111^Department of Psychiatry and Legal Medicine, Faculty of Medicine, Universidade Federal do Rio Grande do Sul, Porto Alegre, Brazil. ^112^Department of Biomedicine, University of Bergen, Norway. ^113^Division of Psychiatry, Haukeland University Hospital, Bergen, Norway. ^114^Turner Institute for Brain and Mental Health, School of Psychological Sciences, Monash University, VIC, Australia. ^115^Center for Translational Neuro- and Behavioral Sciences, University Hospital Essen, University of Duisburg-Essen, Essen, Germany. ^116^Department of Child and Adolescent Psychiatry, Psychosomatics and Psychotherapy, University Hospital Essen, University of Duisburg-Essen, Duisburg, DE. ^117^Analytic and Translational Genetics Unit, Department of Medicine, Massachusetts General Hospital, Boston, MA. ^118^Center for Studies of Addiction, University of Pennsylvania Perelman School of Medicine, Philadelphia, PA, USA. ^119^Department of Psychiatry, University of Pennsylvania Perelman School of Medicine, Philadelphia, PA, USA. ^120^Mental Illness Research, Education and Clinical Center, Crescenz VAMC, Philadelphia, PA, USA. ^121^Institute of Neuroscience and Physiology, University of Gothenburg, Gothenburg, Sweden. ^122^Department of Child and Adolescent Psychiatry, Psychosomatics and Psychotherapy, University Hospital Wuerzurg, Wuerzburg, Germany. ^123^Department of Psychiatry and Neuropsychology, School for Mental Health and Neuroscience, Maastricht University Medical Center, Maastricht, the Netherlands. ^124^Department of Human Genetics, McGill University, Montréal, QC, Canada. ^125^Montreal Neurological Institute and Hospital, McGill University, Montréal, QC, Canada. ^126^UCLA Semel Institute for Neuroscience and Human Behavior, University of California, Los Angeles CA. 90095. ^127^David Geffen School of Medicine at UCLA. ^128^School of Psychology and Counselling, Queensland University of Technology, Queensland, Australia. ^129^School of Psychology, University of Queensland, Queensland, Australia. ^130^Medical and Population Genetics, Broad Institute, Cambridge, MA, US. ^131^Centre for Neuropsychiatric Genetics and Genomics, School of Medicine, Cardiff University. ^132^Medical Research Council Centre for Neuropsychiatric Genetics and Genomics, Division of Psychological Medicine and Clinical Neurosciences, Cardiff University, Cardiff, GB. ^133^Department of Clinical Genetics, Amsterdam Neuroscience, Vrije Universiteit Medical Center, Amsterdam, NL. ^134^Department of Complex Trait Genetics, Center for Neurogenomics and Cognitive Research, Amsterdam Neuroscience, Vrije Universiteit Amsterdam, Amsterdam, NL. ^135^Vrije Universiteit Amsterdam, Department Complex Trait Genetics, Amsterdam, The Netherlands. ^136^Vrije Universiteit Medical Centre, Dept Child and Adolescent Psychiatry, Amsterdam, The Netherlands. ^137^Department of Mental Health. Vall Hebron University Hospital. Barcelona. Catalonia. Spain. ^138^Group of Psychiatry, Mental Health and Adicctions. Vall Hebron Research Institute. Barcelona. Spain. ^139^Department of Psychiatry, Psychosomatic Medicine and Psychotherapy, University Hospital Frankfurt, Frankfurt am Main, DE. ^140^Fraunhofer Institute for Translational Medicine and Pharmacology ITMP, Theodor-Stern-Kai 7, 60596 Frankfurt am Main, Germany. ^141^Goethe University Frankfurt, University Hospital, Department of Psychiatry, Psychosomatic Medicine and Psychotherapy, Frankfurt, Germany. ^142^Department of Physiology and Biophysics, Instituto de Ciencias Biomedicas Universidade de Sao Paulo, São Paulo, Brazil. ^143^Laboratory of Physiological Genomics of Mental Health (PhysioGen Lab), Institute of Biomedical Sciences, University of Sao Paulo, São Paulo, Brazil. ^144^Dept. of Molecular Genetics and McLaughlin Centre, University of Toronto, Toronto, ON, Canada. ^145^The Centre for Applied Genomics and Dept. of Genetics and Genome Biology, The Hospital for Sick Children, Toronto, ON, Canada. ^146^Institute of Psychiatry, Psychology and Neuroscience, King’s College London. ^147^School of Medicine, Aarhus University. ^148^School of Psychology, University of Hong Kong. ^149^Department of Child and Adolescent Psychiatry, University of Zurich, Switzerland. ^150^Department of Mental Health and Suicide, Norwegian Institute of Mental Health, Oslo, Norway. ^151^Central Institute of Mental Health, Medical Faculty Mannheim, Heidelberg University, Mannheim, Germany. ^152^Department of Genetic Epidemiology in Psychiatry, Central Institute of Mental Health, Medical Faculty Mannheim, Heidelberg University, Mannheim, Germany. ^153^Department of Psychiatry and Behavioral Sciences, State University of New York Upstate Medical University, Syracuse, NY. ^154^University of Illinois, Chicago, Illinois, USA. ^155^Bioinformatics Research Centre, Aarhus University, Aarhus, Denmark. ^156^Center for Genomics and Personalized Medicine, Aarhus, Denmark. ^157^Broad Institute of MIT and Harvard, Cambridge, MA, USA. ^158^Carolina Institute for Developmental Disabilities. ^159^University of North Carolina at Chapel Hill, Chapel Hill, North Carolina USA. ^160^Department of Psychiatry and Behavioral Sciences, UCSF Weill Institute for Neurosciences, University of California, San Francisco, San Francisco, CA 94158, USA. ^161^Institute of Developmental and Regenerative Medicine, Department of Paediatrics, University of Oxford, Oxford, OX3 7TY, UK. ^162^New York Genome Center, New York, NY 10013, USA. ^163^Center for Applied and Translational Genomics (CATG), MBRU, Dubai Health, Dubai, UAE. ^164^GenomeArc Inc., Mississauga, Ontario, Canada. ^165^Department of Psychiatry and Program in Genetics and Genome Biology, The Hospital for Sick Children, Toronto, ON, Canada. ^166^Departments of Psychiatry and Psychology, University of Cambridge, UK. ^167^Institute for Human Genetics, Department of Psychiatry and Behavioral Science, Weill Institute for Neurosciences, University of California San Francisco, San Francisco CA, USA. ^168^Department of Clinical Sciences, Psychiatry, Umeå University Medical Faculty, Umeå, SE. ^169^Department of Psychiatry and Psychotherapy, LMU University Hospital, LMU Munich, Munich, Germany. ^170^Department of Psychiatric Research, Diakonhjemmet Hospital, Oslo, NO. ^171^NORMENT, KG Jebsen Centre for Psychosis Research, Oslo University Hospital, Oslo, NO. ^172^National Centre for Register-based Research, Aarhus University, Aarhus, Denmark. ^173^Division of Psychiatry, University College London, London, UK. ^174^Department of Neuroscience, Istituto Di Ricerche Farmacologiche Mario Negri IRCCS, Milano, IT. ^175^National Institute of Mental Health, Klecany, CZ. ^176^Institute of Environmental Medicine, Karolinska Institutet, Stockholm, Sweden. ^177^Department of Psychiatry and Behavioral Neuroscience, University of Chicago, Chicago, IL, US. ^178^Northwestern University, Chicago, IL, US. ^179^Department of Neurology, Klinikum rechts der Isar, School of Medicine, Technical University of Munich, Munich, Germany. ^180^Division of Mental Health and Addiction, Oslo University Hospital, Oslo, NO. ^181^Psychiatry, Berkshire Healthcare NHS Foundation Trust, Bracknell, GB. ^182^National and Kapodistrian University of Athens, 2nd Department of Psychiatry, Attikon General Hospital, Athens, Greece. ^183^Department of Psychiatry and Psychotherapy, Charité - Universitätsmedizin, Berlin, DE. ^184^Center for Molecular Medicine, Karolinska University Hospital, Stockholm, Sweden. ^185^Department of Molecular Medicine and Surgery, Karolinska Institutet, Stockholm, Sweden. ^186^Department of Psychiatry, Sungkyunkwan University School of Medicine, Samsung Medical Center, Seoul, South Korea. ^187^Center for Neonatal Screening, Department for Congenital Disorders, Statens Serum Institut, Copenhagen, DK. ^188^Department of Quantitative Health Sciences, Mayo Clinic, Rochester, MN, USA. ^189^Department of Psychiatry and Psychotherapy, University Hospital Carl Gustav Carus, Technische Universität Dresden, Dresden, DE. ^190^Department of Psychiatry, Melbourne Medical School, The University of Melbourne, Melbourne, VIC, Australia. ^191^Department of Psychiatry, University of Melbourne, Melbourne, Australai. ^192^Department of Psychiatry, University of Münster, Münster, Germany. ^193^The Florey Institute of Neuroscience and Mental Health, The University of Melbourne, Parkville, VIC, Australia. ^194^APHP Nord, DMU Neurosciences, GHU Saint Louis-Lariboisière-Fernand Widal, Département de Psychiatrie et de Médecine Addictologique, Paris, France. ^195^Université de Paris, INSERM, Optimisation Thérapeutique en Neuropsychopharmacologie, UMRS-1144, Paris, France. ^196^Medical University of Graz, Department of Psychiatry and Psychotherapeutic Medicine, Graz, Austria. ^197^Psychiatry, University of Pennsylvania, Philadelphia, PA, US. ^198^Department of Health Sciences Research, Mayo Clinic, Rochester, MN, US. ^199^Department of Psychiatry and Psychology, Mayo Clinic, Rochester, MN, USA. ^200^Department of Psychiatry and Behavioral Sciences, Institute for Genomics in Health, Department of Epidemiology and Biostatistics, State University of New York Downstate Health Sciences University, Brooklyn, NY. ^201^Department of Psychiatry and Behavioral Sciences, SUNY Downstate Health Sciences University, Brooklyn, NY, US. ^202^Department of Veterans Affairs (VA) New York Harbor Healthcare System, Brooklyn, NY. ^203^VA NY Harbor Healthcare System, Brooklyn, NY, US. ^204^Division of Psychiatry, University of Edinburgh, Edinburgh, GB. ^205^Center for Statistical Genetics and Department of Biostatistics, University of Michigan, Ann Arbor, MI, US. ^206^Psychiatry, Brain Center UMC Utrecht, Utrecht, NL. ^207^Department of Biomedicine and the iSEQ Center, Aarhus University, Aarhus, DK. ^208^Department for Congenital Disorders, Statens Serum Institut, Copenhagen, Denmark. ^209^Psychiatry, University of California San Francisco, San Francisco, CA, US. ^210^University of Newcastle, Newcastle, NSW, AU. ^211^School of Psychiatry, University of New South Wales, Sydney, NSW, AU. ^212^University of Queensland, Brisbane, QLD, AU. ^213^Department of Psychiatry, Mood Disorders Program, McGill University Health Center, Montreal, QC, CA. ^214^Department of Psychiatry, Icahn School of Medicine at Mount Sinai, New York, NY, US. ^215^Department of Biomedicine, University of Basel, Basel, Switzerland. ^216^Institute of Medical Genetics and Pathology, University Hospital Basel, Basel, Switzerland. ^217^Neuropsychiatric Genetics Research Group, Dept of Psychiatry and Trinity Translational Medicine Institute, Trinity College Dublin, Dublin, IE. ^218^Department of Translational Research in Psychiatry, Max Planck Institute of Psychiatry, Munich, DE. ^219^Department of Psychiatry, Universidad Autonoma de Nuevo Leon, Monterrey, Mexico. ^220^Department of Laboratory Medicine & Pathology, Mayo Clinic, Rochester, MN, US. ^221^Centre for Psychiatry, Queen Mary University of London, London, GB. ^222^UCL Genetics Institute, University College London, London, GB. ^223^Department of Psychiatry, Laboratory of Psychiatric Genetics, Poznan University of Medical Sciences, Poznan, PL. ^224^Center for Multimodal Imaging and Genetics, Departments of Neurosciences, Radiology, and Psychiatry, University of California, San Diego, CA, US. ^225^Institute for Translational Psychiatry, University of Münster, Münster, Germany. ^226^Department of Psychiatry and Psychotherapy, University of Marburg, Marburg, Germany. ^227^Department of Psychiatry and Behavioral Sciences, Johns Hopkins School of Medicine, Baltimore, MD, USA. ^228^Department of Psychiatry, University of North Carolina at Chapel Hill, Chapel Hill, NC, USA. ^229^National and Kapodistrian University of Athens, 1st Department of Psychiatry, Eginition Hospital, Athens, Greece. ^230^Department of Medical Genetics, Oslo University Hospital Ullevål, Oslo, NO. ^231^NORMENT, Department of Clinical Science, University of Bergen, Bergen, NO. ^232^Department of Genetics and Genomic Sciences, Icahn School of Medicine at Mount Sinai, New York, NY, US. ^233^Biochemistry and Molecular Biology, Indiana University School of Medicine, Indianapolis, IN, US. ^234^Department of Medical & Molecular Genetics, Indiana University, Indianapolis, IN, US. ^235^Department of Genetics, Harvard Medical School, Boston, MA, US. ^236^Division of Endocrinology, Children’s Hospital Boston, Boston, MA, US. ^237^Estonian Genome Center, Institute of Genomics, University of Tartu, Tartu, Estonia. ^238^Academic Psychiatry, Newcastle University, Newcastle upon Tyne, GB. ^239^Center for Neurobehavioral Genetics, Semel Institute for Neuroscience and Human Behavior, David Geffen School of Medicine, University of California Los Angeles, Los Angeles, USA. ^240^Department of Psychiatry and Biobehavioral Science, Semel Institute for Neuroscience & Human Behavior, David Geffen School of Medicine, University of California Los Angeles, Los Angeles, CA, US. ^241^Neuroscience Research Australia, Randwick, NSW, Australia. ^242^Neuroscience Research Australia, Sydney, NSW, AU. ^243^School of Medical Sciences, University of New South Wales, Sydney, NSW, AU. ^244^University of New South Wales, Faculty of Medicine, School of Biomedical Science, Kensington, NSW, Australia. ^245^Department of Psychiatry and Psychotherapy, University Medical Center Göttingen, Göttingen, DE. ^246^Department of Psychiatry, Yale School of Medicine, New Haven, CT, USA. ^247^Departments of Genetics and Neuroscience, Yale University School of Medicine, New Haven, CT, USA. ^248^Department of Human Genetics, University of Chicago, Chicago, IL, US. ^249^Department of Psychological Sciences, University of Missouri, Columbia, MO, US. ^250^Psychological Medicine, University of Worcester, Worcester, GB. ^251^Biometric Psychiatric Genetics Research Unit, Alexandru Obregia Clinical Psychiatric Hospital, Bucharest, Romania. ^252^Mental Health Department, University Regional Hospital, Biomedicine Institute (IBIMA), Málaga, ES. ^253^Department of Psychiatry, Seoul National University College of Medicine, Seoul, South Korea. ^254^Landspitali University Hospital, Reykjavik, IS. ^255^Department of Psychology, Eberhard Karls Universität Tübingen, Tubingen, Germany. ^256^Department of Psychiatry, Fujita Health University School of Medicine, Toyoake, Japan. ^257^University of Western Australia, Nedlands, WA, AU. ^258^Faculté de Santé, Université Paris Est, Créteil, FR. ^259^Neuropsychiatrie Translationnelle, Inserm U955, Créteil, FR. ^260^Cell Biology, SUNY Downstate Medical Center College of Medicine, Brooklyn, NY, US. ^261^Department of Genetics and the Human Genetics Institute of New Jersey, Rutgers University. ^262^Institute for Genomic Health, SUNY Downstate Medical Center College of Medicine, Brooklyn, NY, US. ^263^International Max Planck Research School for Translational Psychiatry (IMPRS-TP), Munich, Germany. ^264^Laboratory for Statistical and Translational Genetics, RIKEN Center for Integrative Medical Sciences, Yokohama, Japan. ^265^Laboratory of Complex Trait Genomics, Department of Computational Biology and Medical Sciences, Graduate School of Frontier Sciences, The University of Tokyo, Tokyo, Japan. ^266^ISGlobal, Barcelona, ES. ^267^University of Patras, School of Health Sciences, Department of Pharmacy, Laboratory of Pharmacogenomics and Individualized Therapy, Patras, Greece. ^268^RIKEN Center for Integrative Medical Sciences, Yokohama, Japan. ^269^Psychiatry, Altrecht, Utrecht, NL. ^270^Psychiatry, GGZ inGeest, Amsterdam, NL. ^271^Psychiatry, VU medisch centrum, Amsterdam, NL. ^272^Department of Psychiatry, Erasmus MC, University Medical Center Rotterdam, Rotterdam, The Netherlands. ^273^Psychiatry, North East London NHS Foundation Trust, Ilford, GB. ^274^Clinic for Psychiatry and Psychotherapy, University Hospital Cologne, Cologne, DE. ^275^Department of Psychiatry and Addiction Medicine, Assistance Publique - Hôpitaux de Paris, Paris, FR. ^276^Department of Psychiatry, Korea University College of Medicine, Seoul, South Korea. ^277^Psychiatric and Neurodevelopmental Genetics Unit (PNGU), Massachusetts General Hospital, Boston, MA, US. ^278^HudsonAlpha Institute for Biotechnology, Huntsville, AL, US. ^279^Department of Medical & Molecular Genetics, King’s College London, London, GB. ^280^Janssen Research and Development, LLC, Titusville, NJ, 08560. ^281^Neuroscience Therapeutic Area, Janssen Research and Development, LLC, Titusville, NJ, US. ^282^Cancer Epidemiology and Prevention, M. Sklodowska-Curie National Research Institute of Oncology, Warsaw, PL. ^283^Department of Psychiatry, Amsterdam University Medical Center, Amsterdam, the Netherlands. ^284^GGZ inGeest Mental Health Care, Amsterdam, The Netherlands. ^285^Division of Psychiatry, Centre for Clinical Brain Sciences, University of Edinburgh, Edinburgh, UK. ^286^deCODE Genetics / Amgen, Reykjavik, IS. ^287^Department of Psychiatry and Psychotherapy, University Hospital Bonn, Bonn, Germany. ^288^Department of Medical Science and Public Health, University of Cagliari, Italy. ^289^Department of Pharmacology, Dalhousie University, Halifax, Nova Scotia, Canada. ^290^National and Kapodistrian University of Athens, Medical School, Clinical Biochemistry Laboratory, Attikon General Hospital, Athens, Greece. ^291^Department of Psychiatry and Genetics Institute, University of Florida, Gainesville, FL, USA. ^292^UF Center for OCD, Anxiety, and Related Disorders. ^293^Department of Medical Research, Bærum Hospital, Vestre Viken Hospital Trust, Rud, Norway. ^294^Research Institute, Lindner Center of HOPE, Mason, OH, US. ^295^Systems Genetics Working Group, Department of Genetics, Stellenbosch University, Stellenbosch, South Africa. ^296^Centre for Cognitive Ageing and Cognitive Epidemiology, University of Edinburgh, Edinburgh, GB. ^297^Genetic Cancer Susceptibility Group, International Agency for Research on Cancer, Lyon, FR. ^298^Human Genetics Branch, Intramural Research Program, National Institute of Mental Health, Bethesda, MD, US. ^299^Division of Mental Health and Addiction, University of Oslo, Institute of Clinical Medicine, Oslo, NO. ^300^Discipline of Psychiatry and Mental Health, School of Medicine and Health, University of New South Wales, Sydney, Australia. ^301^Institute for Molecular Bioscience, University of Queensland, Brisbane, Australia. ^302^Department of Psychiatry, Massachusetts General Hospital, Harvard Medical School, Boston, MA, USA. ^303^Psychiatry, St Olavs University Hospital, Trondheim, NO. ^304^Centre for Neuroimaging & Cognitive Genomics (NICOG), National University of Ireland Galway, Galway, IE. ^305^Psychosis Research Unit, Aarhus University Hospital - Psychiatry, Risskov, DK. ^306^Munich Cluster for Systems Neurology (SyNergy), Munich, DE. ^307^University of Liverpool, Liverpool, GB. ^308^Department of Neuropsychiatry, Seoul National University Bundang Hospital. ^309^Research/Psychiatry, Veterans Affairs San Diego Healthcare System, San Diego, CA, US. ^310^Department of Clinical Medicine, University of Copenhagen, Copenhagen, Denmark. ^311^Mental Health Services in the Capital Region of Denmark, Mental Health Center Copenhagen, University of Copenhagen, Copenhagen, Denmark. ^312^Psychiatry, Indiana University School of Medicine, Indianapolis, IN, US. ^313^Department of Psychiatry, Department of Medical and Molecular Genetics, Stark Neurosciences Research Institute, Indiana University School of Medicine, Indianapolis, IN, USA. ^314^Faculty of Medicine and Dentistry, University of Bergen, Bergen, NO. ^315^Department of Clinical Neuroscience and Center for Molecular Medicine, Karolinska Institutet at Karolinska University Hospital, Solna, Sweden. ^316^Department of Human Genetics, David Geffen School of Medicine, University of California Los Angeles, Los Angeles, CA, USA. ^317^Medical faculty, University Sarajevo School of Science and Technology, Sarajevo, Bosnia and Herzegovina. ^318^Human Genetics and Computational Biomedicine, Pfizer Global Research and Development, Groton, CT, US. ^319^University of Melbourne, VIC, AU. ^320^Erasmus MC, Faculty of Medicine and Health Sciences, Department of Pathology, Bioinformatics Unit, Rotterdam, the Netherlands. ^321^United Arab Emirates University, College of Medicine and Health Sciences, Department of Genetics and Genomics, Al-Ain, Abu Dhabi, UAE. ^322^United Arab Emirates University, College of Medicine and Health Sciences, Department of Pathology, Al-Ain, UAE. ^323^United Arab Emirates University, Zayed Center for Health Sciences, Al-Ain, Abu Dhabi, UAE. ^324^University of Patras School of Health Sciences, Department of Pharmacy. ^325^Division of Clinical Research, Massachusetts General Hospital, Boston, MA, US. ^326^Psychiatry, Harvard Medical School, Boston, MA, US. ^327^Department of Psychiatry, University of Oxford, Oxford, UK. ^328^Oxford Health NHS Foundation Trust, Warneford Hospital, Oxford, UK. ^329^Department of Neuroscience, Icahn School of Medicine at Mount Sinai, New York, NY, US. ^330^Estelle and Daniel Maggin Department of Neurology, Icahn School of Medicine at Mount Sinai, New York, NY, US. ^331^Ronald M. Loeb Center for Alzheimer’s Disease, Icahn School of Medicine at Mount Sinai, New York, NY, US. ^332^Department of Psychiatry and Behavioral Sciences, Emory University School of Medicine, Atlanta, GA, US. ^333^Outpatient Clinic for Bipolar Disorder, Altrecht, Utrecht, NL. ^334^Department of Psychiatry, Washington University, St. Louis, MO, USA. ^335^Department of Biochemistry and Molecular Biology II, Faculty of Pharmacy, University of Granada, ES. ^336^Institute of Neurosciences, Biomedical Research Center (CIBM), University of Granada, ES. ^337^Department of Neurology and Neurosurgery, McGill University, Faculty of Medicine, Montreal, QC, CA. ^338^Medicine, Psychiatry, Biomedical Informatics, Vanderbilt University Medical Center, Nashville, TN, US. ^339^Department of Psychiatry, University Hospital, Faculty of Medicine, University of Bonn, Bonn, Germany. ^340^Department of Biomedical and Neuromotor Sciences, University of Bologna, Bologna, Italy. ^341^Department of Medicine and Surgery, Kore University of Enna, Italy. ^342^Department of Neuroscience, SUNY Upstate Medical University, Syracuse, NY, USA. ^343^Faculty of Medicine, Department of Psychiatry, School of Health Sciences, University of Iceland, Reykjavik, IS. ^344^Psychiatry and the Behavioral Sciences, University of Southern California, Los Angeles, CA, US. ^345^Mood Disorders, PsyQ, Rotterdam, NL. ^346^Faculty of Medicine, University of Iceland, Reykjavik, IS. ^347^Department of Genetics, University of North Carolina at Chapel Hill, Chapel Hill, NC, USA. ^348^Department of Environmental Epidemiology, Nofer Institute of Occupational Medicine, Lodz, Poland. ^349^Centro de Biología Molecular Severo Ochoa, Universidad Autónoma de Madrid & CSIC, Madrid, Spain. ^350^Department of Psychiatry, McGill University, Montreal, Canada. ^351^Dept of Psychiatry, Sankt Olavs Hospital Universitetssykehuset i Trondheim, Trondheim, NO. ^352^Department of Psychiatry and Human Behavior, School of Medicine, University of California, Irvine, CA, USA. ^353^Psychiatry, Psychiatrisches Zentrum Nordbaden, Wiesloch, DE. ^354^Clinical Institute of Neuroscience, Hospital Clinic, University of Barcelona, IDIBAPS, CIBERSAM, Barcelona, ES. ^355^Brain Molecular Science, Centre for Addiction & Mental Health, Toronto, Ontario, Canada. ^356^Department of Psychology, Emory University, Atlanta, Georgia, USA. ^357^Center for GeoGenetics, GLOBE Institute, University of Copenhagen, Copenhagen, Denmark. ^358^Institute of Biological Psychiatry, Mental Health Services, Copenhagen University Hospital, Copenhagen, Denmark. ^359^Samsung Advanced Institute for Health Sciences and Technology (SAIHST), Sungkyunkwan University, Samsung Medical Center, Seoul, South Korea. ^360^Queensland Brain Institute, The University of Queensland, Brisbane, QLD, AU. ^361^Computational Sciences Center of Emphasis, Pfizer Global Research and Development, Cambridge, MA, US. ^362^Dalla Lana School of Public Health, University of Toronto, Toronto, ON, CA. ^363^Department of Psychological Medicine, Institute of Psychiatry, Psychology and Neuroscience, King’s College London, London, GB. ^364^South London and Maudsley NHS Foundation Trust, Bethlem Royal Hospital, Monks Orchard Road, Beckenham, Kent, GB. ^365^Section of Biomedical Informatics and Data Science, Yale School of Medicine, New Haven, CT, USA. ^366^Dept of Translational Neuroscience, UMC Utrecht, Netherlands. ^367^Rintveld Eating disorder clinic, Altrecht GGZ, Zeist, Netherlands. ^368^Department of Nutrition, University of North Carolina at Chapel Hill, Chapel Hill, NC, USA. ^369^l’institut du thorax, INSERM, CNRS, Nantes Université. ^370^Department of Psychiatric Genetics, Department of Psychiatry, University of Medical Sciences, Poznan, Poland. ^371^Department of Human Genetics, University Hospital of Liège, Liège, Belgium. ^372^Rheumatology Department, University Hospital of Liège, Liège, Belgium. ^373^Ciber Physiopathology of Obesity and Nutrition (CIBERObn), Instituto de Salud Carlos III, Spain. ^374^Department of Clinical Psychology, University Hospital of Bellvitge-IDIBELL, Barcelona, Spain. ^375^Department of Clinical Sciences, Shool of Medicine and Health Sciences, University of Barcelona, Spain. ^376^Psychoneurobiology of Eating and Addictive Behaviors Group, Neuroscience Program, Bellvitge Biomedical Research Institute (IDIBELL), L’Hospitalet de Llobregat, Spain. ^377^Center for Excellence in Eating Disorders Tübingen (KOMET). ^378^German Center for Mental Health (DZPG). ^379^Medical University Hospital Tübingen, Dpt Psychosomatic Medicine & Psychotherapy. ^380^Université Paris Cité, Institute of Psychiatry and Neuroscience of Paris (INSERM U1266), Paris, France. ^381^Department of Psychiatry, McLean Hospital, Harvard Medical School, Belmont, MA, USA. ^382^Department of Psychological Medicine, University of Otago, Christchurch, New Zealand. ^383^Te Whatu Ora -Waitaha (Health New Zealand), Christchurch, New Zealand. ^384^Institute of Public Health and Clinical Nutrition, University of Eastern Finland, Kuopio, Finland. ^385^Groningen Institute for Evolutionary Life Sciences, University of Groningen, Groningen, The Netherlands. ^386^Department of Pathology and Biomedical Science, University of Otago, Christchurch, New Zealand. ^387^Department of Clinical Psychology, College of Professional Psychology, The Chicago School, Washington DC. ^388^Department of Psychiatry, University of Campania L. Vanvitelli, Naples, Italy. ^389^Department of Community, Family, and Addiction Sciences, Texas Tech University, Lubbock, Texas, USA. ^390^Department of Neuroscience, Psychology, Drug Research and Child Health University of Florence, Italy. ^391^IRCCS Fondazione Don Carlo Gnocchi, Florence, Italy. ^392^Department of Psychiatry, Genetics and Genomic Sciences, Icahn School of Medicine at Mount Sinai, New York, NY. ^393^Department of Psychiatry, Division of Adolescent Psychiatry, Helsinki University Hospital, Helsinki, Finland. ^394^University of Helsinki, Clinicum, Faculty of Medicine. ^395^Department of Health Science, University of Florence, Firenze, Italy. ^396^Department of General Practice & Primary Healthcare, Faculty of Medical & Health Sciences, University of Auckland, New Zealand. ^397^Department of Adult Psychiatry, University of Medical Sciences, Poznan, Poland. ^398^Department of Psychological Medicine, Centre for Research in Eating and Weight Disorders, King’s College London, UK. ^399^Medical University Vienna, Department of Psychiatry and Psychotherapy, Austria. ^400^Sigmund Freud University Vienna, Faculty of Medicine, Austria. ^401^Department of Psychiatry, School of Medicine, The University of North Carolina at Chapel Hill, United States. ^402^Discipline of Psychology, School of Population Health, Curtin University, Perth, Australia. ^403^Division of Paediatrics, School of Medicine, The University of Western Australia, Perth, Australia. ^404^Centre of Excellence for Eating Disorders Tuebingen Germany (KOMET). ^405^Department of Psychosomatic Medicine and Psychotherapy, University Medical Hospital University Tuebingen, Germany. ^406^German Centre for Mental Health (DZPG) Tuebingen, Germany. ^407^Copenhagen Research Centre for Biological and Precision Psychiatry, Mental Health Centre Copenhagen, Copenhagen University Hospital. ^408^Department of Psychiatry, Universidade Federal de Sao Paulo. ^409^Child Health Research Centre, The University of Queensland, Brisbane, Australia. ^410^Department of Psychiatry and Medical Psychology, Universidade Federal de São Paulo (UNIFESP), São Paulo, Brazil. ^411^Department of Psychiatry and Psychotherapy, University Medicine Greifswald, Greifswald, Germany. ^412^Department of Psychiatry, Stanford University, 401 Quarry Rd., Stanford, CA 94305-5797, USA. ^413^Department of Biological Sciences, Purdue University, 915 Mitch Daniels Boulevard, West Lafayette, IN 47906. ^414^Departments of Psychiatry and Human & Molecular Genetics, Virginia Commonwealth University, Richmond, VA, USA. ^415^Child Mind Institute, NY. ^416^Department of Psychiatry, Universidade Federal do Rio Grande do Sul - Porto Alegre, Brazil. ^417^Hospital de Clinicas de Porto Alegre, Porto Alegre - Brazil. ^418^National Institute of Developmental Psychiatry, Sao Paulo - Brazil. ^419^Department of Biochemistry, Universidade Federal de Sao Paulo, Brasil. ^420^GenOmics and Translational Research Center, RTI International, Durham NC, USA. ^421^Department of Medicine, University of Wisconsin School of Medicine and Public Health. ^422^Department of Psychiatry, Robert Wood Johnson Medical School, Rutgers—The State University of New Jersey, New Brunswick, NJ 08901, USA. ^423^Department of Psychiatry, Rutgers Robert Wood Johnson School of Medicine, Rutgers University, Piscataway, NJ 08854. ^424^Drichel Analytics, Bonn, Germany. ^425^Department of Medicine (Biomedical Genetics), Boston University School of Medicine, Boston, MA, 02118, USA. ^426^RTI International, Research Triangle Park, NC. ^427^Department of Epidemiology, University of Colorado Anschutz Medical Campus, Aurora, CO, 80045, USA. ^428^Fellow Program, RTI International, Durham NC, USA. ^429^Center for Craniofacial and Dental Genetics, Department of Oral and Craniofacial Sciences, School of Dental Medicine, University of Pittsburgh, Pittsburgh, PA 15219. ^430^Department of Community Dentistry and Behavioral Science, University of Florida, Gainesville, FL 32608, USA. ^431^Cologne Center for Genomics, University of Cologne, Cologne, Germany. ^432^Department of Genetics, Washington University School of Medicine, St. Louis, MO, 63110, USA. ^433^Division of Biostatistics, Washington University School of Medicine, Saint Louis, MO 63110. ^434^National Human Genome Research Institute, National Institutes of Health, Bethesda, MD, 20892, USA. ^435^Biomedical Genetics, Boston University Chobanian & Avedisian School of Medicine, Boston, MA 02131. ^436^Department of Psychology, University of Minnesota Twin Cities, Minneapolis, MN, 55455, USA. ^437^Biobot Analytics, Cambridge, MA, 02139, USA. ^438^PI Pharmaimage Biomarker Solutions GmbH, Cambridge, MA, 02142, USA. ^439^Department of Molecular Biology, Semmelweis University, Budapest, Hungary. ^440^Duke University Hospital, Department of Surgery, Durham, NC. ^441^Department of Pharmacogenetics, Instituto Nacional de Psiquiatría Ramon de la Fuente Muñiz. ^442^Department of psychiatry, UMCG & RUG. ^443^GGZ Drenthe, the Netherlands. ^444^Accare Child Study Center, Groningen, Netherlands. ^445^University of Groningen, University Medical Center Groningen, Department of Child and Adolescent Psychiatry, Groningen, Netherlands. ^446^Child Study Center and Department of Psychiatry, Yale School of Medicine, New Haven, CT, USA. ^447^Department of Child and Adolescent Psychiatry and Psychotherapy, Psychiatric University Hospital Zurich, University of Zurich, Zurich, Switzerland. ^448^Neuroscience Center Zurich, University of Zurich and the ETH Zurich, Zurich, Switzerland. ^449^Zurich Center for Integrative Human Physiology, University of Zurich, Switzerland. ^450^Bergen Center for Brain Plasticity (BCBP), Haukeland University Hospital, Bergen, Norway. ^451^Helse Møre and Romsdal Hospital Trust, Department of Psychiatry, Molde, Norway. ^452^Norwegian University for Science and Technology, Trondheim, Norway. ^453^Department of Psychiatry, University of Michigan, Ann Arbor, Michigan, USA. ^454^Centre for Crisis Psychology, University of Bergen, Norway. ^455^Rutgers, the State University of New Jersey, Piscataway, NJ, 08854. ^456^Department of Medicine, MSB Medical School Berlin, Berlin, Germany. ^457^Department of Clinical Psychology, University of Bergen, Norway. ^458^Carracci Medical Group, Mexico City, Mexico. ^459^Department of Clinical Science, University of Bergen, Bergen, Norway. ^460^University of California, Irvine - Dept of Psychiatry. ^461^Department of Mental Health, Johns Hopkins Bloomberg School of Public Health, Baltimore, MD, 21205, USA. ^462^Department of Neurology, Norman Fixel Institute for Neurological Diseases, Gainesville FL. ^463^Department of Clinical Sciences, Lund University, Lund, Sweden. ^464^Alpert Medical School of Brown University, Providence, RI, USA. ^465^Butler Hospital, Providence, RI, USA. ^466^Faculdade de Medicina, Universidade de São Paulo. ^467^Laboratory of Genomics of Psychiatric and Neurodegenerative Diseases, National Institute of Genomic Medicine (INMEGEN), Mexico City, Mexico. ^468^Department of Mental Health, Greater Los Angeles VA Healthcare, Los Angeles, CA. ^469^Department of Psychiatry and Biobehavioral Sciences, University of California at Los Angeles, Los Angeles, CA. ^470^Centro de Investigación Biomédica en Red de Enfermedades Raras (CIBERER), ISCIII, Madrid, Spain. ^471^Institut de Biomedicina de la Universitat de Barcelona (IBUB), Catalonia, Spain. ^472^Institut de Recerca Sant Joan de Déu (IRSJD), Esplugues de Llobregat, Catalonia, Spain. ^473^Cologne Excellence Cluster for Stress Responses in Ageing-Associated Diseases (CECAD), University of Cologne, Cologne, Germany. ^474^Department of Neurodegenerative Diseases and Geriatric Psychiatry, University Hospital Bonn, Bonn, Germany. ^475^Department of Psychiatry and Glenn Biggs Institute for Alzheimer’s and Neurodegenerative Diseases, San Antonio, TX, USA. ^476^Division of Neurogenetics and Molecular Psychiatry, Department of Psychiatry and Psychotherapy, University of Cologne, Medical Faculty, Cologne, Germany. ^477^German Center for Neurodegenerative Diseases (DZNE), Bonn, Germany. ^478^Department of Clinical and experimental Medicine, Child and Adolescent Neuropsichiatry, Catania University, Italy. ^479^Genomics and Bioinformatics, Center for Research in Molecular Medicine and Chronic Diseases (CiMUS), University of Santiago de Compostela, Spain. ^480^Department of Neurology, Massachusetts General Hospital, Harvard Medical School, Boston, MA, USA. ^481^Johns Hopkins University School of Medicine, Department of Neurology and the Kennedy Krieger Institute, Baltimore, MD, USA. ^482^Department of Psychiatry and Behavioral Sciences, Baylor College of Medicine, Houston, TX, USA. ^483^Departments of Psychiatry and Pediatrics, Columbia University, New York, NY, USA. ^484^New York State Psychiatric Institute, New York, NY, USA. ^485^Imaging Future Aps, Denmark. ^486^Biogen. ^487^Department of Psychiatry and Behavioral Sciences, University of Minnesota Medical School. ^488^Minneapolis VA Health Care System. ^489^Case Western Reserve University. ^490^Department of Psychiatry and Behavioral Neurosciences, Wayne State University School of Medicine. ^491^Cohen Veterans Bioscience. ^492^Department of Psychiatry, Boston University Chobanian & Avedisian School of Medicine, Boston, MA 02118, USA. ^493^Million Veteran Program (MVP) Coordinating Center, VA Boston Healthcare System, Boston, MA 02130, USA. ^494^Department of Psychiatry, University of Connecticut School of Medicine, Farmington, CT. ^495^Department of Gynecology and Obstetrics, Emory University, Atlanta, GA. ^496^Department of Psychiatry and Behavioral Sciences, Duke University School of Medicine, Durham, NC, USA. ^497^Durham Veterans Affairs (VA) Health Care System, Durham, NC, USA. ^498^VA Mid-Atlantic Mental Illness Research, Education and Clinical Center, Durham, NC, USA. ^499^Department of Epidemiology, Harvard TH Chan School of Public Health, Boston MA USA. ^500^Department of Psychiatry, State University of New York Downstate Medical Center, Brooklyn, NY. ^501^Connecticut VA Healthcare Center, Orange, CT, USA. ^502^Department of Psychological & Brain Sciences, Indiana University, Bloomington IN 47405. ^503^South African Medical Research Council Genomics of Brain Disorders Research Unit, Department of Psychiatry, Faculty of Medicine and Health Sciences, Stellenbosch University, Cape Town, South Africa. ^504^Department of Human Genetics, Emory University, Atlanta, GA. ^505^Department of Pharmaceutical Sciences, University of Pittsburgh. ^506^Mental Health Nuroscience Research Department, Division of Psychiatry. ^507^Centre Hospitalier du Rouvray, Rouen 76000 France. ^508^INSERM U1245, 76000 Rouen, Normandie, France. ^509^Centre de Référence des Maladies Rares à Expression Psychiatrique, Department of Child and Adolescent Psychiatry, AP-HP Sorbonne Université, Hôpital Universitaire de la Pitié-Salpêtrière, 47 - 83 Boulevard de l’Hôpital, 75651 Paris Cedex 13, France. ^510^Faculté de Médecine Sorbonne Université, Groupe de Recherche Clinique n°15 - Troubles Psychiatriques et Développement (PSYDEV), Department of Child and Adolescent Psychiatry, Hôpital Universitaire de la Pitié-Salpêtrière, 47-83 Boulevard de l’Hôpital, 75651 Paris Cedex 13, France. ^511^Institut des Systèmes Intelligents et de Robotique (ISIR), CNRS UMR7222, Sorbonne Université, Campus Pierre et Marie Curie, Faculté des Sciences et Ingénierie, Pyramide, Tour 55, Boîte courrier 173, 4 Place Jussieu, 75252 Paris Cedex 05, France. ^512^Department of general practice and primary health care. University of Helsinki, Finland. ^513^Department of obstetrics and gynecology, National University of Singapore, Singapore. ^514^Mental Health Research Institute, Tomsk National Research Medical Center, Tomsk, Russia. ^515^School of Non-Destructive Testing, Tomsk Polytechnic University, Tomsk, Russia. ^516^Department of Psychiatry, University of Colorado Denver School of Medicine, Aurora, CO, USA. ^517^Institute of Translational Biomedicine, Saint Petersburg State University, Saint Petersburg, Russia. ^518^Saint Petersburg University Hospital, Saint Petersburg State University, Saint Petersburg, Russia. ^519^Comprehensive Center for Clinical Neurosciences and Mental Health (C3NMH), Medical University of Vienna, Austria. ^520^Departments of Psychiatry and Human and Molecular Genetics, INSERM, Institut de Myologie, Hôpital de la Pitiè-Salpêtrière, Paris, 75013, France. ^521^UFR santé, Université de Rouen Normandie, Rouen, France. ^522^Department of Psychiatry and Psychotherapy, Comprehensive Center for Clinical Neurosciences and Mental Health (C3NMH), Medical University of Vienna, Austria. ^523^Psychiatry, Addictology and Psychotherapy Department, Siberian State Medical University, Tomsk, Russia. ^524^Department of Psychiatric Genomics, Bekhterev National Medical Research Center for Psychiatry and Neurology, Saint Petersburg, Russia. ^525^Valdman Institute of Pharmacology, First St.-Petersburg Pavlov State Medical University, Saint Petersburg, Russia. ^526^Saint Petersburg State University. ^527^Dept Psychosis Studies, Institute of Psychiatry, Psychology and Neuroscience, King’s College, London. ^528^Généthon, 1 bis, Rue de l’Internationale, 91000 Evry, France. ^529^Faculty of Science, Medicine and Health, School of Chemistry and Molecular Bioscience, University of Wollongong, Wollongong NSW 2522, Australia. ^530^Illawarra Health and Medical Research Institute, Wollongong NSW 2522, Australia. ^531^INSERM U1266, Institut de psychiatrie et de neurosciences, Paris, France. ^532^Université de Paris, Faculté de médecine, Hôpital Cochin-Tarnier, Paris 75006, France. ^533^School of Psychiatry and Clinical Neurosciences, The University of Western Australia, Perth WA 6009, Australia. ^534^Department of Psychological & Brain Sciences, Washington University, St. Louis, MO, USA. ^535^Institute of Behavioral Science and Department of Sociology, University of Colorado, Boulder, CO USA. ^536^Department of Molecular Pharmacology and Experimental Theraeputics, Mayo Clinic College of Medicine and Science. ^537^Department of Psychiatry and Psychology, Mayo Clinic College of Medicine and Science. ^538^Department of Psychiatry, VeteransAffairs Connecticut Healthcare Center, West Haven, CT, USA. ^539^Office of the Clinical Director, NIAAA, NIH, Bethesda MD 20892. ^540^Department of Psychiatry, Virginia Commonwealth University School of Medicine, Virginia Institute for Psychiatric and Behavioral Genetics, Richmond, VA, USA. ^541^Office of the Clinical Director and Lab of Neurogenetics, NIAAA, NIH, Rockville, MD 20852. ^542^Department of Psychiatry and Behavioral Sciences, Stanford University School of Medicine, Stanford, CA, USA and Sierra-Pacific Mental Illness Research, Education and Clinical Center (MIRECC), Veterans Affairs Palo Alto Health Care System, Palo Alto, CA, USA. ^543^Department of Sociology and the Carolina Population Center, University of North Carolina at Chapel Hill, Chapel Hill, NC 27516. ^544^The AI for Health Institute, Washington University School of Medicine. ^545^MRC Human Genetics Unit, IGC, University of Edinburgh, Edinburgh, UK EH4 2XU. ^546^Institue for Behavioral Genetics, University of Colorado Boulder. ^547^Department of Molecular, Cellular, and Developmental Biology, University of Colorado, Boulder Colorado, USA. ^548^Vice-Director for Research and Head of the Department of Addictions, Bekhterev National Medical Research Center for Psychiatry and Neurology. ^549^Department of Psychiatry, University of Iowa Carver College of Medicine. ^550^Folkhälsan Research Center. ^551^Centre for Cardiovascular Science, University of Edinburgh, Edinburgh, United Kingdom. ^552^Population Health Unit, Finnish Institute for Health and Welfare, Helsinki and Oulu, Finland. ^553^Institute for Genomic Medicine, University of California San Diego, La Jolla, CA, 92093. ^554^Jackson Laboratory, Bar Harbor, Maine, USA. ^555^Providence VA Medical Center, Providence, Rhode Island, USA. ^556^Department of Medicine, University of Otago, Christchurch, New Zealand. ^557^Department of Psychiatry, State University of New York Downstate Health Science University, Brooklyn, NY. ^558^Department of Psychiatry, Psychotherapie and Psychosomatics, Martin-Luther University Halle-Wittenberg. ^559^RKH Ludwigsburg, Psychiatrie, Psychotherapie und Psychosomatische Medizin. ^560^Department of Medicine, Rutgers New Jersey Medical School, Newark NJ 07103. ^561^Department of Medicine, Division of Genetic Medicine, Vanderbilt University, Nashville, TN, USA. ^562^Department of Psychiatry and Psychotherapy, LVR-University Hospital Essen, University of Duisburg-Essen, Germany. ^563^Departments of Pharmaceutical Sciences, Psychiatry, and Human Genetics, University of Pittsburgh. ^564^Department of Sociology, Purdue University. ^565^Department of Psychiatry and Psychotherapy, Center of Addiction Medicine, University Hospital Regensburg at the Bezirksklinikum, Regensburg, Germany. ^566^Epidemiology and Biometry Branch, National Institute on Alcohol Abuse and Alcoholism, National Institutes of Health, Bethesda, MD 20892. ^567^Department of Biostatistics, Yale School of Public Health, New Haven, CT 06520. ^568^Departments of Psychiatry and Physiology, University of Toronto, Toronto, ON, Canada. ^569^Division of Experimental and Translational Neuroscience, Krembil Research Institute, University Health Network, Toronto, ON, Canada. ^570^Department of Psychiatry, Socialpsychiatry and Psychotherapy, Hannover

