## Supplementary Note for "The Landscape of Shared and Divergent Genetic Influences across 14 Psychiatric Disorders"

**Table of Contents**

***Follow-up Analyses: Freeze 3 PTSD and MD	3***

***Univariate MiXeR	3***

***PheWAS in Mayo Clinic Participant Sample	4***

***MAGMA Enrichment	5***

***Supplementary Figure 1. Participant Sample Sizes for CDG2 vs CDG3	6***

***Supplementary Figure 2. Genetic Correlations and Heritabilities across Data Freezes for MD and PTSD	7***

***Supplementary Figure 3. Local genetic correlation patterns for MD and PTSD	8***

***Supplementary Figure 4. LAVA PTSD and MD Venn Diagram	9***

***Supplementary Figure 5. Univariate MiXeR Results	10***

***Supplementary Figure 6. Cross-trait MiXeR results for Schizophrenia and Bipolar disorder	11***

***Supplementary Figure 7. Cross-trait MiXeR results for Major Depressive Disorder and Anxiety Disorders	12***

***Supplementary Figure 8. Cross-trait MiXeR results for Post-traumatic Stress Disorder and Attention-deficit/hyperactivity Disorder	13***

***Supplementary Figure 9. Cross-trait MiXeR results for Alcohol Use Disorder and Anorexia Nervosa	14***

***Supplementary Figure 10. Concordance between global and local genetic correlations	15***

***Supplementary Figure 11. Miami plots of local rg for disorder pairs that exhibited significant negative local rg	16***

***Supplementary Figure 12a. Miami plots of local rg for disorder pairs with significant rg correlation in top hotspot locus on chromosome 11	17***

***Supplementary Figure 12b. Miami plots of local rg for disorder pairs with significant rg correlation in top hotspot locus on chromosome 11	18***

***Supplementary Figure 12c. Miami plots of local rg for disorder pairs with significant rg correlation in top hotspot locus on chromosome 11	19***

***Supplementary Figure 13. Multivariate GWAS Miami Plots.	20***

***Supplementary Figure 14. Factor QQ-Plots.	21***

***Supplementary Figure 15. Factor 5 QQ-Plot excluding top QSNP hits	22***

***Supplementary Figure 16a. PheWAS results for the Compulsive Disorders Factor	23***

***Supplementary Figure 16b. PheWAS results for the Schizophrenia and Bipolar (SB) Factor	24***

***Supplementary Figure 16c. PheWAS results for the Neurodevelopmental Disorders Factor.	25***

***Supplementary Figure 16d. PheWAS results for the Internalizing Disorders Factor	26***

***Supplementary Figure 16e. PheWAS results for the Substance Use Disorders Factor	27***

***Supplementary Figure 16f. PheWAS results for the Hierarchical p-Factor	28***

***Supplementary Figure 17. Predicting target genes of CDG variants and temporal expression pattern along developmental trajectory in human developing brain	29***

***Supplementary Figure 18. Enrichment of CDG variants near risk genes of related disorders	30***

***Supplementary Figure 19. Cell type specificity of CDG variants target genes	31***

***References for Online Supplement	32***

### Follow-up Analyses: Freeze 3 PTSD and MD

One surprising result was the LDSC estimated genetic correlation of .99 (*SE* = .03) across the Freeze 3 iterations of the PTSD and MD GWAS summary statistics. We ran several follow-up analyses to try and better understand this particular result. First, we note that the attenuation ratio for the HapMap3 SNPs used by LDSC—calculated as (univariate LDSC Intercept – 1) / (mean $\chi^{2}$-1)—was similar and within a reasonable range for both PTSD (.041) and MD (.048), suggesting that neither disorder might be characterized by uncontrolled confounding that is driving the effects. Second, we examined the SNP-based heritabilities and genetic correlations across the three freezes of PTSD and MD (**Suppl. Fig. 2**). We find that, as would be expected given smaller sample sizes, estimates of this genetic correlation using earlier freezes were imprecise. At the same time, the 95% confidence intervals on these estimates did not include the current estimate of .99. This tentatively indicates that there has been a shift in the underlying population estimate, as opposed to a gradual homing in on a more precise estimate of the same underlying population.

To investigate the possibility of cohort-level confounding, we obtained a version of the summary statistics for PTSD excluding the largest cohort added in Freeze 3: the Million Veterans Program (MVP). We find that the inclusion of MVP is highly unlikely to be a cause for the upward trend in genetic correlation, as PTSD Freeze 3 excluding MVP was estimated to have a comparable genetic correlation to MD Freeze 3 (*r_g_* = 1.02, *SE* = .04). We also examined the relationship between PTSD Freeze 3 and summary statistics from Coleman et al. (2021)^1^ that examined GWAS estimates for MD for individuals with and without self-reported trauma exposure in the UK Biobank. These results revealed a similar level of genetic overlap across PTSD Freeze 3 and both MD with (*r_g_* = .72, *SE* = .07) and without trauma exposure (*r_g_* = .70, *SE* = .06). This suggests results are not being driven by MD cohorts, for which cases are more likely to have been exposed to trauma (e.g., veterans-based cohorts). We then examined results when pruning the MD Freeze 3 summary statistics with heterogeneity *I^2^* statistic *p*-values < 1$\times$10^-5^. These pruned MD summary statistics were also estimated to have a .99 *r_g_* with PTSD. However, it is important to note that LAVA estimated an average local *r_g_* of .6, with several loci that are uniquely associated with only one of the two disorders (**Suppl. Figs. 3-4**).

It is unclear, based on these results, what is driving the general increase in genetic correlation across PTSD and MD across freezes. Given decreasing SNP-based heritability estimates across the freezes for both disorders, it may be that with increasing numbers of contributing cohorts to the univariate GWAS meta-analysis that the psychiatric signal is being reduced to more transdiagnostic components (**Suppl. Fig. 2**). As the explicit purpose of CDG3 was to triangulate across methods, we considered the framework of this paper an opportunity to identify what is shared and unique across even the highly overlapping freezes of these two disorders. It will be of critical import for future work to further disentangle what may be driving this estimate with respect to particular cohorts, ascertainment strategies, and shared biology.

### Univariate MiXeR

We applied univariate MiXeR^2^ to estimate polygenicity, measuring the effective number of causal variants for each phenotype. Formally, polygenicity was defined as the number of causal variants that are estimated to account for 90% of a trait's SNP heritability. The estimates of a trait’s polygenicity and discoverability allow one to project the sample size a future GWAS study would need to saturate the yield of genome-wide significant loci (i.e., where genome-wide significant loci are anticipated explain 90% of the trait's heritability). Such “power curves” are shown on **Suppl.** **Fig. 5**. The results are presented in Suppl. Table 5, with SCZ, CUD and TS were among the most “discoverable”, with around 12M subjects needed to saturate the GWAS yield. MD, ANX and PTSD were the least discoverable, with estimated 80M subjects needed to saturate the GWAS yield. We note that in terms of polygenic prediction saturation may happen at a lower N, as in practice the best performing polygenic risk scores are not constrained to genome-wide significant variants. Also, more advanced PRS methods incorporating better priors, continuous shrinkage, and leveraging LD from cross-ancestry GWAS are likely to bring major improvements, and therefore much smaller Ns may be sufficient to reach saturation.

High discoverability in SCZ and TS can be largely attributed to their substantially high SNP heritability, with MiXeR’s estimate approaching 50% on the observed scale. For comparison, observed-scale SNP heritability for ANX, PTSD and MD was between 4% and 8%. Most other disorders were in the “middle” range, with heritability between 20% and 30%.

For polygenicity, SCZ and BIP have similar polygenicity (12k and 11k variants, respectively), while MD has the highest polygenicity and ADHD the lowest polygenicity. It is worth stressing that, while there is some concordance between heritability and polygenicity estimates, they are not perfectly correlated. For example MD has higher polygenicity than SCZ, but SCZ has higher SNP-based heritability.

### PheWAS in Mayo Clinic Participant Sample

Phecodes were ascertained using EHR data from 57,001 patients from the Mayo Clinic Biobank. EHR data for the participants was extracted on September 23, 2022, and included any diagnostics on or before April 6, 2020, the date patient consent was checked. The Institutional Review Board of Mayo Clinic approved this study. All samples were prepared for sequencing using a custom automated sample preparation workflow developed at the Regeneron Genetics Center (RGC). The RGC sequencing assay is a heavily modified version of the TWIST Comprehensive Exome custom capture. The custom design includes additional augmentation of the exome capture with “backbone” regions intended to capture common tagging variation for purposes of genome-wide association studies (GWAS). Specifically, these additional backbone regions are targeted at lower depth, which then undergo substantial post-processing using proprietary algorithms that can boost genotyping quality based on shared information via linkage disequilibrium and population allele frequencies.  This is referred to by RGC as genotyping-by-sequencing (G$\times$S).

The resulting G$\times$S data was run through the Mayo Clinic Genotype QC pipeline. In this QC pipeline, SNPs were excluded using filters for call rate (<95%), minor allele frequency (<0.5%), and deviation from Hardy-Weinberg Equilibrium (*p* < 1$\times$10^-6^). Individuals were excluded for excessive missing data (>5%), sex errors, and abnormal heterozygosity (< 70% on multiple chromosomes). An analysis of genetic ancestry was performed on a subset of 4874 high-quality HapMap3 SNPs. Principal components analysis (PCA) was first performed on the HapMap3 samples, and then MCB samples were projected onto these PCs, with the leading 20 PCs reported. RGC applied a kernel density estimator (KDE) approach to ancestry assignment based on the PCA output. KDEs were trained for each of the individual HapMap3 populations, which were then mapped to one of five main genetic ancestral super-populations (AFR=African; AMR=Admixed American; EAS=East Asian; EUR=European; SAS=South Asian) based on specific likelihood criteria for the individual populations such that the likelihood for a given ancestry group was greater than 0.3, the sample was assigned to that ancestry group. When two ancestry groups had a likelihood of greater than 0.3, RGC arbitrarily assigned AFR over EUR, AMR over EUR, AMR over EAS, SAS over EUR, and AMR over AFR. We restricted all analyses to include only those determined to be of EUR ancestries.

Cryptic relatedness analysis was performed in multiple steps using PLINK^3^ and PRIMUS.^4^ IBD sharing estimates were calculated among all individuals with the same ancestral superclass assignment based on a $\hat{\pi}$ threshold of 0.1875 to capture up to second-degree relationships. A second set of IBD estimates was calculated among all samples based on a $\hat{\pi}$cutoff of 0.3 to capture 1st degree relationships that may span ancestral superclasses. Samples were then grouped into first-degree family networks based on the IBD estimates, which were processed using PRIMUS under default settings.  Finally, samples with cryptic relatedness were removed from the analysis if they had >100 closely related samples ($\hat{\pi}$>0.1875) or >25000 related samples ($\hat{\pi}$>0.08), which was performed iteratively until no such samples remained. For each pair with an estimated 2^nd^ degree or higher relatedness, we removed the individual with shorter length of EHR record. An additional 76 participants were excluded due to overlap with the univariate disorder GWAS for either AUD or BD used in this study.

PRSs were then estimated within this quality-controlled sample using LDpred2-auto^5^ along with the full set (i.e., no *p*-value thresholding) of multivariate GWAS summary statistics for the five factors from the correlated factors model and the transdiagnostic *p-*factor from the hierarchical model. PheWAS Results are reported in **Suppl. Table 30** and displayed for each factor in **Suppl. Fig. 16.** Significance was defined using a Bonferroni corrected threshold of *p* < 4.7$\times$10^-6^, reflecting the standard threshold of *p* < .05 corrected for 10,626 statistical tests. This reflects the product of the number of examined phecodes (1,776) and psychiatric factors (6).

### MAGMA Enrichment

To examine the overlap between our GWAS signal and known disease-causing rare variations, we computed gene-level rare variant heritability using MAGMA.^6^ First, we obtained and harmonized candidate gene lists from contemporary rare variant studies spanning autism spectrum disorder, the broader class of neurodevelopmental disorders, and SCZ.^7–12^ Next, a gene set annotation was generated by pairing each source study with the genes passing significance in that study. This two-column file was fed into MAGMA as a set annotation, with the ‘*--set-annot gene-col=1 set-col=2’* flag, to be compared against gene-level results for the psychiatric factors. To generate continuous annotations, we divided the negative log matrix of gene and study *p*-values into 40 bins, whereby 40 represents a high degree of association between the gene and study and 0 a low one. Studies without *p*-values were assigned a value of 40 for each gene in the study’s curated gene list, and 0 otherwise. This binned gene by study matrix was fed into MAGMA as a covariate file, with the ‘*--gene-covar*’ and ‘*--model direcn=pos*’ flags. Resulting enrichment statistics were Bonferroni corrected for multiple comparisons.

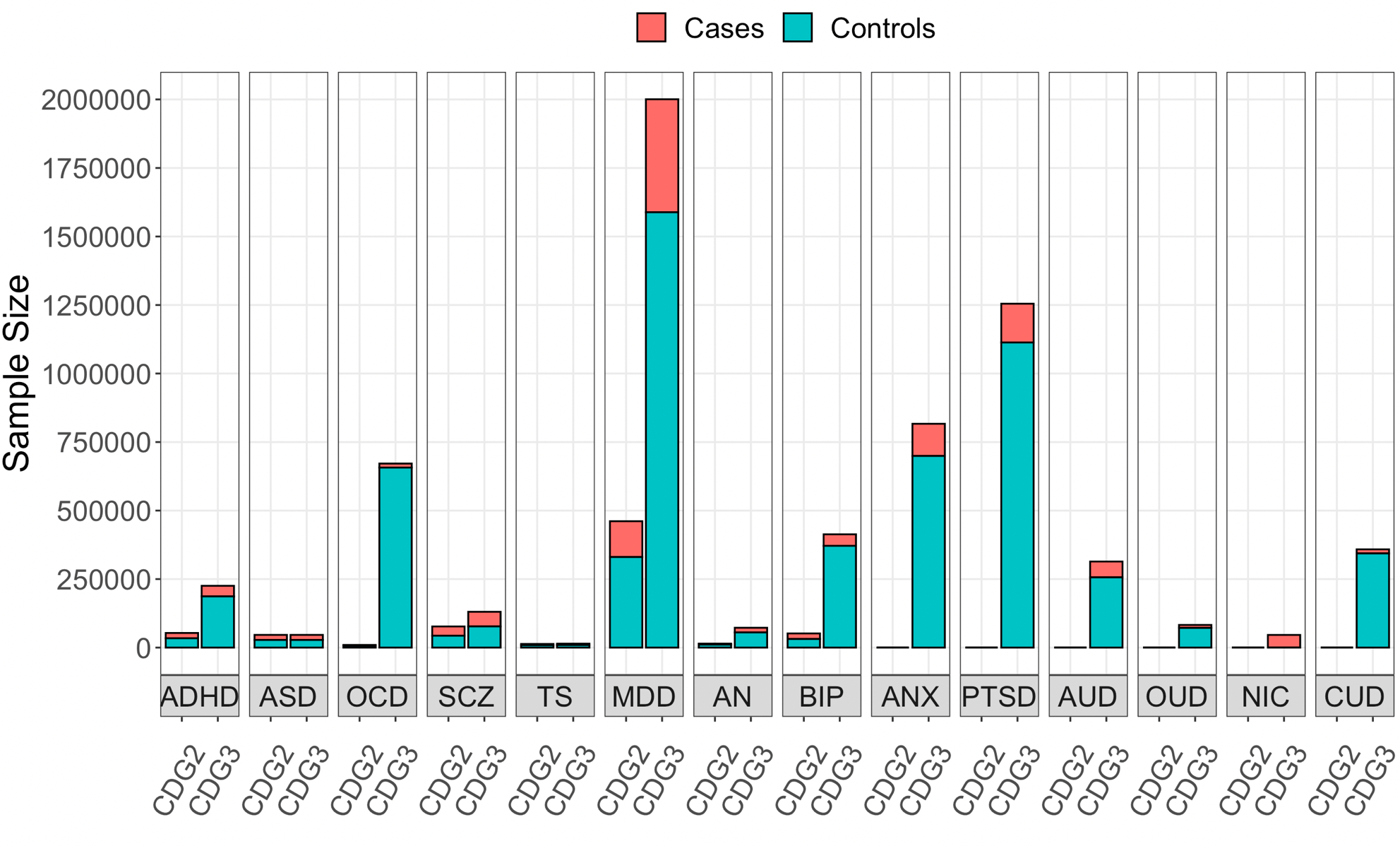

**Supplementary Figure 1. Participant Sample Sizes for CDG2 vs CDG3.** Figure displays the total sample sizes in the CDG2 and CDG3 analyses for the 14 major psychiatric disorders included in the CDG3 analyses. Cases and controls are stacked on top of one another, with controls shown in blue and cases in red. Note that the right-most, six disorders on the x-axis are entirely new relative to the CDG2 analyses. ADHD = attention-deficit hyperactivity disorder; ASD = autism spectrum disorder; OCD = obsessive compulsive disorder; SCZ = schizophrenia; TS = Tourette’s syndrome; MD = major depressive disorder; AN = anorexia nervosa; BIP = bipolar disorder; ANX = anxiety disorder; PTSD = post-traumatic stress disorder; AUD = alcohol use disorder; OUD = opioid use disorder; NIC = nicotine dependence; CUD = cannabis use disorder

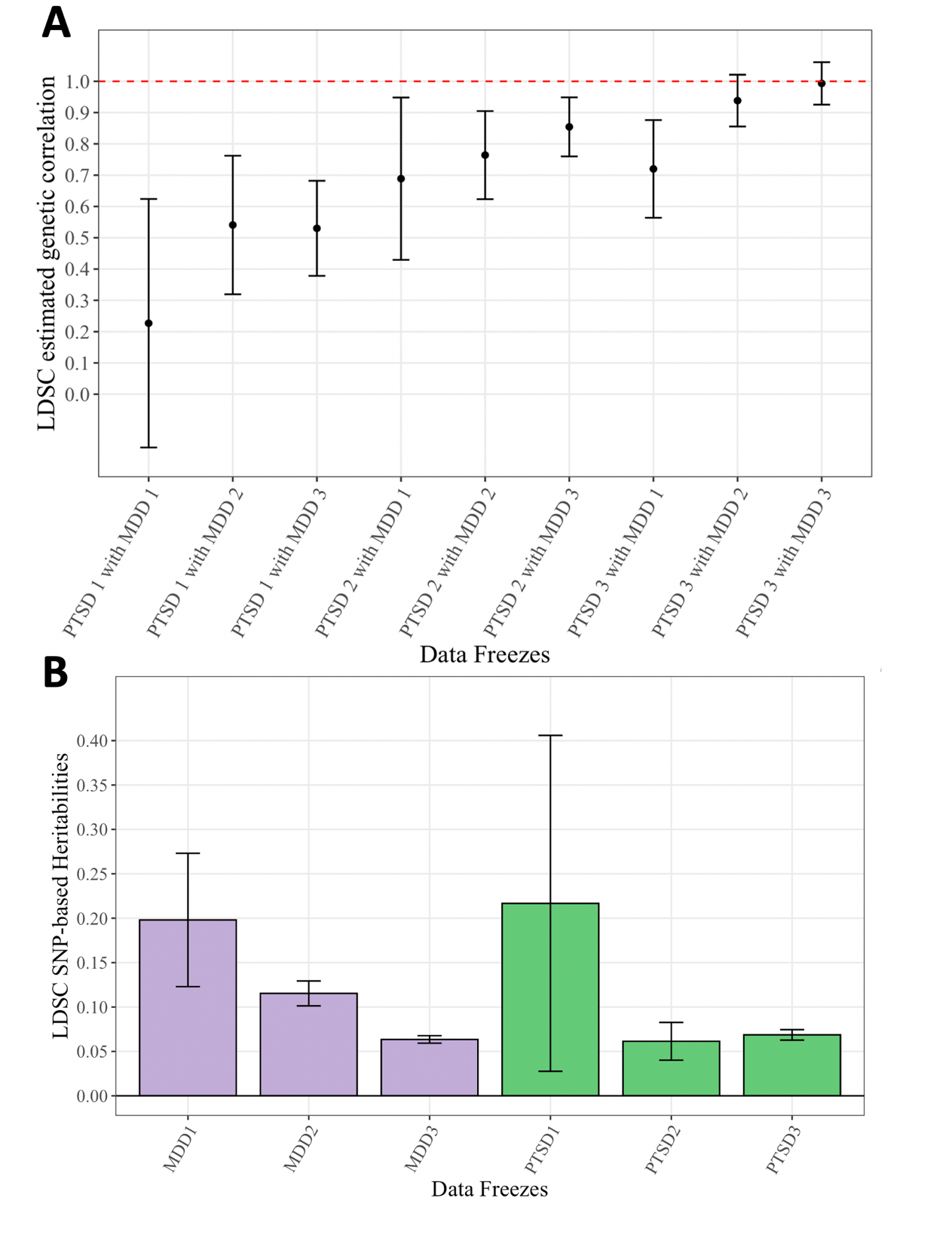

**Supplementary Figure 2. Genetic Correlations and Heritabilities across Data Freezes for MD and PTSD**. *Panel A* depicts the LDSC estimated genetic correlations across data freezes for MD and PTSD. Freezes are depicted sequentially, and as can be readily observed, the genetic correlation across this pair of disorders has been gradually increasing across freezes. *Panel B* depicts the SNP-based heritability estimates from LDSC for these same PTSD and MD data freezes. Error bars depict 95% confidence intervals across both panels.

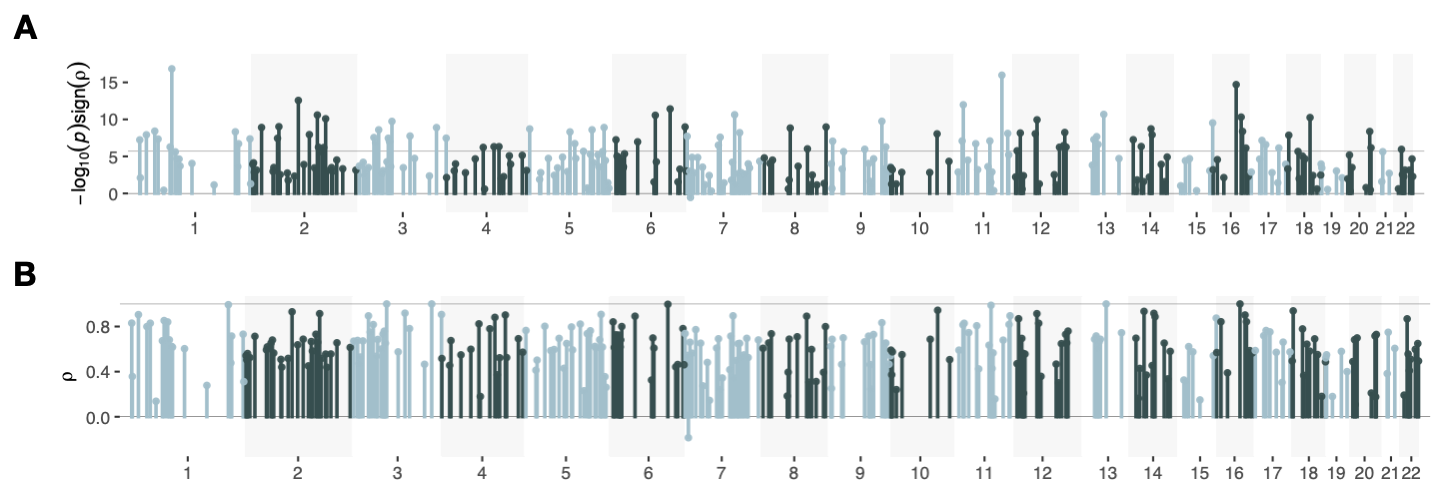

**Supplementary Figure 3. Local genetic correlation patterns for MD and PTSD**. Panel
A shows the -log10 *p*-values for the local genetic correlations scaled by the sign in all evaluated loci (the line indicating the Bonferroni corrected significance threshold of 2.1$\times$10^-6^), while Panel B shows the corresponding local genetic correlation estimate across all evaluated loci (the line indicating an estimate of 1).

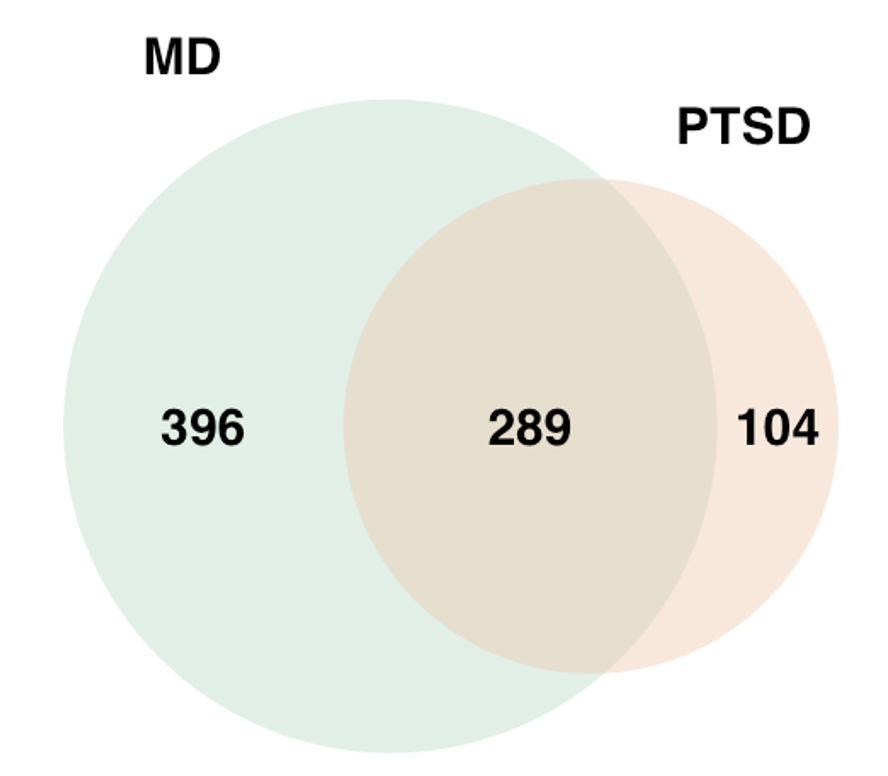

**Supplementary Figure 4. LAVA PTSD and MD Venn Diagram.** Venn diagram illustrating the number of unique and overlapping loci with univariate genetic signal detected at *p* < .05 / 1,093 for MD and PTSD.

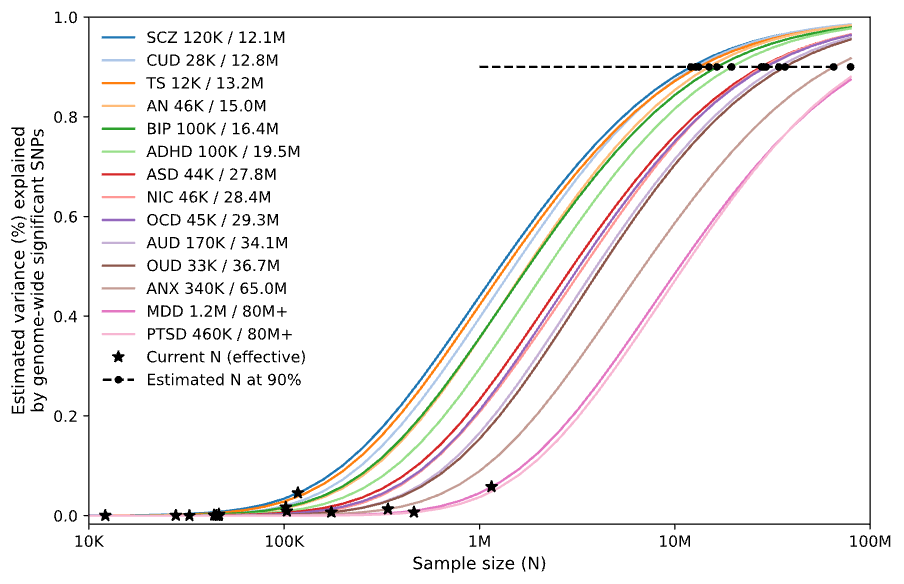

**Supplementary Figure 5. Univariate MiXeR Results.** Power curves estimating the sample size of a GWAS study are needed to saturate the yield of genome-wide significant loci. The legend shows the current effective sample size of today’s GWAS, followed by the projected effective sample size needed for the GWAS yield to saturate.

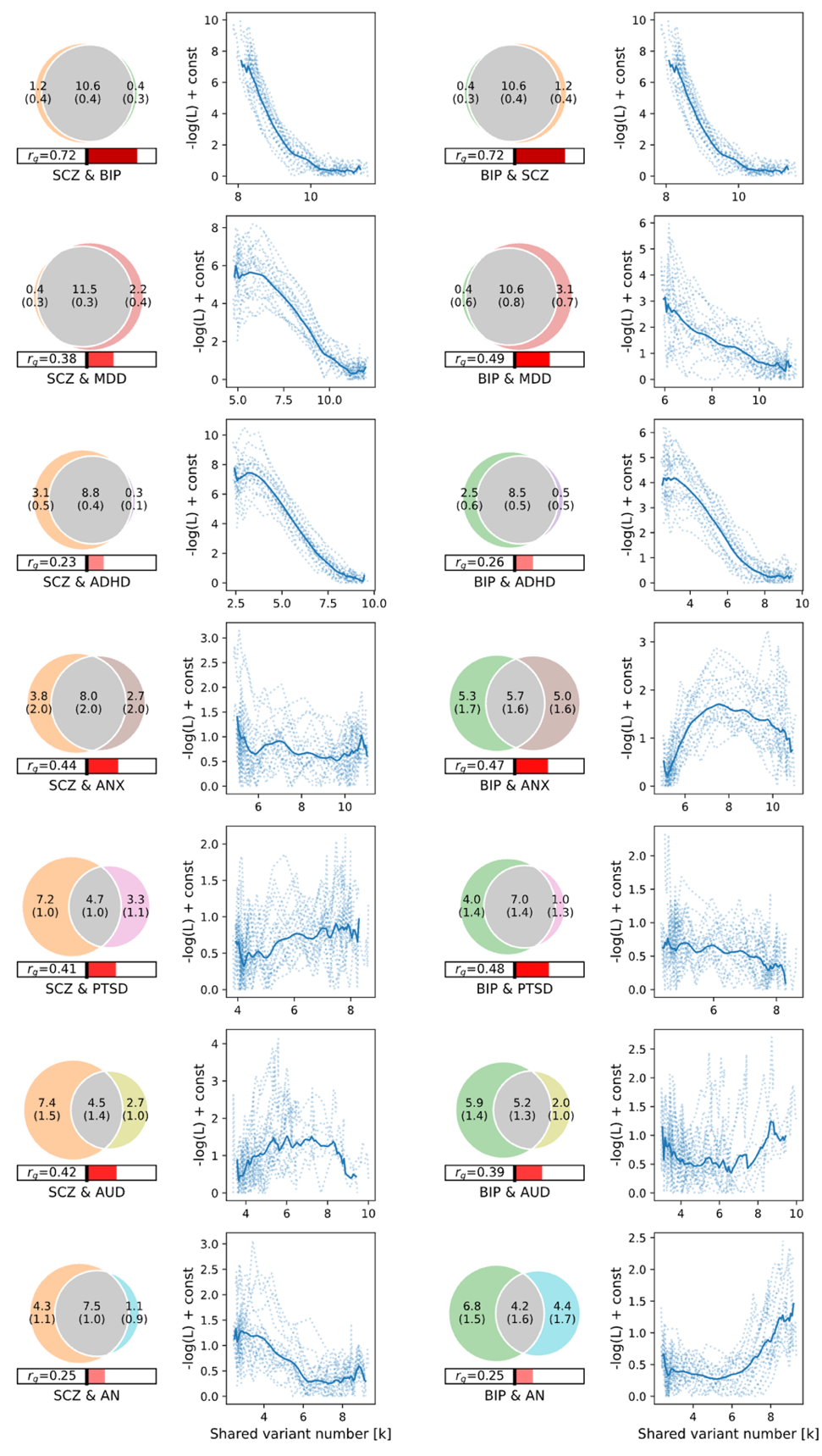

**Supplementary Figure 6. Cross-trait MiXeR results for Schizophrenia and Bipolar disorder.** Venn diagrams of unique and shared polygenic components at the causal level for SCZ (left column) and BIP (right column). The numbers within the Venn diagrams indicate the estimated quantity of causal variants (in thousands) per component, explaining 90% of SNP heritability in each phenotype, followed by the standard error. The size of the circles reflects the degree of polygenicity. Genetic correlation (*r_g_*) is represented in the horizontal bars beneath the Venn diagrams. Right of the central bar (red) indicates positive *r_g_* and left of the central bar (blue) indicates negative *r_g_*. Next to each Venn diagram is a plot of the log-likelihood of the bivariate fit as a function of 𝜋12 (shared variant) parameter. These curves indicate the strength of the evidence of polygenic overlap as compared to minimal overlap. Positive AIC values (**Suppl. Table 5**) and a clear minima on the likelihood curves provide evidence of good model fit.

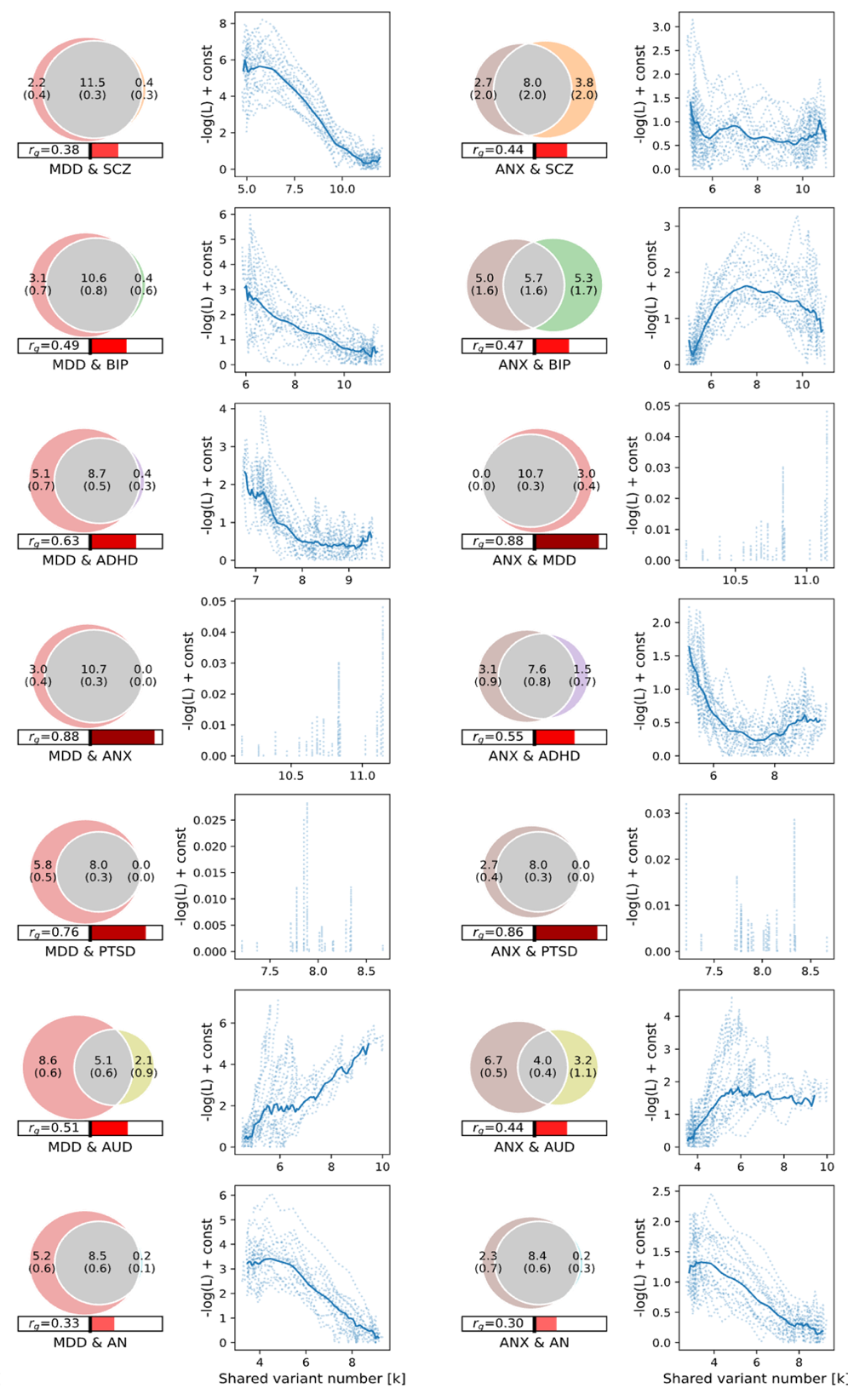

**Supplementary Figure 7. Cross-trait MiXeR results for Major Depressive Disorder and Anxiety Disorders.** Venn diagrams of unique and shared polygenic components at the causal level for MD (left column) and ANX (right column). The numbers within the Venn diagrams indicate the estimated quantity of causal variants (in thousands) per component, explaining 90% of SNP heritability in each phenotype, followed by the standard error. The size of the circles reflects the degree of polygenicity. Genetic correlation (*r_g_*) is represented in the horizontal bars beneath the Venn diagrams. Right of the central bar (red) indicates positive *r_g_* and left of the central bar (blue) indicates negative *r_g_*. Next to each Venn diagram is a plot of the log-likelihood of the bivariate fit as a function of 𝜋12 (shared variant) parameter. These curves indicate the strength of the evidence of polygenic overlap as compared to minimal overlap. Positive AIC values (**Suppl. Table 5**) and a clear minima on the likelihood curves provide evidence of good model fit. We note that for PTSD, ANX and MD the genetic correlations were high to an extent that there was little room for additional overlap beyond correlation, given MiXeR’s modeling assumptions. More specifically, the range in size of the putative shared component is too small to allow for an accurate model fit this situation, as demonstrated by the range on the respective x-axes.

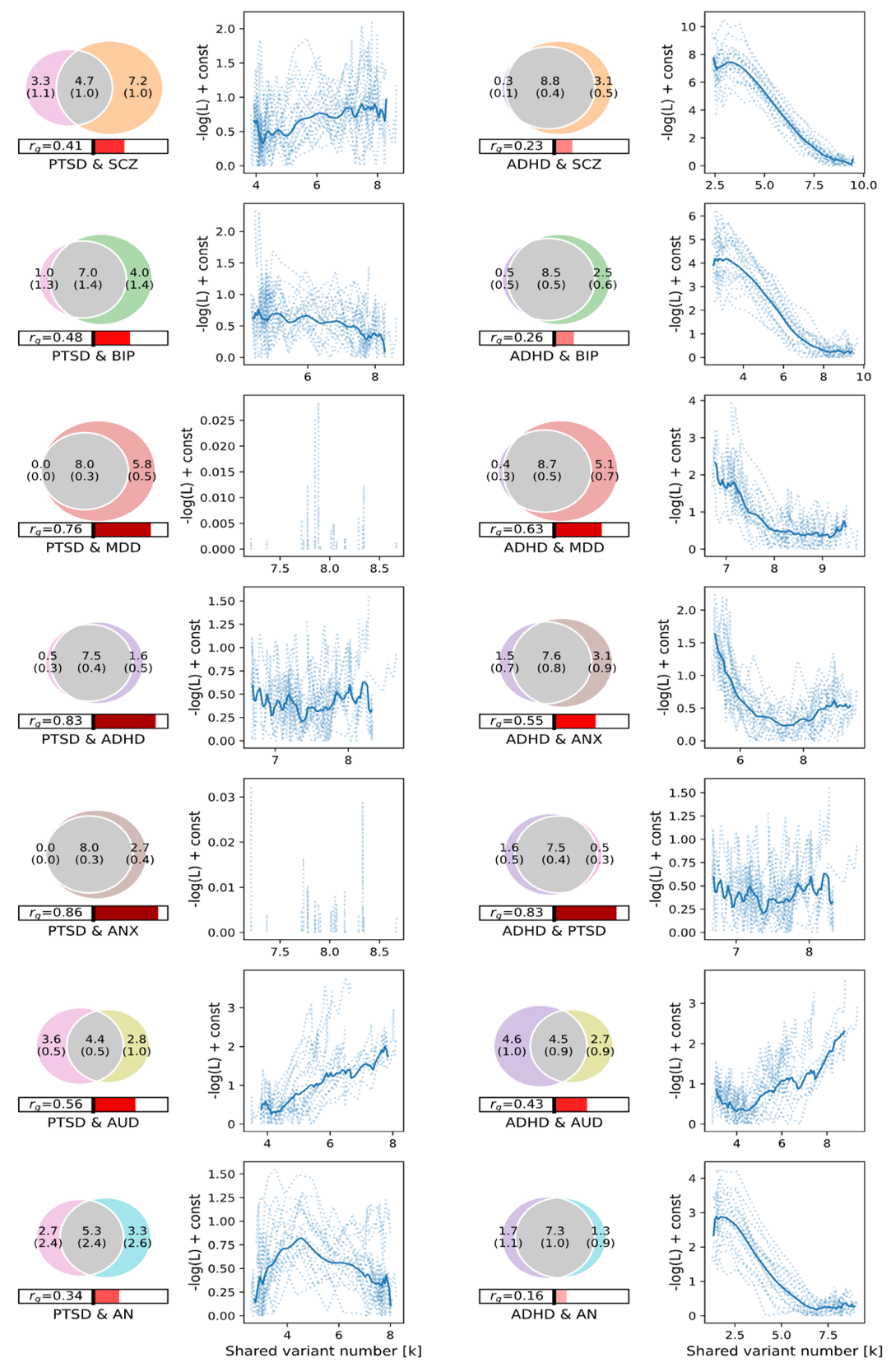

**Supplementary Figure 8. Cross-trait MiXeR results for Post-traumatic Stress Disorder and Attention-deficit/hyperactivity Disorder**. Venn diagrams of unique and shared polygenic components at the causal level for PTSD (left column) and ADHD (right column). The numbers within the Venn diagrams indicate the estimated quantity of causal variants (in thousands) per component, explaining 90% of SNP heritability in each phenotype, followed by the standard error. The size of the circles reflects the degree of polygenicity. Genetic correlation (rg) is represented in the horizontal bars beneath the Venn diagrams. Right of the central bar (red) indicates positive rg and left of the central bar (blue) indicates negative rg. Next to each Venn diagram is a plot of the log-likelihood of the bivariate fit as a function of 𝜋12 (shared variant) parameter. These curves indicate the strength of the evidence of polygenic overlap as compared to minimal overlap. Positive AIC values (Suppl. Table 5) and a clear minima on the likelihood curves provide evidence of good model fit. We note that for PTSD, ANX and MD the genetic correlations were high to an extent that there was little room for additional overlap beyond correlation, given MiXeR’s modeling assumptions. More specifically, the range in size of the putative shared component is too small to allow for an accurate model fit this situation, as demonstrated by the range on the respective x-axes.

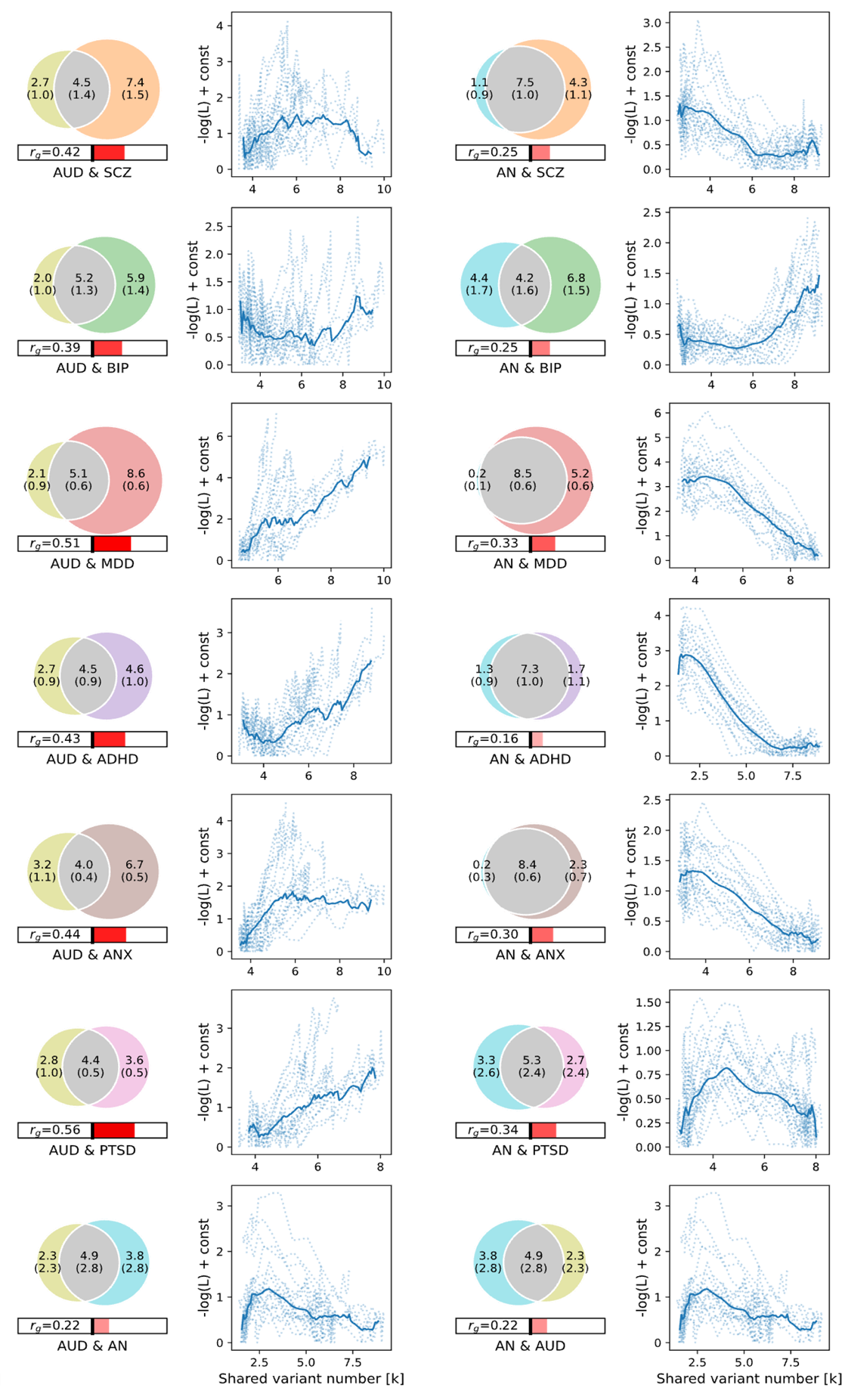

**Supplementary Figure 9. Cross-trait MiXeR results for Alcohol Use Disorder and Anorexia Nervosa.** Venn diagrams of unique and shared polygenic components at the causal level for AUD (left column) and AN (right column). The numbers within the Venn diagrams indicate the estimated quantity of causal variants (in thousands) per component, explaining 90% of SNP heritability in each phenotype, followed by the standard error. The size of the circles reflects the degree of polygenicity. Genetic correlation (rg) is represented in the horizontal bars beneath the Venn diagrams. Right of the central bar (red) indicates positive rg and left of the central bar (blue) indicates negative rg. Next to each Venn diagram is a plot of the log-likelihood of the bivariate fit as a function of 𝜋12 (shared variant) parameter. These curves indicate the strength of the evidence of polygenic overlap as compared to minimal overlap. Positive AIC values (Suppl. Table 5) and a clear minima on the likelihood curves provide evidence of good model fit.

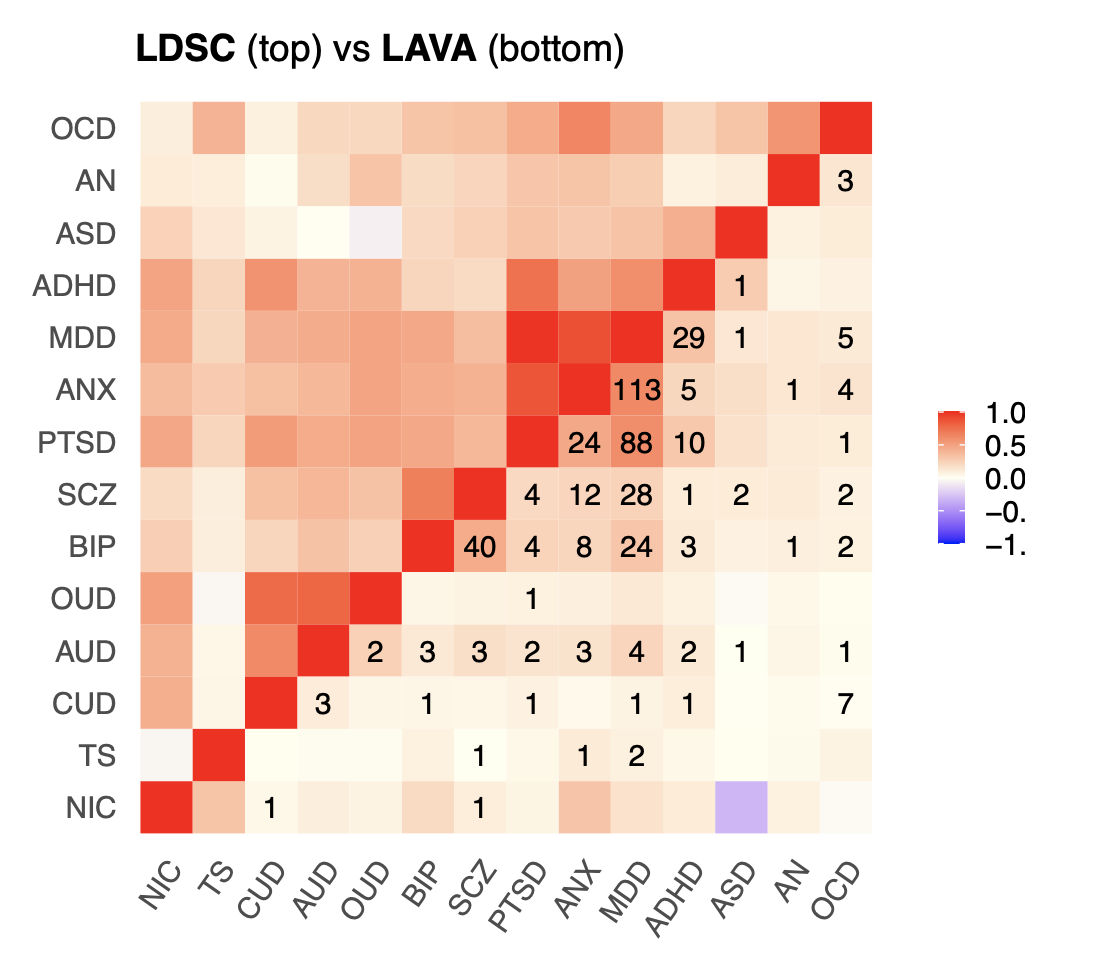

**Supplementary Figure 10. Concordance between global and local genetic correlations.** The global genetic correlations from LDSC have been plotted above the diagonal. This is contrasted against the average local genetic correlations across all tested loci from LAVA below the diagonal. The numbers represent the total number of significant local genetic correlations that were detected.

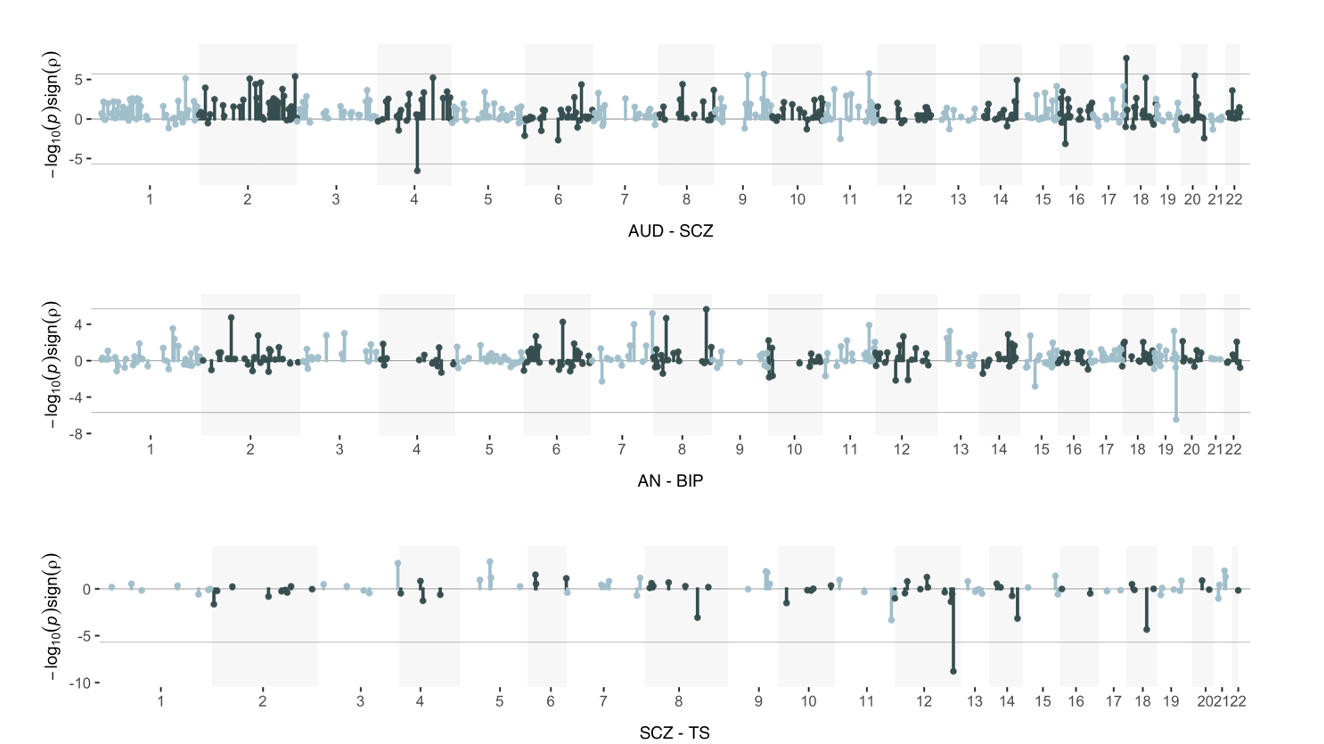

**Supplementary Figure 11. Miami plots of local rg for disorder pairs that exhibited significant negative local rg.** All analyzed loci have been plotted in order of genomic position on the X-axis, binned by chromosome. The Y-axis shows the -log10 *p*-values for all tested local genetic correlations scaled by the direction of the association. The line indicates the Bonferroni corrected significance threshold of 2.1$\times$10^-6^.

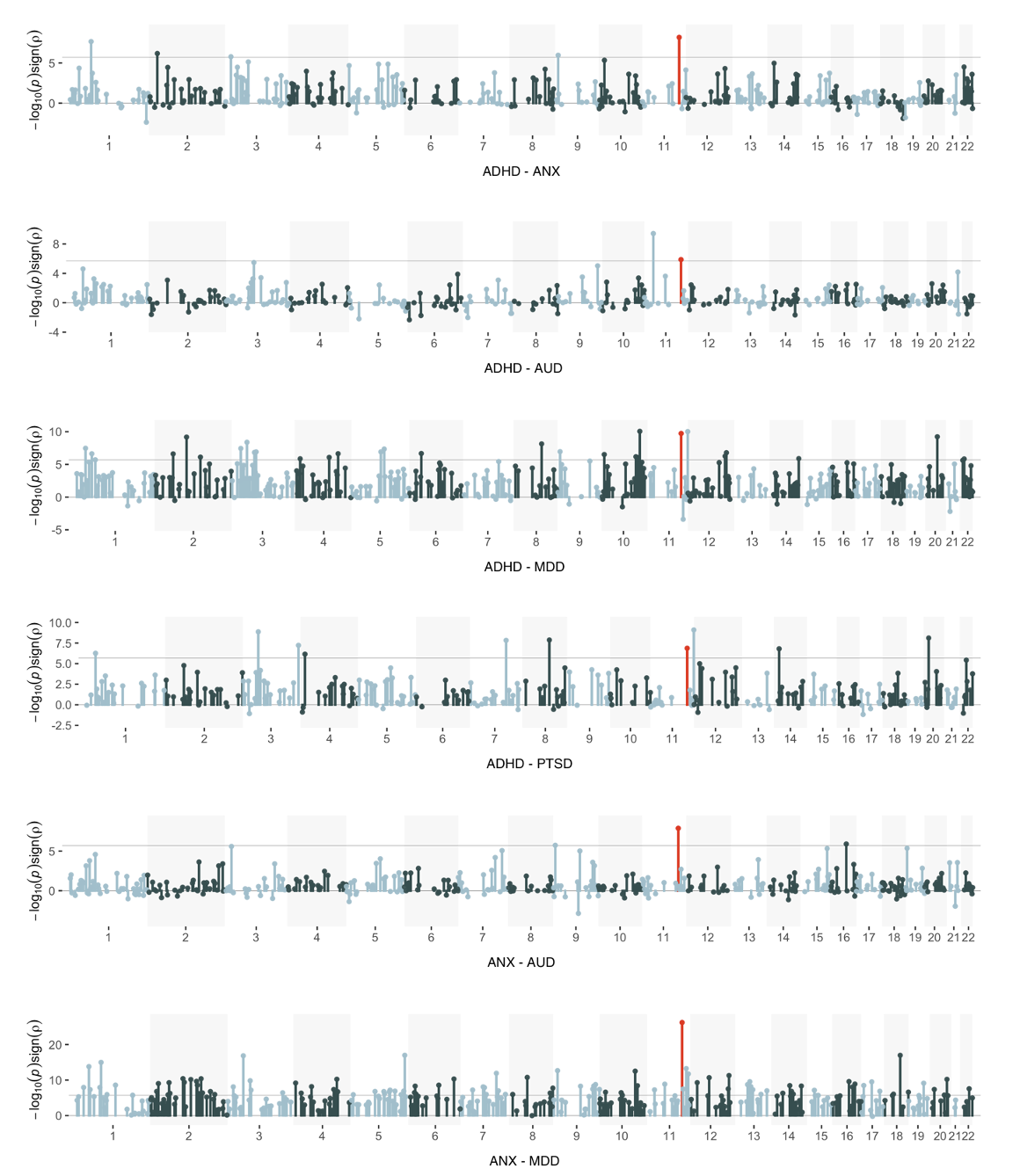
 **Supplementary Figure 12a. Miami plots of local rg for disorder pairs with significant rg correlation in top hotspot locus on chromosome 11.** All analyzed loci have been plotted in order of genomic position on the X-axis, binned by chromosome. The Y-axis shows the -log10 *p*-values for all tested local genetic correlations scaled by the direction of the association. The line indicates the Bonferroni corrected significance threshold of 2.1$\times$10^-6^. The hotspot locus has been colored red.

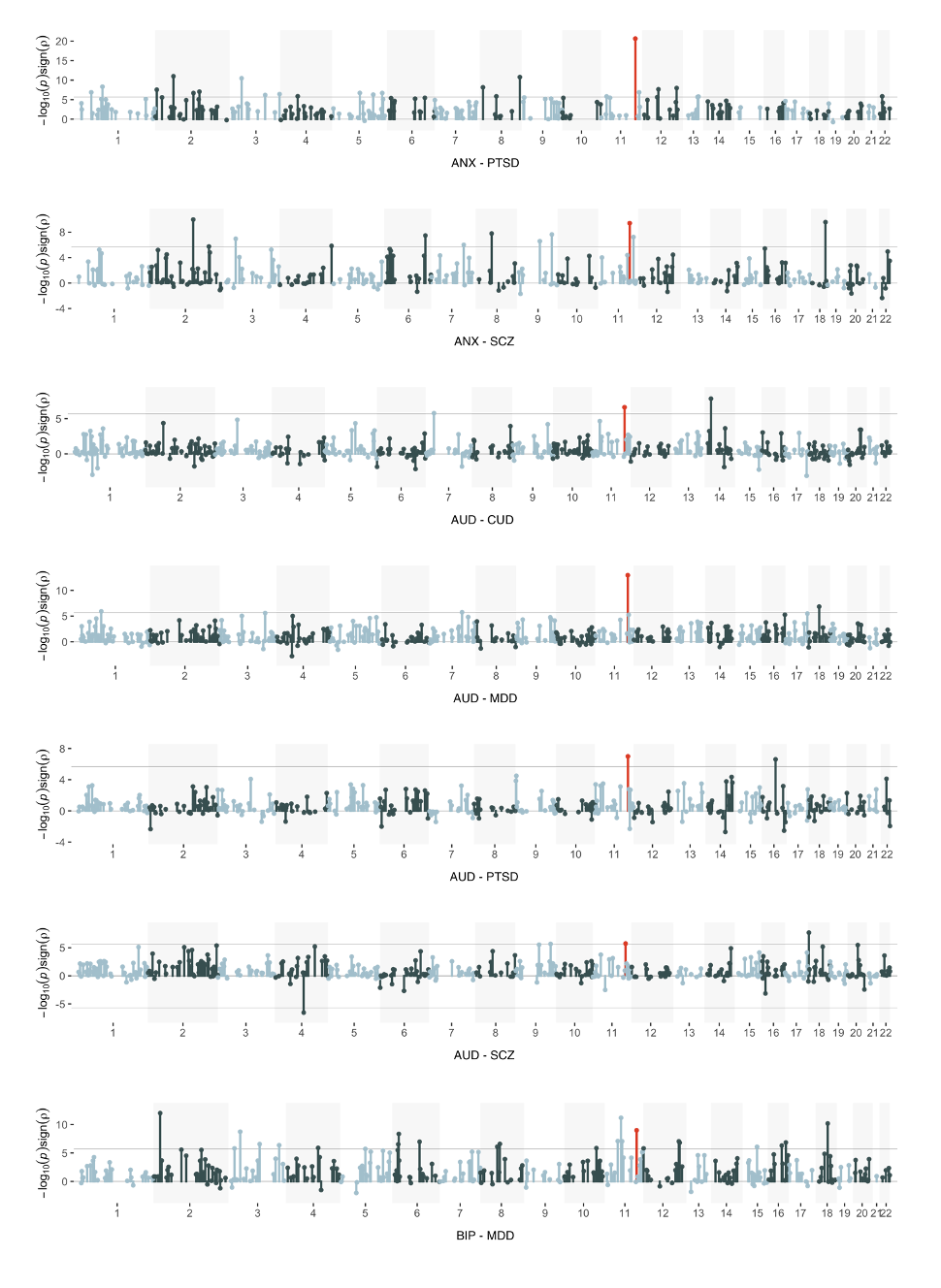

**Supplementary Figure 12b. Miami plots of local rg for disorder pairs with significant rg correlation in top hotspot locus on chromosome 11.** All analyzed loci have been plotted in order of genomic position on the X-axis, binned by chromosome. The Y-axis shows the -log10 *p*-values for all tested local genetic correlations scaled by the direction of the association. The line indicates the Bonferroni corrected significance threshold of 2.1$\times$10^-6^. The hotspot locus has been colored red.

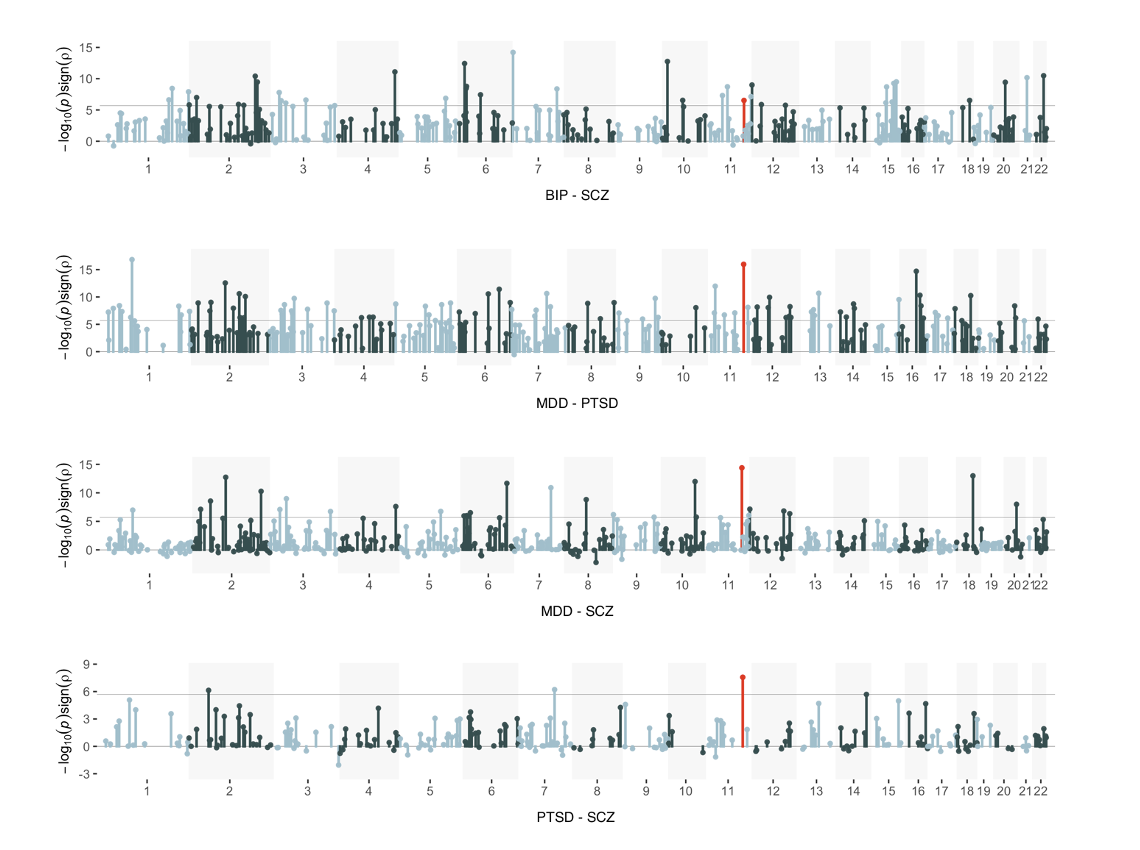

**Supplementary Figure 12c. Miami plots of local rg for disorder pairs with significant rg correlation in top hotspot locus on chromosome 11.** All analyzed loci have been plotted in order of genomic position on the X-axis, binned by chromosome. The Y-axis shows the -log10 *p*-values for all tested local genetic correlations scaled by the direction of the association. The line indicates the Bonferroni corrected significance threshold of 2.1$\times$10^-6^. The hotspot locus has been colored red.

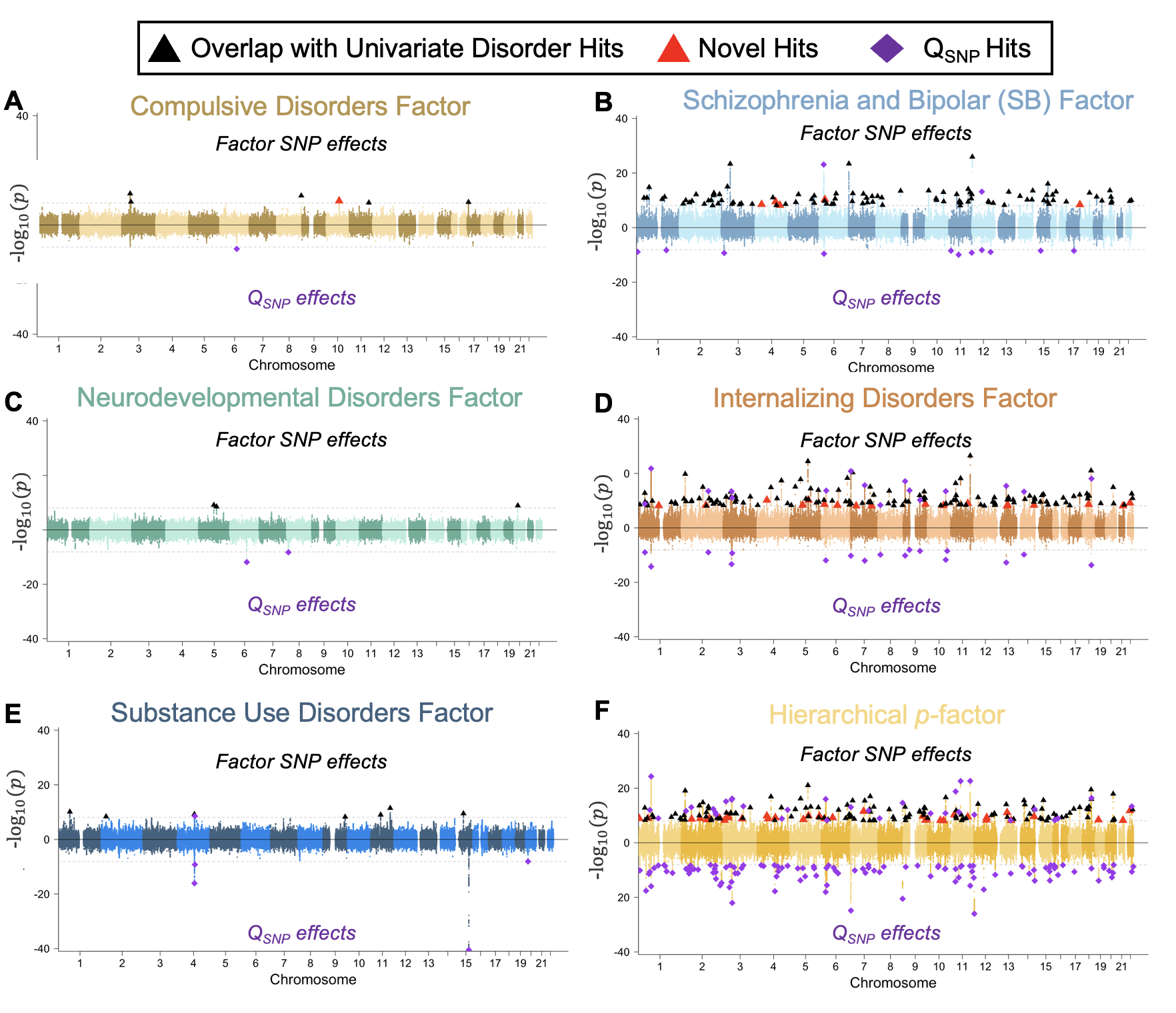

**Supplementary Figure 13. Multivariate GWAS Miami Plots.** *Panels A-E* depict multivariate GWAS results for the five factors from the correlated factors model and *Panel F* the results for the *p-*factor from the hierarchical model. All panels depict results for the -log10(*p*-values) for the factor on the top half of the Miami plot and the log10(*p*-values) for Q_SNP_ on the bottom half. Per the legend at the top of the figure, factors hits that were within 100kb of univariate hits are shown in black triangles, novel hits for the factors that were not within 100kb of a univariate or Q_SNP_ are depicted as red triangles, and Q_SNP_ hits as purple diamonds. The gray dashed line indicates the Bonferroni corrected, genome-wide significance threshold.

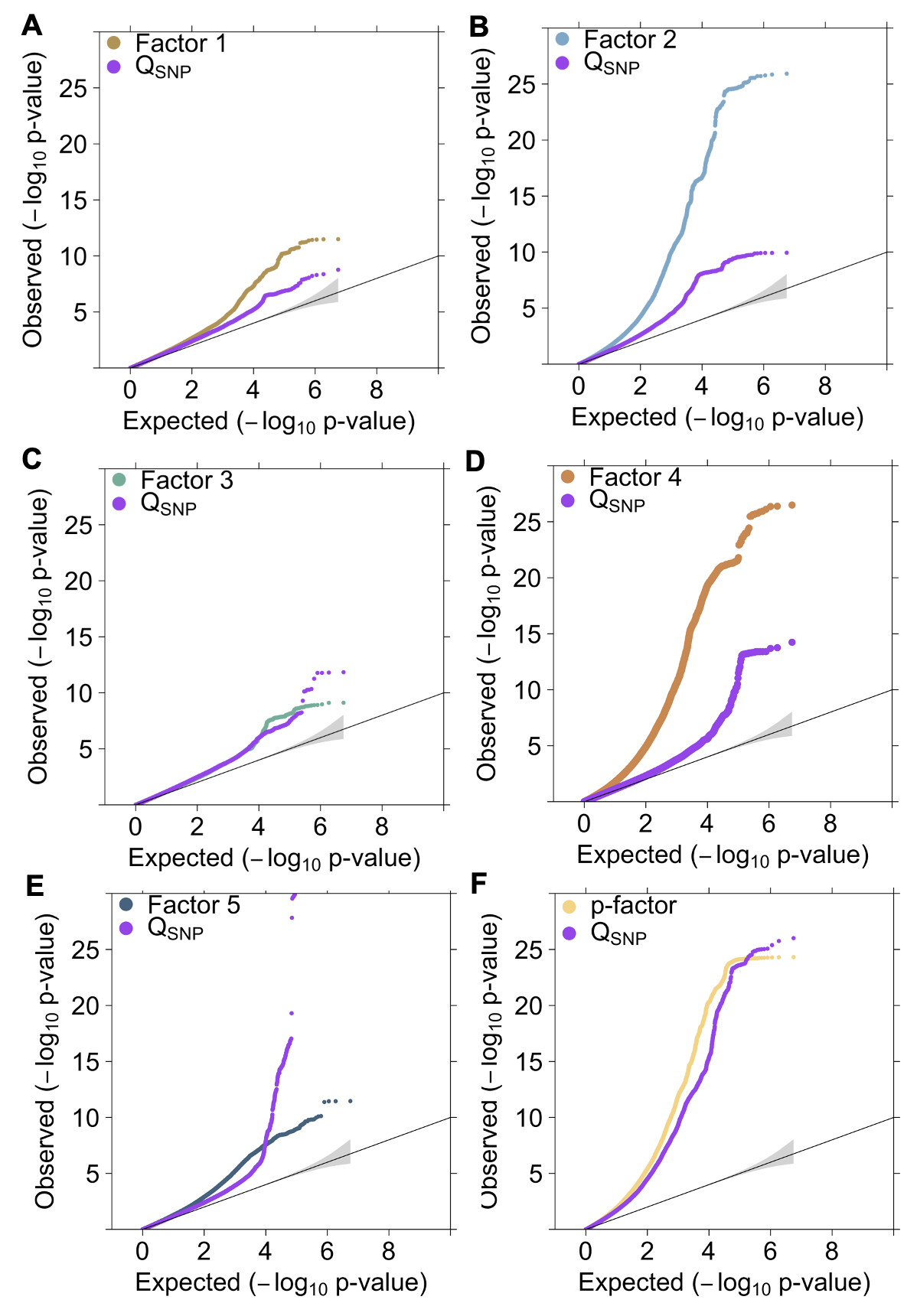

**Supplementary Figure 14. Factor QQ-Plots.** Panels depict the QQ-plots for each of the psychiatric factors from the multivariate GWAS analyses conducted in Genomic SEM. All panels depict factor-specific Q_SNP_ results in purple. *Panels A-E* depict results for the five factors from the correlated factors model and *Panel F* depicts results for the *p*-factor from the hierarchical model.

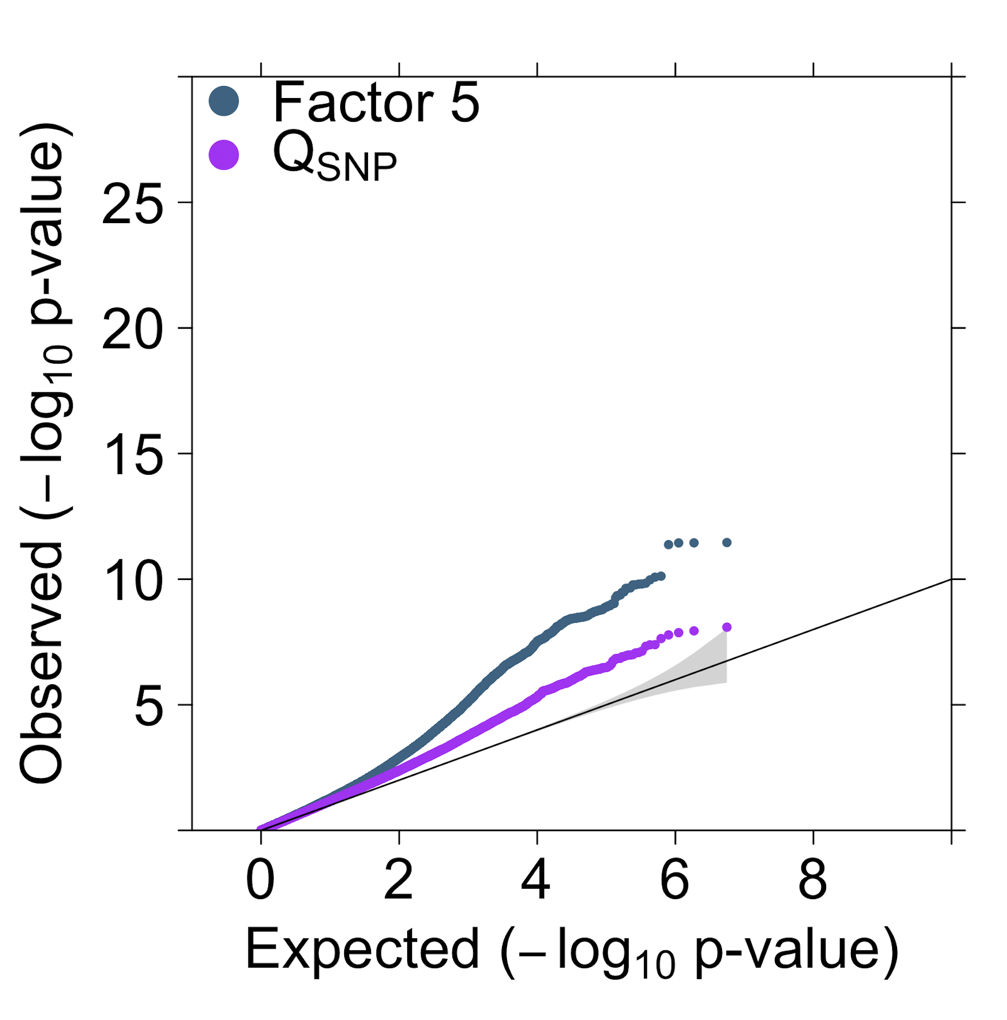

**Supplementary Figure 15. Factor 5 QQ-Plot excluding top Q_SNP_ hits.** Figure depicts the QQ-plot for the substance use disorders when excluding the three loci with the strongest Q_SNP_ signal in the alcohol dehydrogenase and nicotinic acetylcholine receptor genes. When these three loci in substance metabolizing pathways are removed the factor signal (depicted in blue) is stronger relative to Q_SNP_ (depicted in purple).

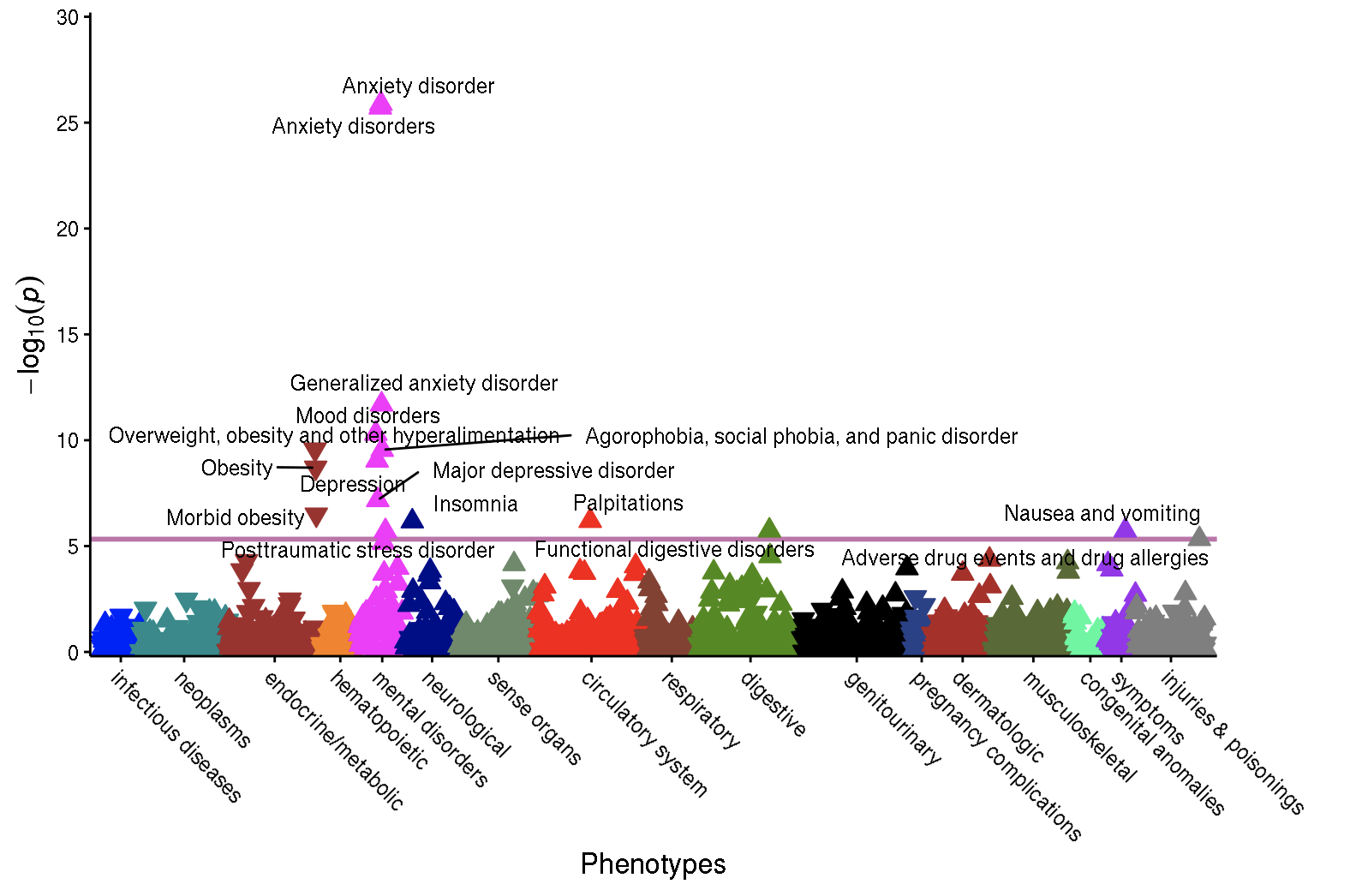

**Supplementary Figure 16a. PheWAS results for the Compulsive Disorders Factor.** Each point reflects associations between the different phecodes in the European genetic ancestry subset of the Mayo Clinic participant sample (*N* = 46,329) and the Compulsive disorders factor polygenic risk score (PRS). Results are grouped and color-coded on the x-axis according to different phenotype groups and vertically positioned on the y-axis according to their -log10(p-values) in the PheWAS. Upward and downward triangles indicate positive and negative PRS-phecode associations, respectively. The purple line indicates the Bonferroni corrected threshold used to define statistical significance in the current analyses (i.e., *p* < 4.7$\times$10^-6^).

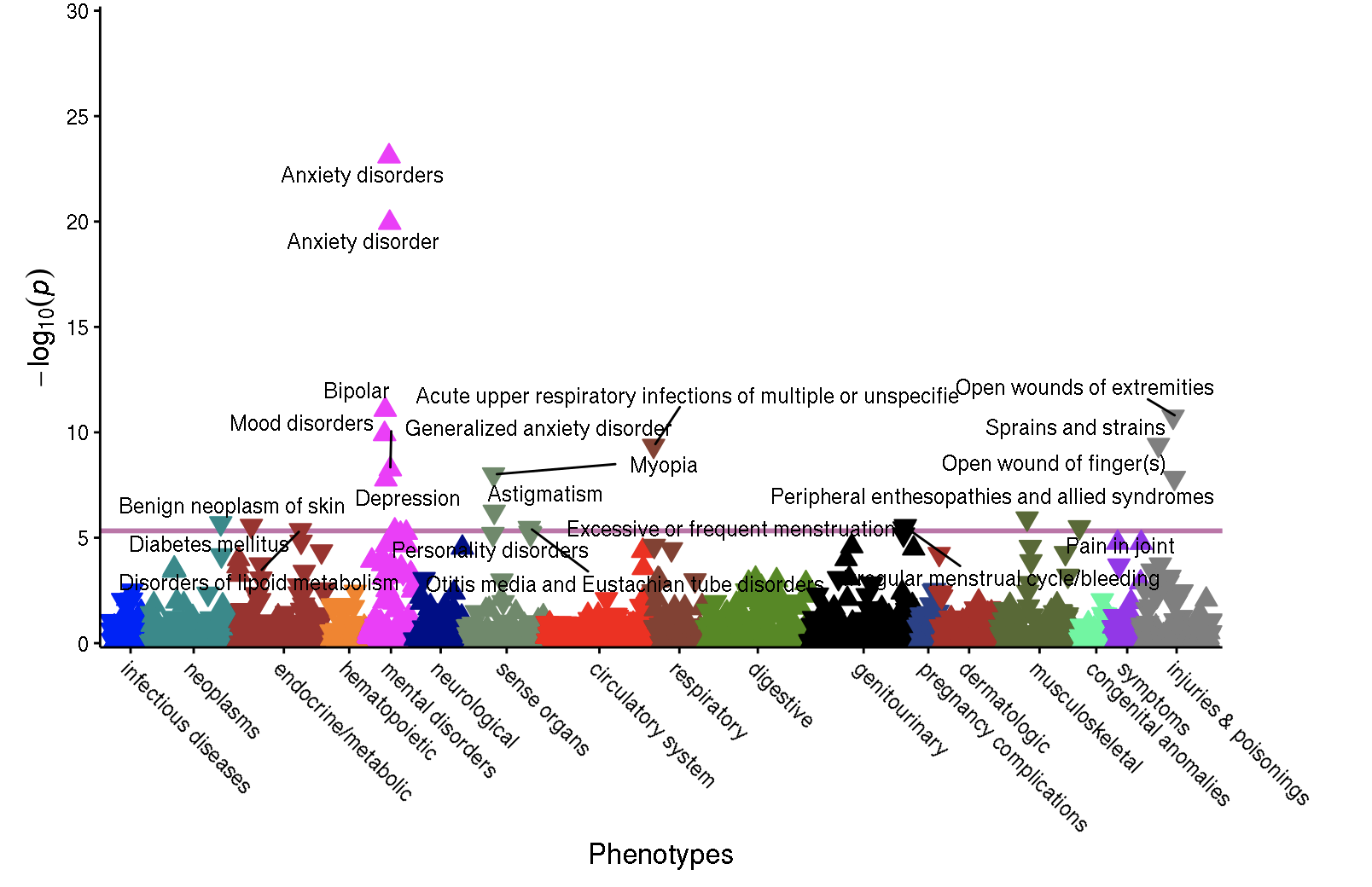

**Supplementary Figure 16b. PheWAS results for the Schizophrenia and Bipolar (SB) Factor.** Each point reflects associations between the different phecodes in the European ancestry subset of the Mayo Clinic participant sample (*N* = 46,329) and the Schizophrenia and Bipolar factor (SB) polygenic risk score (PRS). Results are grouped and color-coded on the x-axis according to different phenotype groups and vertically positioned on the y-axis according to their -log10(p-values) in the PheWAS. Upward and downward triangles indicate positive and negative PRS-phecode associations, respectively. The purple line indicates the Bonferroni corrected threshold used to define statistical significance in the current analyses (i.e., *p* < 4.7$\times$10^-6^).

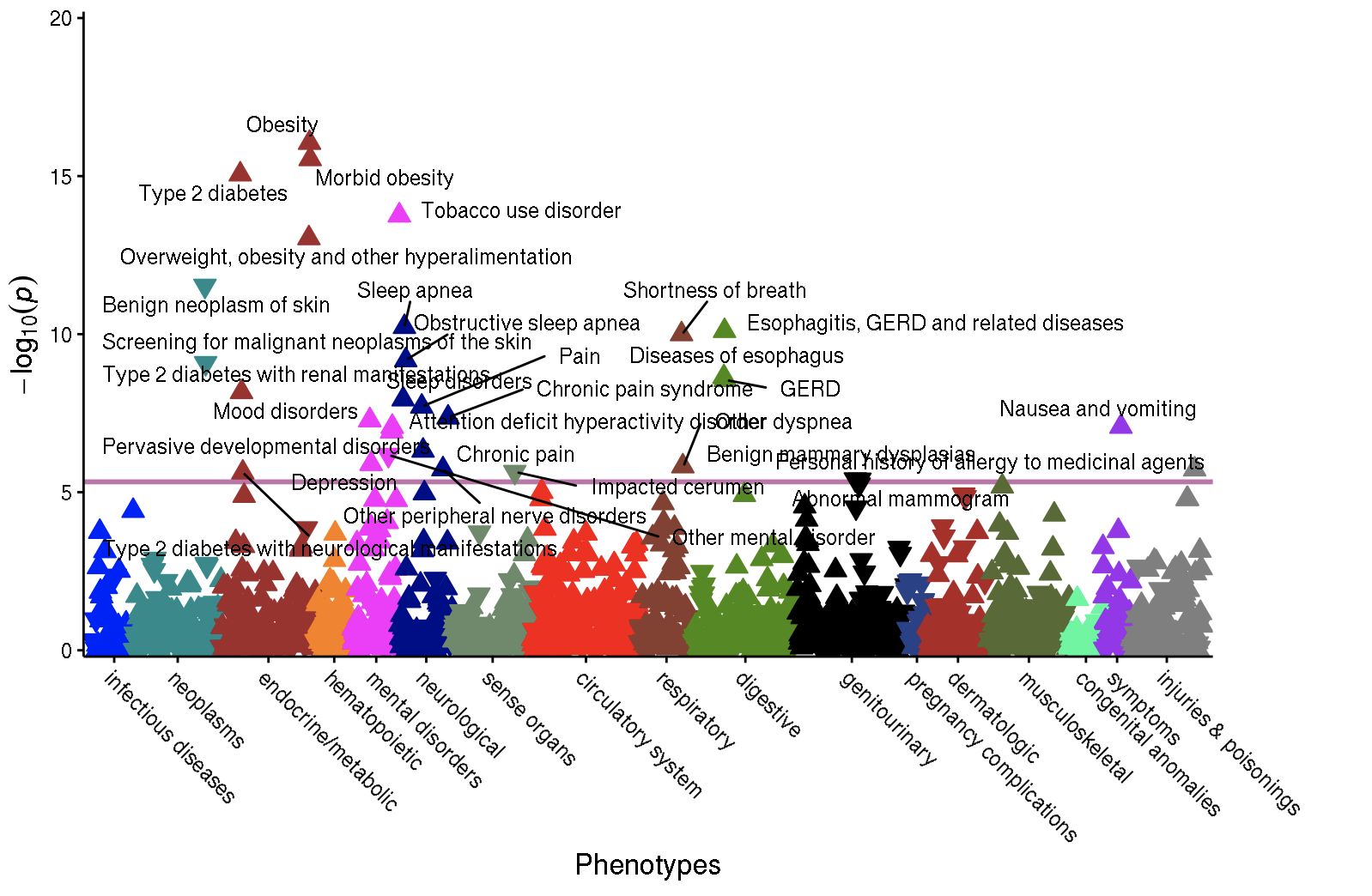

**Supplementary Figure 16c. PheWAS results for the Neurodevelopmental Disorders Factor.** Each point reflects associations between the different phecodes in the European genetic ancestry subset of the Mayo Clinic participant sample (*N* = 46,329) and the Neurodevelopmental disorders factor polygenic risk score (PRS). Results are grouped and color-coded on the x-axis according to different phenotype groups and vertically positioned on the y-axis according to their -log10(p-values) in the PheWAS. Upward and downward triangles indicate positive and negative PRS-phecode associations, respectively. The purple line indicates the Bonferroni corrected threshold used to define statistical significance in the current analyses (i.e., *p* < 4.7$\times$10^-6^).

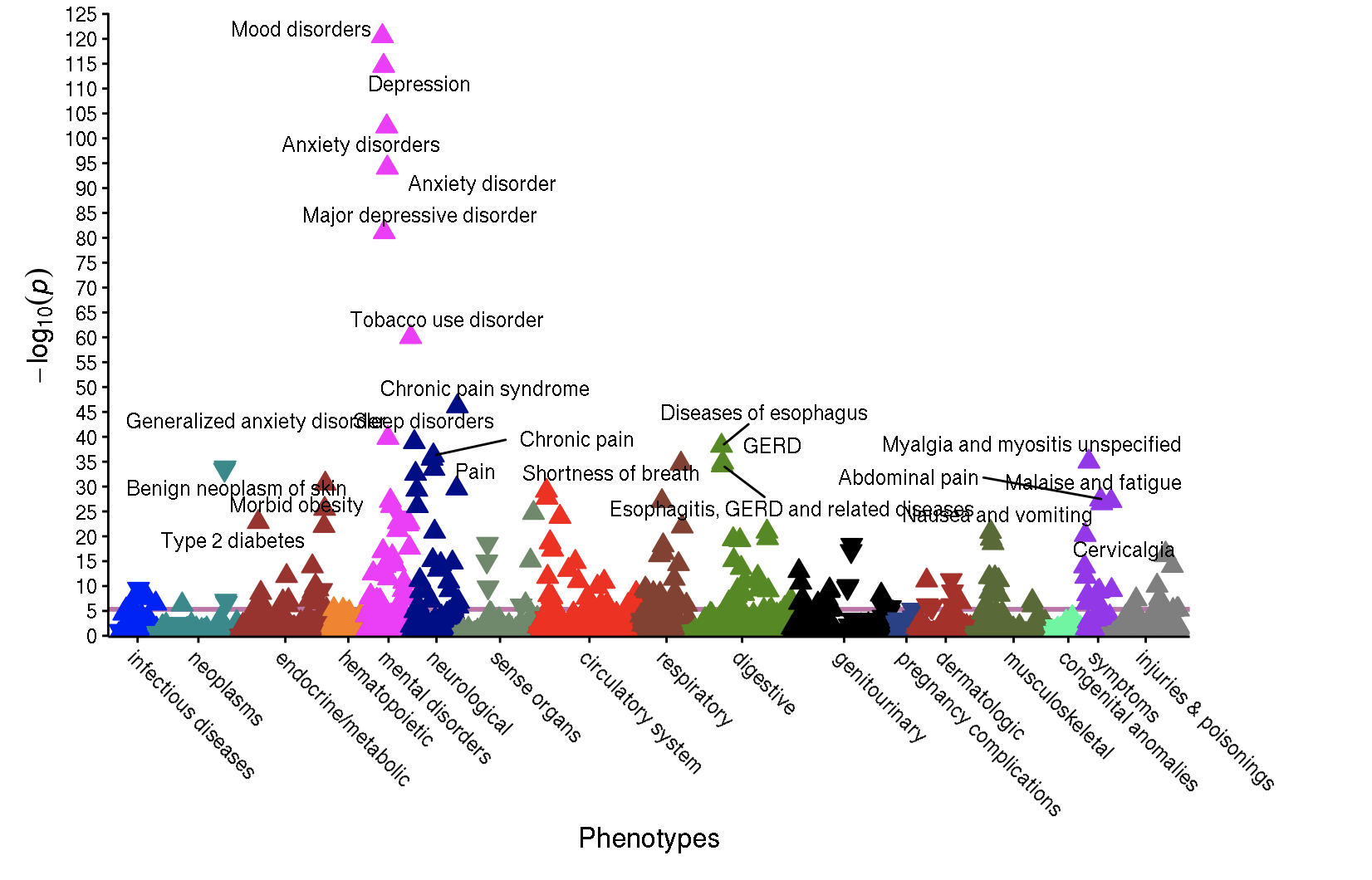

**Supplementary Figure 16d. PheWAS results for the Internalizing Disorders Factor.** Each point reflects associations between the different phecodes in the European ancestry subset of the Mayo Clinic participant sample (*N* = 46,329) and the Internalizing disorders factor polygenic risk score (PRS). Results are grouped and color-coded on the x-axis according to different phenotype groups and vertically positioned on the y-axis according to their -log10(p-values) in the PheWAS. Upward and downward triangles indicate positive and negative PRS-phecode associations, respectively. The purple line indicates the Bonferroni corrected threshold used to define statistical significance in the current analyses (i.e., *p* < 4.7$\times$10^-6^).

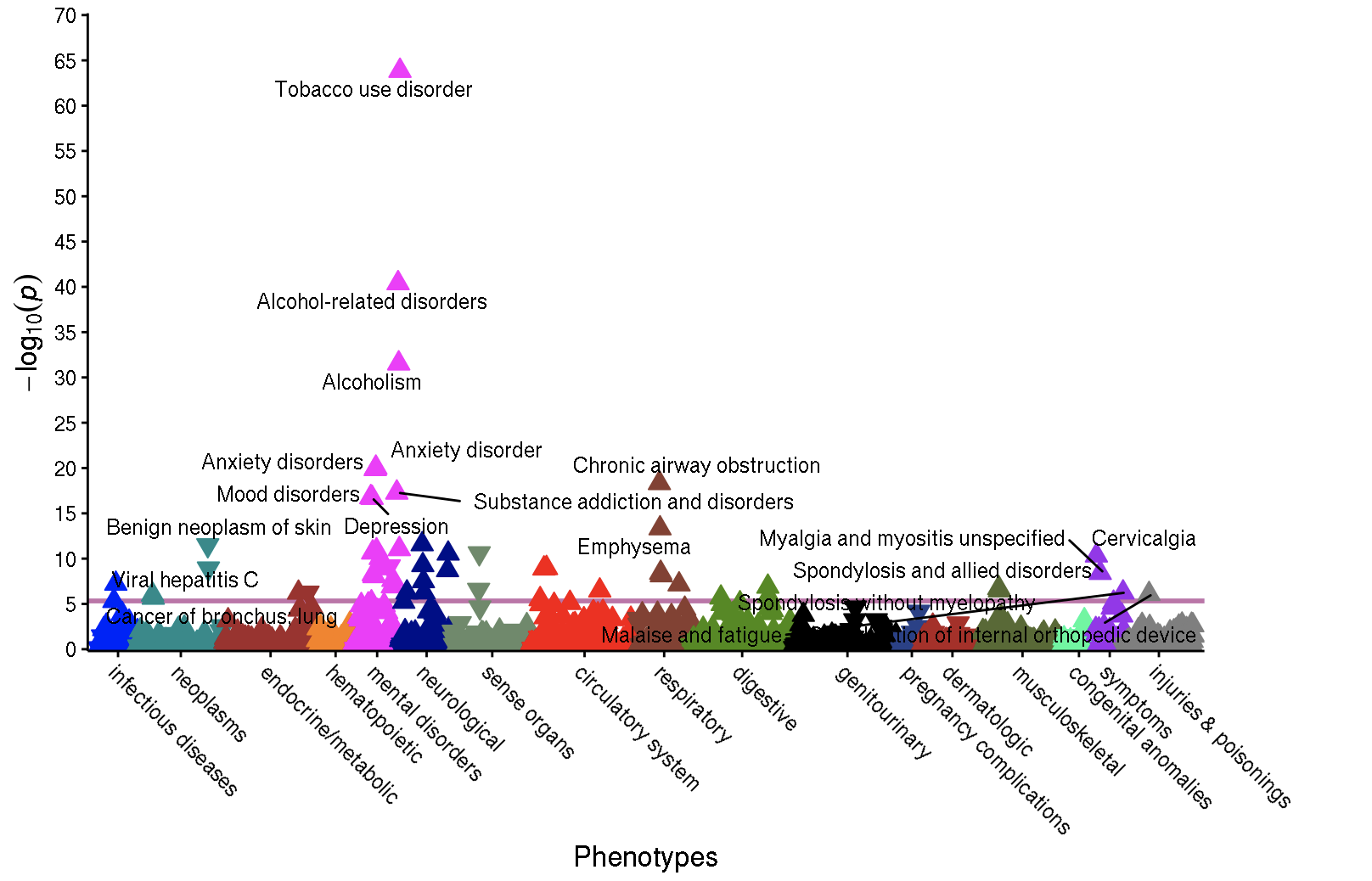

**Supplementary Figure 16e. PheWAS results for the Substance Use Disorders Factor.** Each point reflects associations between the different phecodes in the European genetic ancestry subset of the Mayo Clinic participant sample (*N* = 46,329) and the Substance Use disorders factor polygenic risk score (PRS). Results are grouped and color-coded on the x-axis according to different phenotype groups and vertically positioned on the y-axis according to their -log10(p-values) in the PheWAS. Upward and downward triangles indicate positive and negative PRS-phecode associations, respectively. The purple line indicates the Bonferroni corrected threshold used to define statistical significance in the current analyses (i.e., *p* < 4.7$\times$10^-6^).

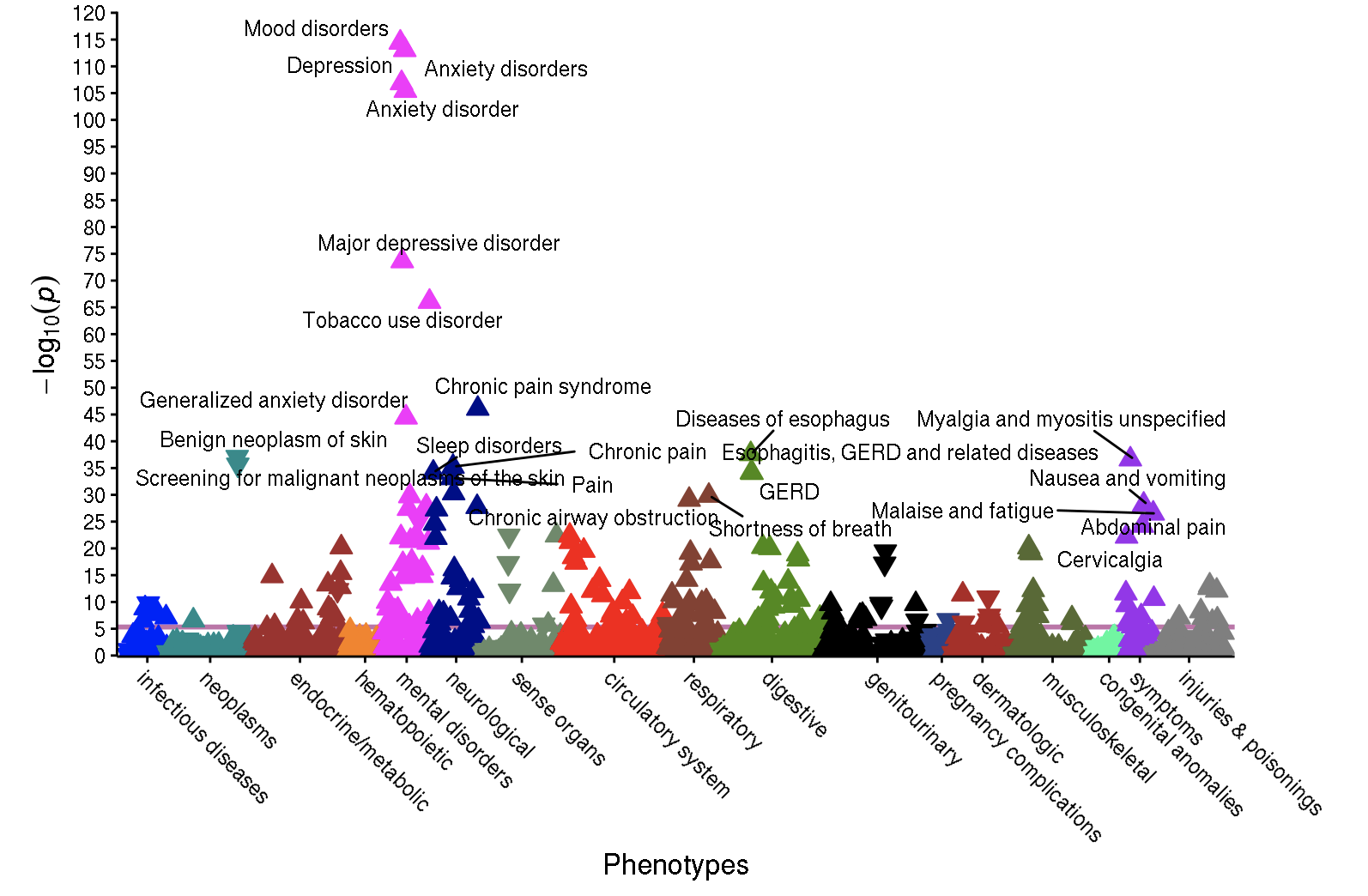

**Supplementary Figure 16f. PheWAS results for the Hierarchical p-Factor.** Each point reflects associations between the different phecodes in the European genetic ancestry subset of the Mayo Clinic participant sample (*N* = 46,329) and the Hierarchical *p*-factor polygenic risk score (PRS). Results are grouped and color-coded on the x-axis according to different phenotype groups and vertically positioned on the y-axis according to their -log10(p-values) in the PheWAS. Upward and downward triangles indicate positive and negative PRS-phecode associations, respectively. The purple line indicates the Bonferroni corrected threshold used to define statistical significance in the current analyses (i.e., *p* < 4.7$\times$10^-6^).

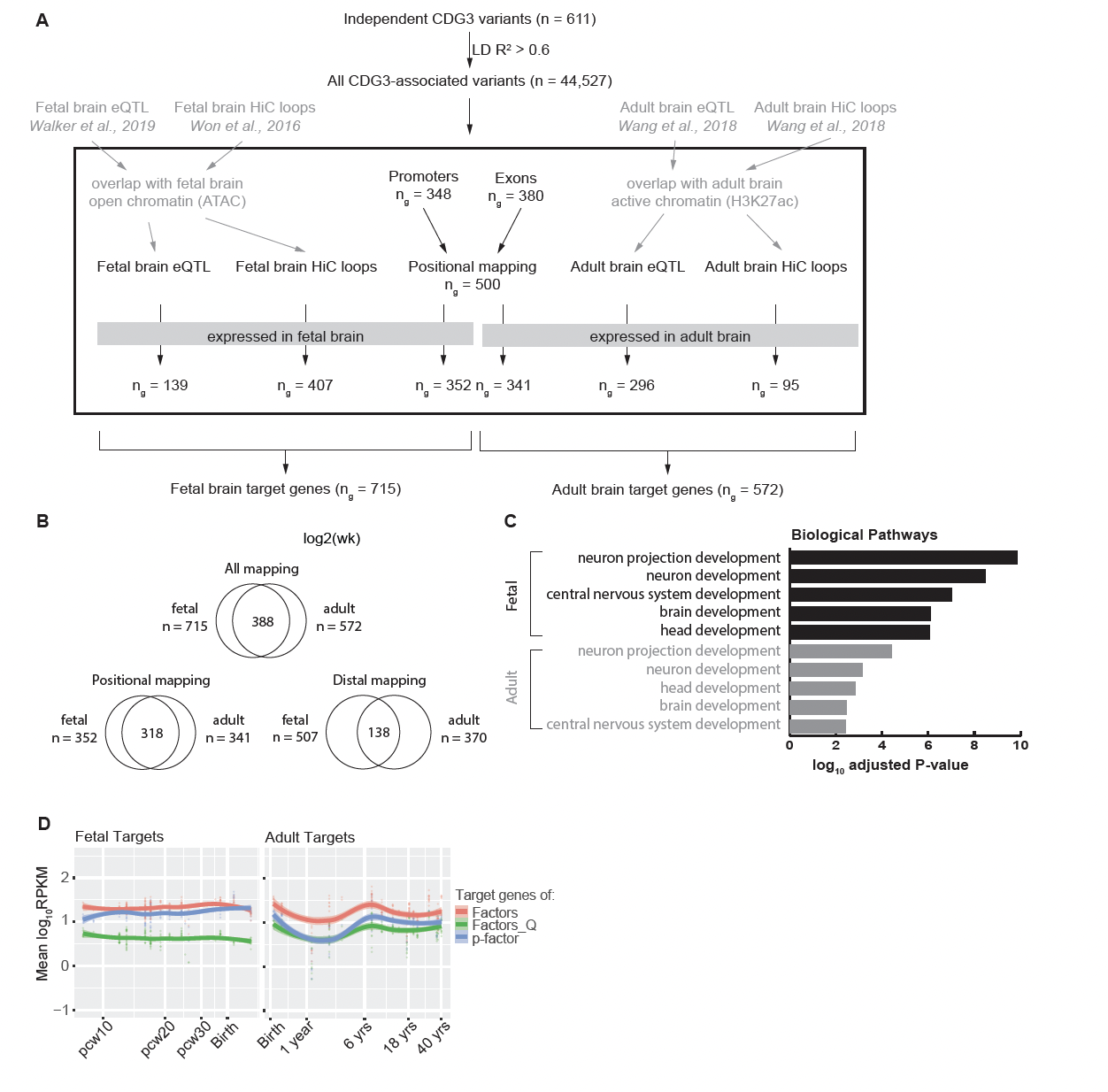

**Supplementary Figure 17. Predicting target genes of CDG variants and temporal expression patterns along the developmental trajectory of the human developing brain. (**A) Schematics showing the procedure of target genes prediction. The independent variants were first expanded in their respective LD block, before either direct mapping based on TSS/exon overlap, or distal mapping with eQTL or HiC loops. The distal mapping was further filtered by corresponding open chromatin data. Ultimately, only genes expressed in the related tissues were retained. See Methods for more details. (B) Overlap of fetal and adult target genes. Most of the overlap is attributable to positional mapping result (bottom-left). See more details in Methods. (C) Gene ontology enrichment analysis of predicted target genes of all the target genes mapped with fetal or adult epigenomics data sets. (D) Averaged and normalized expression levels of target genes of the indicated classes along the temporal trajectory of human brain development. ‘p-factor’, all predicted target genes of class variants for the *p-*factor from the hierarchical model; ‘Factors’, all predicted target genes of variants associated with any of the five psychiatric factors from the correlated factors model; ‘Factors_Q’, all predicted targets of variants associated with any of the 5 factor-specific Q_SNP_ estimates from the correlated factors model.

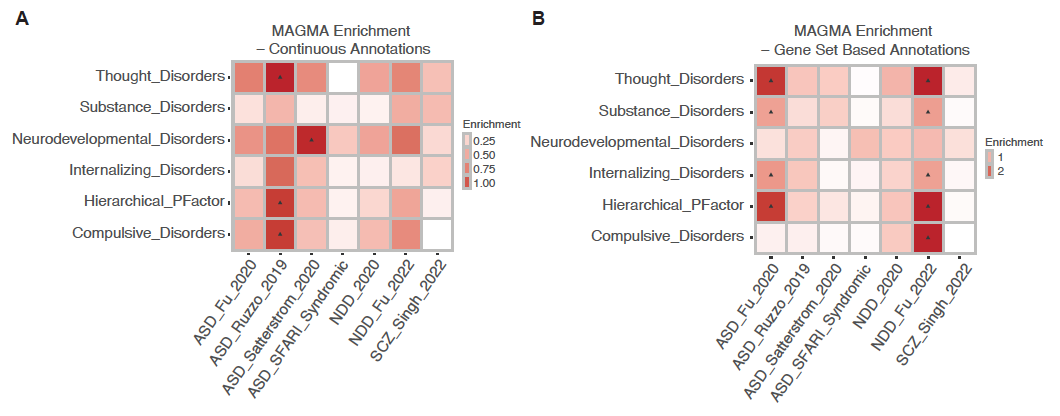

**Supplementary Figure 18. Enrichment of CDG variants near risk genes of related disorders.** (A and B) Enrichment of the indicated variants near genes from the studies indicated using MAGMA. Enrichment using (A) the continuous annotation mode and (B) the gene set based annotation.

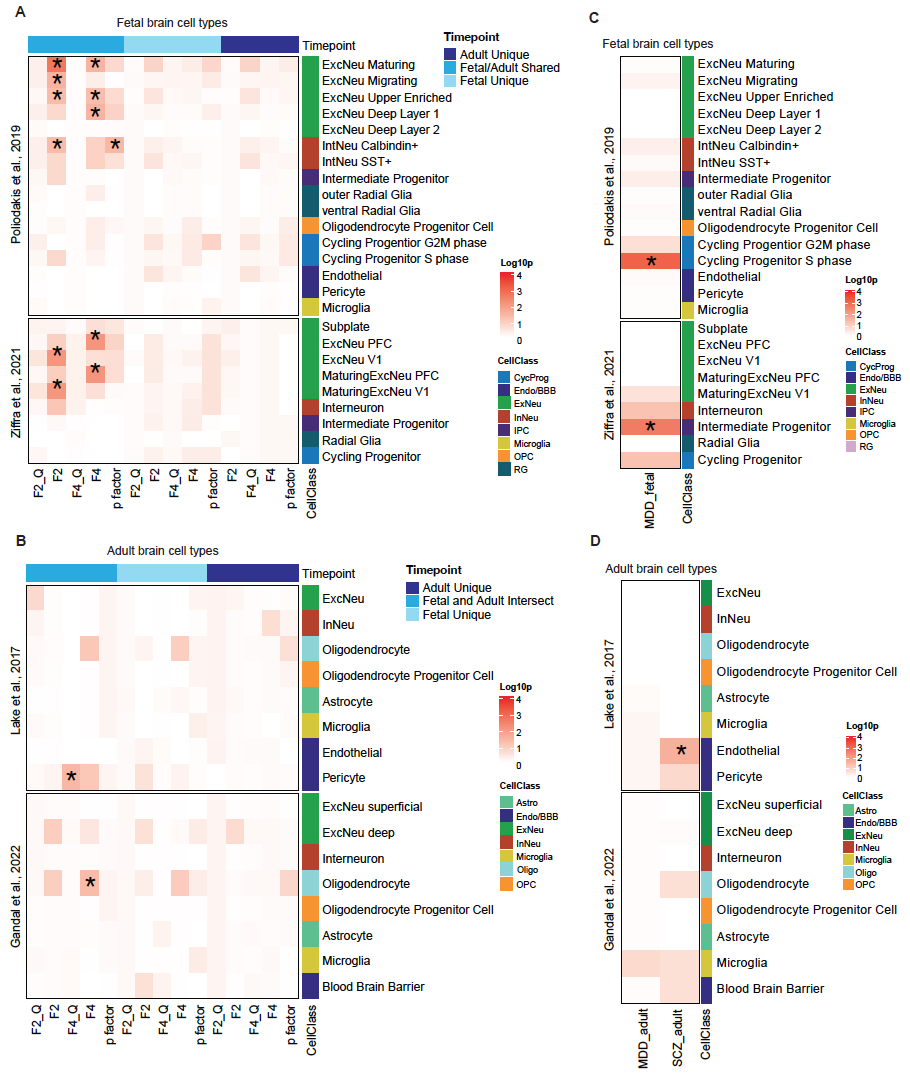

**Supplementary Figure 19. Cell type specificity of CDG variants target genes.** (A and B) Enrichment by expression of CDG3 variants target genes associated with the indicated factors in (A) fetal brain cell types using two independent scRNA-Seq data sets^13,14^ or (B) adult brain cell types using two independent snRNA-Seq data sets.^15,16^ F2_Q = Schizophrenia and Bipolar (SB) Q_SNP;_ F2 = Schizophrenia and Bipolar (SB) (SMI); F4_Q = Internalizing Disorders Q_SNP;_ F4 = Internalizing Disorders. The target genes were segregated into those linked to “Fetal” only, “Adult” only or shared between the two categories. (C and D) Enrichment of target genes of CC-GWAS hits between MD and SCZ based on the single cell data sets as mentioned above. Note that the MD CC-GWAS hits are also MD GWAS hits, and the SCZ CC-GWAS hits are also SCZ GWAS hits.
